# Disruption to diabetes and hypertension care during the COVID-19 pandemic in Latin America and the Caribbean and mitigation approaches: A Scoping Review

**DOI:** 10.1101/2024.04.17.24305997

**Authors:** Oluwabunmi Ogungbe, Samira Barbara Jabakhanji, Roopa Mehta, John McCaffrey, David Byrne, Sinéad Hurley, Lori Rosman, Eyram Cyril Bansah, Folahan Ibukun, Irene Afua Quarshie, Katherine Lord, Yidan Lu, Yunzhi Wang, Asma Rayani, Hairong Liu, Ann Joseph, Alejandro Escobosa, Ivy Nyamuame, Jieun Lee, Ning Meng, Ibrahim Jehanzeb, Temitope Akinyemi, Shoichiro Nohara, Mauro F. F. Mediano, Yvette Yeboah-Kordieh, Cecilia de Sousa, Juliana Farhat, Renato Bandeira de Mello, Tara Taeed, Lawrence J. Appel, Sonia Y. Angell, Edward W. Gregg, Kunihiro Matsushita

## Abstract

**Background:** The COVID-19 pandemic disrupted care for non-communicable diseases globally. This study synthesizes evidence on disruptions to primary care, focusing on hypertension and diabetes care and mitigation approaches taken during the pandemic in Latin America and the Caribbean (LAC).

**Methods:** We conducted a scoping review, searching nine electronic databases for studies from January 2020 to December 2022 on COVID-19-related primary care disruptions and interventions, including studies on hospital-based interventions given their relevance to the pandemic response in LAC. We adapted the Primary Health Care Performance Initiative framework to develop our search strategy and synthesize data. For studies reporting interventions, we included studies conducted outside of LAC.

**Findings:** Of 33,510 references screened, 388 studies were included (259 reported disruptions in LAC, 61 interventions in LAC, 63 interventions outside LAC, and five interventions from countries within and outside LAC), with three-quarters presenting data from Brazil, Argentina, Mexico, and Peru; few studies focused on rural areas. Additionally, the few studies that adequately quantified care disruptions reported a reduction in hypertension and diabetes control during the pandemic (e.g., hypertension control rate decreased from 68% to 55% in Mexico). Frequently reported causes of disruption included burnout and mental health challenges among healthcare workers (with disproportionate effects by type of worker), reduced medication supplies, and reduced frequency of clinic visits by patients (e.g., due to financial constraints). The most reported interventions included remote care strategies (e.g., smartphone applications, virtual meeting platforms) and mental health programs for healthcare workers. Remote care strategies were deemed feasible for care delivery, triaging, and clinical support for non-physicians. Patients were generally satisfied with telemedicine, whereas providers had mixed perceptions. Robust evidence on the effectiveness of remote care strategies for diabetes and hypertension care was unavailable in LAC.

**Interpretation:** Hypertension and diabetes control appeared to worsen in LAC during the pandemic. Major reported causes of care disruptions were workforce issues, reduced medication supply, and changes in patient perceptions of seeking and receiving primary healthcare. Remote care strategies were feasible for various purposes and were well received by patients. However, the lack of data on intervention effectiveness underscores the importance of strengthening research capacity to generate robust evidence during future pandemics. Developing resilient healthcare systems able to provide care for hypertension and diabetes during future pandemics will depend on investment in the healthcare workforce, medical supply chain, health data and research infrastructure, and technology readiness.

**Funding:** This work was supported by funding from the World Bank to Johns Hopkins Bloomberg School of Public Health and RCSI University of Medicine and Health Sciences. Additional support to RCSI was provided by Science Foundation Ireland, Converge Programme, grant number 22/RP/10091.

---

Panel: Research in context

*Evidence before this study*

While evidence on health system impacts of the COVID-19 pandemic and potential interventions has been emerging rapidly, comprehensive syntheses in Latin America and the Caribbean (LAC) were unavailable.

*Added value of this study*

Drawing from 388 studies on primary care disruptions in LAC and corresponding interventions in and outside of LAC, this scoping review describes pandemic disruptions and mitigation interventions for diabetes and hypertension care from 2020 to 2022. Differences were identified in the volume of studies describing care disruptions and interventions by country/geography, and by the domain of primary care provision. Most publications were from Brazil, Mexico, Argentina, Peru, and urban settings. Most disruptions were identified in the workforce, medication supplies, and patient perceptions of care. Mitigation responses predominantly included remote care strategies and mental health programs for the health workforce; however, robust data for their effectiveness in LAC were unavailable.

*Implications based upon the available evidence*

This scoping review shows that action is needed to strengthen healthcare systems and their resilience in LAC. Research gaps remain in many countries and healthcare domains, a major gap being the lack of studies investigating intervention effectiveness. To provide care for persons with hypertension and diabetes during the next pandemic, countries in LAC should invest in the healthcare workforce, health data and research infrastructure, and technology readiness.

## 1 Introduction

The COVID-19 pandemic disrupted care for non-communicable diseases (NCDs) like hypertension and diabetes around the world. A negative impact on the management of these conditions in Latin America and the Caribbean (LAC) was likely, as the region has some of the highest rates of chronic diseases globally, and a need for improvements to NCD care delivery was recognized before the pandemic.^1–3^ ^4^

In 2021, Pan American Health Organization (PAHO) surveys across LAC found disruptions in the delivery of about half of essential healthcare services, including hypertension and diabetes management and prescriptions.^5^ A study using household surveys in 14 LAC countries found that healthcare disruption was highest at the beginning of the pandemic in May-June 2020, with an average of 20.4% of households reporting disruption, and declined to 2.9% by May-July 2021.^6^ Furthermore, a qualitative study with health system decision-makers in 8 LAC countries found that the main mitigation strategies focused on telemedicine, community-based care, expansion of hospital capacity and health workforce, and public-private collaborations. However, the magnitude of care disruptions and recovery approaches have yet to be systematically characterized.^7^

We conducted a scoping review to answer two research questions: 1) To what extent were diabetes and hypertension care in the LAC region disrupted during the COVID-19 pandemic? and 2) Which interventions were shown to mitigate, reverse, or support the recovery of diabetes and hypertension care disruptions due to the pandemic? Our main focus was diabetes and hypertension since they are leading causes of mortality worldwide and often considered pathfinders for reforming primary care systems. Based on findings from the present scoping review, we provide potential actions to strengthen healthcare systems capable of providing care during future pandemics.

## 2 Methods

This scoping review adheres to the PRISMA Extension for Scoping Reviews (**appendix 1 pp 1-2**).^8^ Before commencing this review, we modified the Primary Health Care Performance Initiative (PHCPI) Framework and used it to organize our analysis. The PHCPI Framework includes various domains (e.g., service delivery) and sub-domains (e.g., facility organization and management) of healthcare provision (**appendix 1 pp 3**). We have published our study protocol^9^ and summarized key review components below and in **Table 1**.

**Table 1.** Scoping review stages and description.

| **Scoping Review Steps** | **Description** |
| --- | --- |
| Conceptual framework | We adapted the Primary Health Care Performance Initiative Framework<sup>5</sup> (**appendix p.3**) |
| Search strategy | Detailed in **reference 5**, and search terms listed in **appendix p.4-5** |
| Inclusion/exclusion criteria | Detailed in **reference 5**, and search terms listed in **appendix p.6** |
| Title/Abstract screening | Dual reviewers with a third reviewer if inconsistent screening |
| Full-text review | Dual reviewers with a third reviewer if inconsistent review |
| Data extraction | Dual reviewers with a third reviewer if inconsistent extraction (**appendix 7-8**) |
| Quality Assessment | Dual reviewers with a third reviewer if inconsistent quality assessment (**appendix p.9-12**). |

### 2.1 Search strategy and information sources

Guided by the Population, Concept, and Context (PCC) framework and the modified PHCPI Framework (**appendix 1 pp 3-4**), we iteratively developed the search strategy (**appendix 1 pp 5-6**) with an information specialist (L.R.). While the PHCPI framework focuses on primary care, we also included studies on hospital-based interventions, given their relevance to the pandemic response in LAC. We searched MEDLINE via PubMed, CINAHL, Global Health, Embase, Cochrane, Scopus, Web of Science, and LILACS (Latin American and Caribbean Health Sciences Literature) from January 1, 2020, until December 12, 2022. The World Bank, PAHO, and four content experts from Barbados, Brazil, Mexico, and Peru recommended grey literature.

### 2.2 Study selection and eligibility

After removing duplication,^10,11^ two reviewers independently screened each title/abstract against specified criteria and reviewed potentially eligible full-text publications using Covidence® software. Conflicts were resolved by a third reviewer or team consensus. For the eligibility (**appendix 1 pp 3**), the *population* of interest is all adults eligible for screening or receiving diabetes or hypertension care and children with diabetes or hypertension. The *concept* is a disruption to the delivery of primary care services and mitigation, reversal, or recovery of those services where interruptions occur. While prioritizing diabetes and hypertension care, we also included literature on other primary care services. The *context* is LAC during the COVID-19 pandemic. For literature on interventions, we also included studies conducted outside LAC since their results may inform healthcare reform in LAC. Original studies, reports, and reviews with systematic search strategies in English, Spanish, or Portuguese were included.

### 2.3 Data extraction

A data extraction and quality assessment template was developed and embedded in Covidence® to record study information and to map studies according to the modified PHCPI Framework (**appendix 1 pp 7-8).** Two independent reviewers extracted data and assessed study quality, and a third reviewer contributed to establishing consensus when needed. Studies were assessed for quality using the Mixed Methods Assessment Tool (MMAT),^12^ due to the heterogeneity of study designs represented.

### 2.4 Data synthesis

We synthesized study findings narratively. As telemedicine was the most common type of intervention identified, we categorized findings by feasibility and utilization, acceptability and satisfaction, and effectiveness.^14^ MMAT results were displayed in a traffic plot using the *robvis* tool (**appendix 1 pp 9-23**).^13^

### 2.5 Developing a list of potential actions

Together with PAHO, we established a panel of content experts from LAC. We shared preliminary study findings with the panel, World Bank, and PAHO representatives and received initial feedback in May/June 2023. Using the final scoping review findings, we developed a list of potential actions and refined these based on feedback from the expert panel, World Bank, and PAHO.

### 2.6 Reflexivity

Our team includes authors who currently or have previously worked in LAC countries (RM, KL, AJ, AE, IN, TA, MFFM, CS, JF, RBM, SYA), as well as senior researchers with experience in international collaborations (KM, EWG, LJA, SYA). We approached this study with the intention of equitable inclusion of researchers with contextual knowledge of the region (**appendix 2**).^15^

### 2.7 Role of the funding source

This project was supported by the World Bank, and additional support to investigators was provided by Science Foundation Ireland. The expert panel was supported by PAHO. Although the World Bank, PAHO, and the expert panel had opportunities to review relevant materials and provide feedback, the content of the present study is solely the responsibility of the authors and may not represent the views of the World Bank, PAHO, Science Foundation Ireland, or the expert panel members.

## 3 Results

Our search identified 49,954 records (**Figure 1**). After removing 16,441 duplicates, 33,510 titles/abstracts were screened for eligibility, and 1,511 remained for full-text review. Of these, 388 studies were included in this review, 259 reporting care disruptions in LAC and 129 exploring interventions within and outside LAC (**Table 2, appendix 1 pp 24**). Of the 388 studies, most studies were from Brazil (n=169), Argentina (n=45), Mexico (n=44), and Peru (n=35); 59% (n=228) were written in English only (**Figure 2**, **Table 2**).

**Figure 1.**
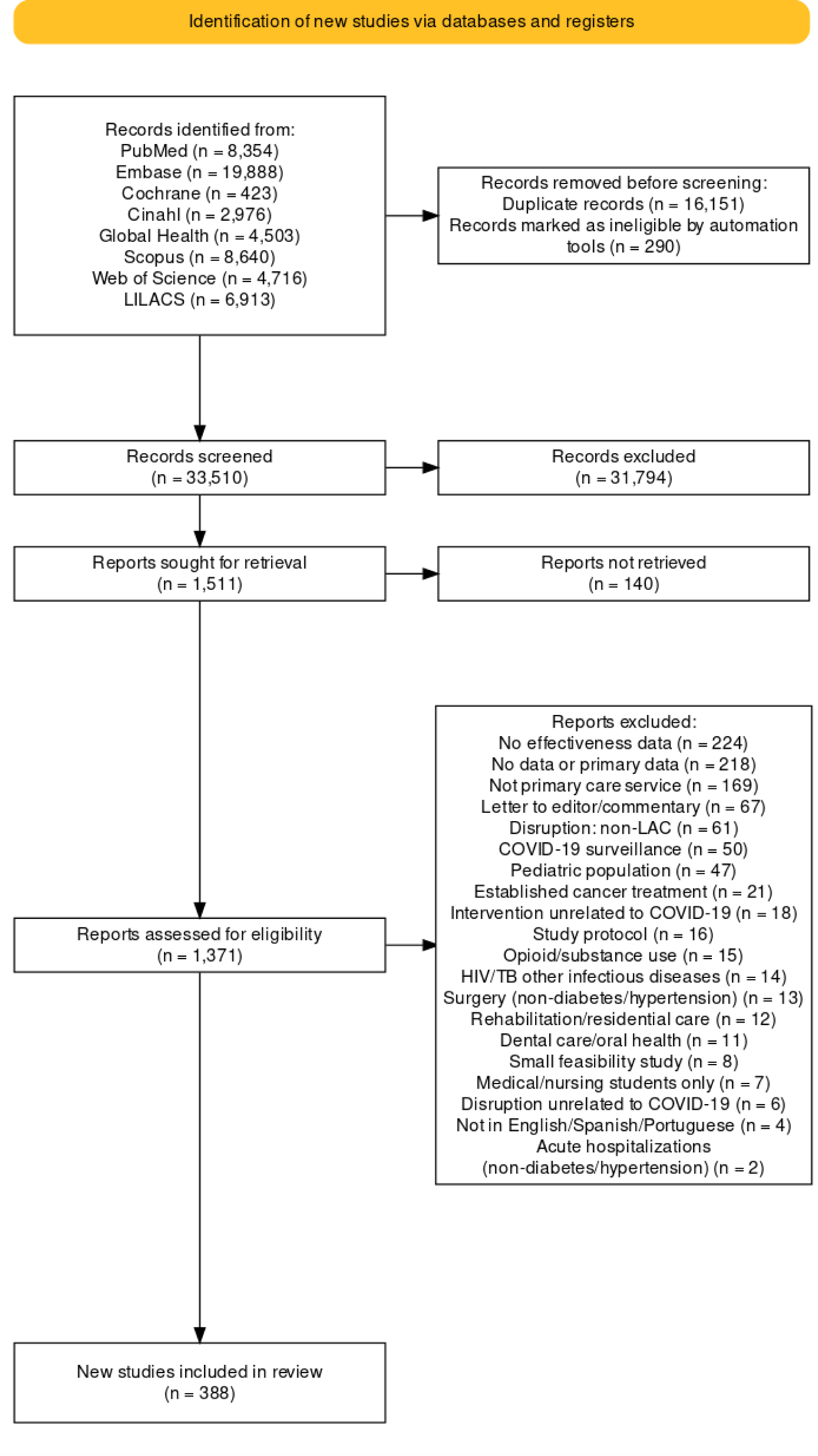
Selection of eligible studies, PRISMA flowchart^1^.

**Figure 2.**
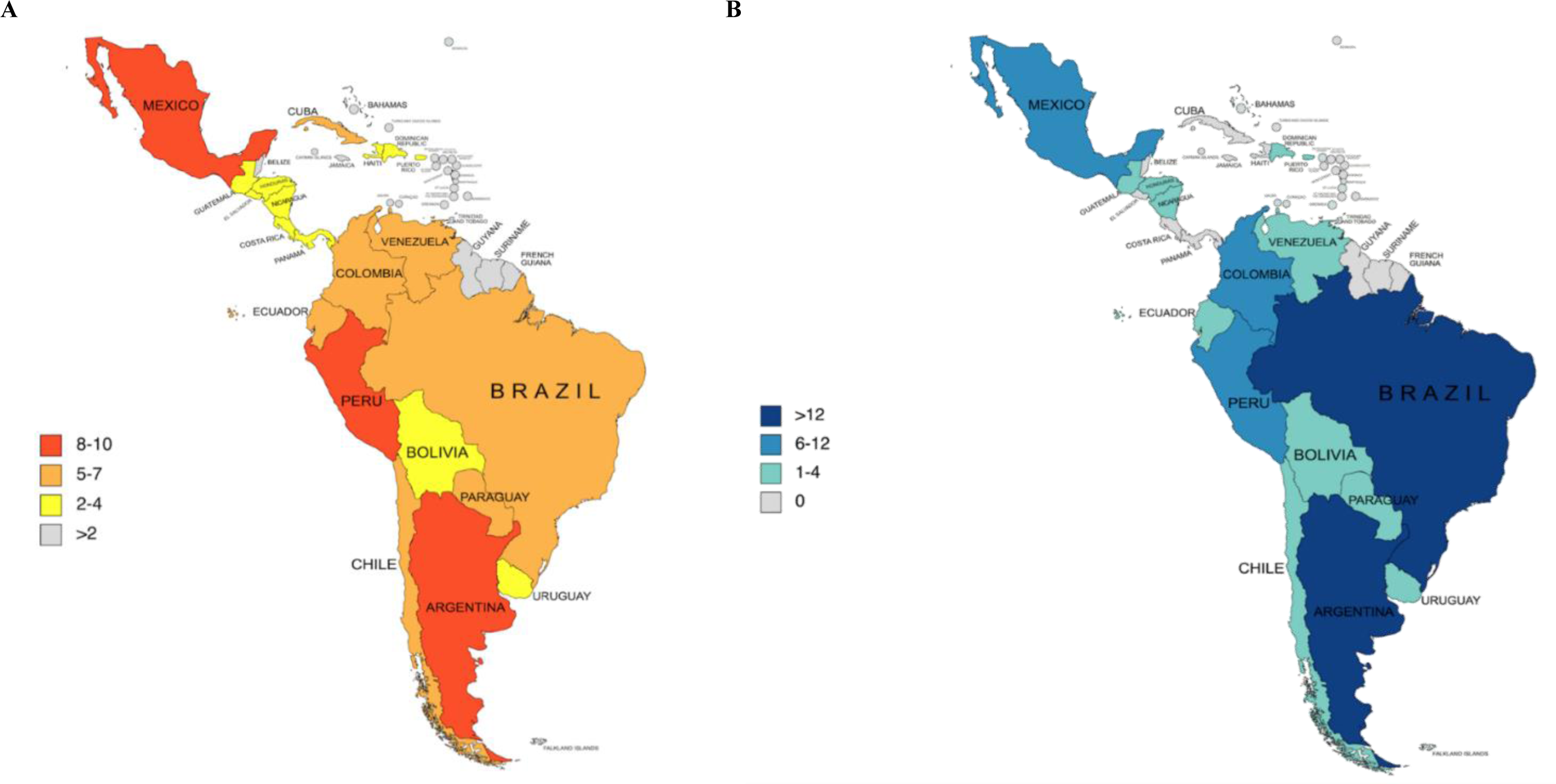
Geographical representation of included studies within Latin America and the Caribbean (LAC) on (A) disruptions to primary care; (B) interventions to mitigate disruptions.

**Table 2.**
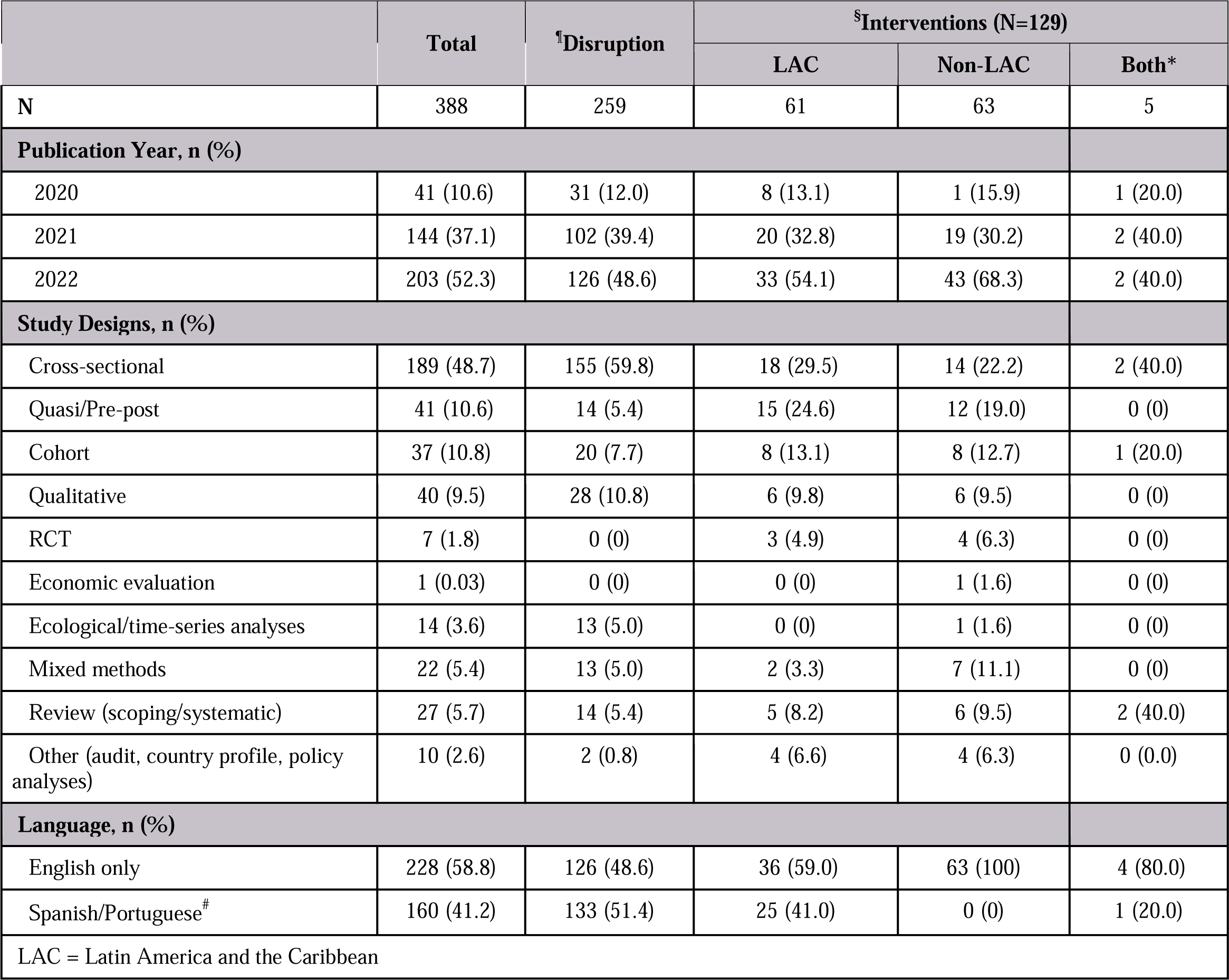
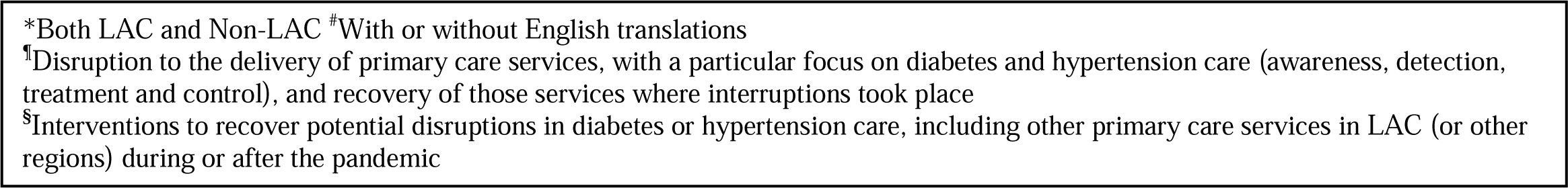
Characteristics of included studies.

|  | Total | <sup>†</sup>Disruption | <sup>§</sup>Interventions (N=129) |  |  |
| --- | --- | --- | --- | --- | --- |
|  |  |  | LAC | Non-LAC | Both* |
| N | 388 | 259 | 61 | 63 | 5 |
| **Publication Year, n (%)** |  |  |  |  |  |
| 2020 | 41 (10.6) | 31 (12.0) | 8 (13.1) | 1 (15.9) | 1 (20.0) |
| 2021 | 144 (37.1) | 102 (39.4) | 20 (32.8) | 19 (30.2) | 2 (40.0) |
| 2022 | 203 (52.3) | 126 (48.6) | 33 (54.1) | 43 (68.3) | 2 (40.0) |
| **Study Designs, n (%)** |  |  |  |  |  |
| Cross-sectional | 189 (48.7) | 155 (59.8) | 18 (29.5) | 14 (22.2) | 2 (40.0) |
| Quasi/Pre-post | 41 (10.6) | 14 (5.4) | 15 (24.6) | 12 (19.0) | 0 (0) |
| Cohort | 37 (10.8) | 20 (7.7) | 8 (13.1) | 8 (12.7) | 1 (20.0) |
| Qualitative | 40 (9.5) | 28 (10.8) | 6 (9.8) | 6 (9.5) | 0 (0) |
| RCT | 7 (1.8) | 0 (0) | 3 (4.9) | 4 (6.3) | 0 (0) |
| Economic evaluation | 1 (0.03) | 0 (0) | 0 (0) | 1 (1.6) | 0 (0) |
| Ecological/time-series analyses | 14 (3.6) | 13 (5.0) | 0 (0) | 1 (1.6) | 0 (0) |
| Mixed methods | 22 (5.4) | 13 (5.0) | 2 (3.3) | 7 (11.1) | 0 (0) |
| Review (scoping/systematic) | 27 (5.7) | 14 (5.4) | 5 (8.2) | 6 (9.5) | 2 (40.0) |
| Other (audit, country profile, policy analyses) | 10 (2.6) | 2 (0.8) | 4 (6.6) | 4 (6.3) | 0 (0.0) |
| **Language, n (%)** |  |  |  |  |  |
| English only | 228 (58.8) | 126 (48.6) | 36 (59.0) | 63 (100) | 4 (80.0) |
| Spanish/Portuguese<sup>#</sup> | 160 (41.2) | 133 (51.4) | 25 (41.0) | 0 (0) | 1 (20.0) |
| LAC = Latin America and the Caribbean |  |  |  |  |  |

\*Both LAC and Non-LAC #With or without English translations
<sup>¶</sup>Disruption to the delivery of primary care services, with a particular focus on diabetes and hypertension care (awareness, detection, treatment and control), and recovery of those services where interruptions took place
<sup>§</sup>Interventions to recover potential disruptions in diabetes or hypertension care, including other primary care services in LAC (or other regions) during or after the pandemic

### 3.1 Overview of studies on care disruption in LAC

Of 259 studies, 126 were from Brazil, 22 from Argentina, 22 from Mexico, and 19 from Peru (**appendix 1 pp 25**). Where indicated, most studies described disruptions to primary care services in urban (n=92) or mixed urban and rural settings (n=69) and in public (n=94) or mixed public and private healthcare settings (n=69). Only five studies explicitly studied data in rural areas and only eight in private healthcare settings. Cross-sectional studies (n=155) constituted the most common design, followed by qualitative (n=28) and cohort studies (n=20). Diabetes was explored in 41 studies (16%), and hypertension in 26 studies (10%) (**Table 2, appendix 1 pp 26**).

Each sub-domain of the PHCPI Framework was represented in at least 20 papers (**Figure 3, appendix 1 pp. 27-36**). In the System or Inputs domains, workforce was studied most (n=138), followed by facility infrastructure (n=104), governance and leadership (n=68), and drugs and supplies (n=47). Within the Service Delivery domain, access to primary care services and availability of services were most frequently studied (n=83), followed by patient perceptions of seeking and receiving primary healthcare and facility organization (n=51). In the Outputs domain, key process measures were studied most (n=121), followed by hypertension or diabetes treatment and control (n=57). Within the Outcomes domain, resilience was addressed most (n=148), followed by responsiveness to people (n=69). However, the coverage of domains and subdomains in **Figure 3** does not necessarily reflect the level of evidence for each due to variations in quality and objectivity of data (**Figures e2-e6, Tables e5 & e7**).

**Figure 3.**
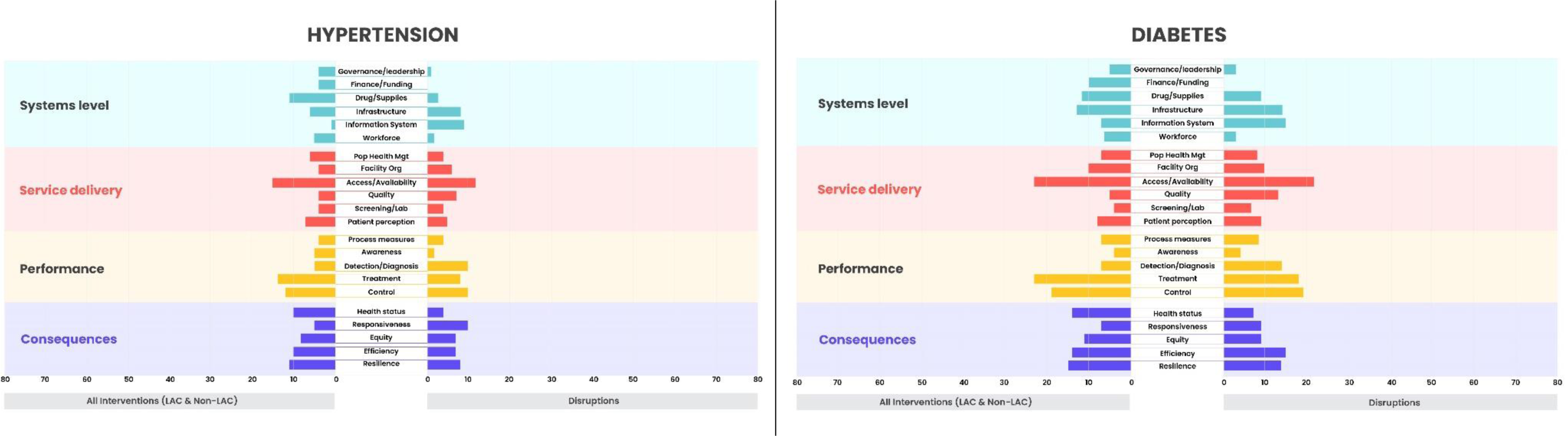
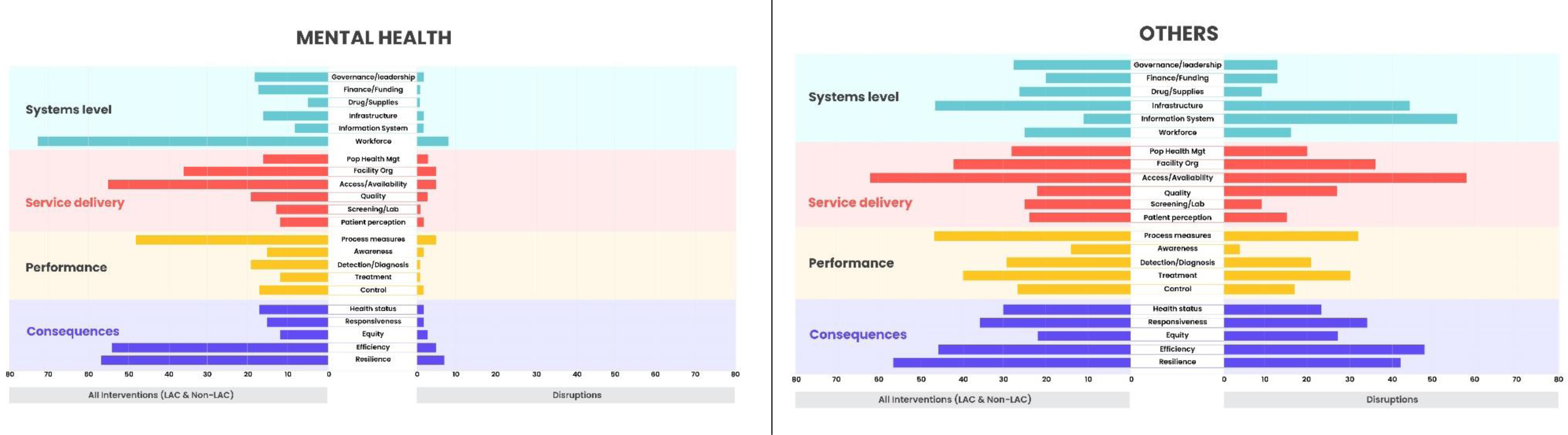
Domains of primary care services addressed in Latin America and the Caribbean (LAC) and Non-LAC countries, by disease area.

#### 3.1.1 Magnitude of disruptions to diabetes and hypertension care

Few studies provided quantitative data on treatment or control rates of diabetes and hypertension compared to periods before the pandemic. A Chilean study examined national clinical data from sites implementing the World Health Organization HEARTS Technical Package^16^ and reported a 12% and a 14% reduction in treatment and control rates of hypertension, respectively, in 2021 vs. 2017-2019. Similarly, social security data representing more than half of Mexico’s population^17^ demonstrated a decrease in hypertension control rates from 68% to 55%. This same investigation showed a reduction in diabetes control rates, from 36% to 26% during the pandemic. Decreases in hypertension and diabetes screening, including its complications, were also reported.^18,19^ In Brazil, there was a decrease in guideline-recommended screening examinations among patients with type 1 diabetes from 23% pre-pandemic to under 4% in 2022.^20^ Decreasing clinic visits for diabetes and hypertension management were also reported in Chile, Haiti, and Mexico.^21^

#### 3.1.2 Major disrupted subdomains

##### 3.1.2.1 Workforce

###### Mental Health of Healthcare Workers

115 papers reported workforce-related mental health outcomes. Increases in burnout, anxiety, depression, and insomnia among doctors, nurses, and other healthcare staff were described in several countries.^22–27^ Administrative data from Brazil showed a 17% increase in psychiatric sick leave during the pandemic.^28^ Another Brazilian study noted a 39% increase in mental health absenteeism among 32,691 hospital workers.^29^ Nursing assistants and other frontline workers were disproportionately impacted compared to physicians and nurses.^30–32^ Contributing factors included lack of personal protective equipment (PPE), increased patient volume due to COVID-19, disruptions to workflow, social isolation, and perception of insufficient organizational and governmental support.^22,33–37^ A Colombian study reported that workers with an increased workload due to COVID-19 had ∼2 times higher odds of anxiety and depression compared to those without workload changes.^32^ Increased negative health behaviors, including smoking, alcohol use, and reduced physical activity, were also reported among healthcare workers.^38^

###### Changes in Work Environment

During the pandemic, changes to the work environment and conditions were reported.^39–42^ In Brazil, increases in physician income were noted at public facilities, whereas income was reduced in the private sector.^40^ A Brazilian study showed increases in job availabilities for registered nurses (14%), nurse assistants (9%), physiotherapists (8%), and physicians (5%).^41^ A Brazilian survey of 1,609 healthcare workers found that 65% of psychologists were working remotely, while only 20% of dentists were doing so.^42^ Moreover, while opportunities for early assumption of clinical responsibilities among young doctors in Brazil arose during the pandemic, challenges related to support and social integration of early career physicians were noted.^43^

##### 3.1.2.2 Medication and essential supplies

In a 2020 survey of 40 diabetes organizations in 18 LAC countries, approximately two-thirds of respondents reported shortages of diabetes medications and a lack of critical supplies, such as syringes/needles and glucose strips. Only 37% of the respondents reported that a policy had been introduced to allow for the multi-month delivery of diabetes medications and supplies at home or dispensaries for two-to three months.^44^ In Peru, availability of essential medicines like losartan and metformin declined initially, but rebounded in primary care, exceeding pre-pandemic levels. Recovery appeared lower, however, in secondary and tertiary care facilities.^45^ Lack of PPE and other essential supplies were also reported in some studies.^30,40–42^

##### 3.1.2.3 Access and availability of relevant services

Several studies highlighted substantial reductions in preventive and screening services, routine check-ups, and outpatient care in general.^50–54^ For example, a Brazilian study reported a 77% drop in pap smears and a 43% decrease in mammograms in 2020-2021 compared to pre-pandemic levels. Significant declines in essential health services were reported in Mexico, including screenings (26-96% decrease), outpatient visits (9-40% decrease), and maternal health appointments (5-33% decrease).^17^ Outpatient services and elective procedures were also impacted, with decreases ranging from 46-75% across different studies.^55–57^ Many facilities focused resources on pandemic response, resulting in barriers like long wait times for primary care.^51,58^ While several studies mentioned reductions in screening services, none specifically reported on screening for new cases of hypertension or diabetes.

##### 3.1.2.4 Governance and leadership

The pandemic revealed gaps in governance and leadership across multiple levels of health systems in LAC.^59–61^ Studies emphasized challenges in coordinating evidence-based responses, managing scarce resources, and confronting healthcare inequities exacerbated by the pandemic.^62–66^ Significant regional disparities in health workforce availability were also reported.^67^ Delays and inconsistencies in healthcare guidelines and policies were reported in Mexico during the initial pandemic stages.^68^ One study reported that Brazilian municipalities perceived the municipal pandemic management as inadequate across various facets, including infrastructure, supplies, and communication.^58^

##### 3.1.2.5 Patient factors

Studies identified patient-level barriers to disease management, including worsened financial situations, fear of COVID-19 infection in clinics, and reduced adherence to medications and lifestyle changes.^58,70,71^ A Mexican study found that over half of patients with self-reported health needs did not seek care because they considered their issue not severe; other reasons were costs and fear of COVID-19 infection.^18^ Lifestyle changes also impacted disease management. A study in Brazil found that during the pandemic, eating habits worsened among adults with type 1 diabetes.^72^

### 3.2 Overview of intervention studies

Of 129 studies addressing interventions, 61 were carried out in LAC (**Table 2)**, predominantly in Brazil (n=28), Mexico (n=11), Argentina (n=10), and Colombia (n=8) (**appendix 1 pp 37-53, Figure e11**). Three studies included several LAC countries, and five included data from both LAC and non-LAC regions. Outside LAC, most interventions were from the United States (n=38) and European countries (n=9), and eight had multi-country representation.

In LAC, 30% of studies focused on diabetes (n=18) and 16% on hypertension (n=10) (**Figure 3**). Other major investigated conditions included mental health (n=12, 20%) and general medical encounters (n=14, 23%). The corresponding numbers in non-LAC countries were as follows: diabetes (n=13, 21%), hypertension (n=6, 10%), mental health (n=9, 14%), and general medical encounters (n=23, 37%).

Two main types of interventions were implemented both in and outside LAC: remote care strategies, generally termed “telehealth” or “telemedicine” (LAC: n=43, 71%; non-LAC countries: n=52, 83%, **appendix 1 pp 55-62**), and mental healthcare (LAC: n=12, 20%; non-LAC countries: n=8, 13%, **appendix 1 pp 63-64**). Other interventions involved drug access and prescriptions, behavioral interventions, policy, diet, funding, screening campaigns, and team-based care. Interventions targeted either patients or healthcare workers.

### 3.3 Remote care strategies within LAC

Of the 43 studies in LAC, half were cross-sectional descriptive studies (n=20); only three were randomized controlled trials (RCTs).^73–75^ Remote care was applied across numerous conditions, including 11 studies on diabetes and 5 on hypertension. Remote care was predominantly offered in tertiary care settings (n=27; primary care: n=7) delivered during real-time synchronous encounters (n=35). Technologies ranged from telephone calls (n=20) to smartphone applications and virtual meeting platforms such as WhatsApp (n=22), Zoom, and Microsoft Teams (n=13). Studies evaluated various aspects of remote care implementation: feasibility and utilization (n=31), healthcare provider or patient acceptance and satisfaction (n=14), and some level of intervention effectiveness (n=19).

#### 3.3.1 Feasibility and utilization

##### 3.3.1.1 Healthcare delivery and clinical education

Studies reported feasibility based on the number of teleconsultations completed. In Mexico, a primary-care diabetes clinic^76^ and a large hospital-based study^77^ perceived phone calls, text messaging, and video calls helpful for safe continuity of care and monitoring prescription needs among patients with diabetes or hypertension. In Brazil, telephone calls and text messages permitted monitoring of adherence among patients with type 1 diabetes.^78^ Additionally, WhatsApp messages, audio, and video calls improved patients’ access in remote communities.^68^ A cross-sectional analysis of an ongoing randomized trial in Guatemala^80^ reported on the success of telephone calls for verifying delivery of anti-hypertensive medication (e.g., 73% in the intervention group vs. 51% in the control group). Remote healthcare strategies were also used for patient education. In Colombia, web-based video conferencing was used to train patients with type 1 diabetes who were switching from regular insulin pumps to a hybrid closed-loop system.^83^ In Brazil, telephone calls by nurses providing education on hypertension treatment and COVID-19 guidance helped avoid unnecessary emergency room visits.^84^

##### 3.3.1.2 Triaging and clinical support

A Brazilian study concluded that teleophthalmology for patients with diabetes was a feasible low-cost strategy to increase diabetic retinopathy screening in remote areas.^86^ Similarly, in Peru, teleophthalmology was used to identify patients requiring face-to-face visits.^87^ Other examples included WhatsApp video calls to triage for respiratory symptoms,^88^ telephone consultations to triage primary care patients to specialty care,^89^ and a remote medical advice program triaging patients requiring in-person hospital evaluation.^77^ Another study from Mexico implemented teleconsultations to provide remote neurology support.^90^ Over 95% of the 304 teleconsultations conducted by neurology residents successfully addressed neurological issues without needing an in-person evaluation.

#### 3.3.2 Acceptance and satisfaction

##### 3.3.2.1 Acceptance and satisfaction - patients

In Argentina,^92^ patients in a cardiology clinic rated virtual visits highly (e.g., easy to carry out and not inferior to face-to-face visits). Similar findings were reported among primary care and hospital-based patients in Chile.^93^ In 56,560 web-based video consultations in Colombia,^94^ patients’ satisfaction was excellent at 84%. Acceptance also was high (>∼85%) among neurology patients^90^ and patients with diabetes.^76^ Some studies reported that patients accepted virtual visits because of the pandemic, but generally preferred in-person visits.^84,96^

##### 3.3.2.2 Acceptance and satisfaction – healthcare workers

A Brazilian study^97^ found that while only 19% of providers had used telemedicine pre-pandemic, 83% reported engaging in telemedicine training, and 64% used telemedicine to provide care during the pandemic. However, perceptions of telehealth among healthcare professionals were mixed; in Chile, one study^93^ reported that 62% of medical providers felt their clinical skills were challenged on video visits.

#### 3.3.3 Effectiveness

Of 19 studies aiming to evaluate effectiveness, most did not quantify disease outcomes or lacked control/comparator groups. Furthermore, some studies were small (e.g., <50 participants). None conducted a health economic assessment.

##### Diabetes

In a Colombian study using video conferences for monitoring and training 91 patients with type 1 diabetes, there were significant improvements in percent of time in glucose control range (77% to 82%) comparing pre- and post-intervention.^83^ An observational study in Brazil carried out telephone monitoring of 237 patients with type 2 diabetes^78^ and reported a small but statistically significant reduction in HbA1c (7.9% vs. 7.7%, p=0.004).

However, a study^100^ (n=143) that used telephone calls and text messages to send blood glucose values, check insulin doses, and periodically monitor for acute complications in patients with type 1 diabetes observed worsening of HbA1c in 46% of patients. A Brazilian center^101^ (n=747) assessed a telemedicine program without specific training and found hospitalization rates increased by ∼50%. In contrast, a pre-post study from the Dominican Republic^102^ used WhatsApp and telephone calls to monitor 946 dialysis patients and observed decreased hospitalization and a 2% COVID-19 diagnosis rate over a 3-month follow-up period.

##### Hypertension

A Chilean study used video calls from a mobile device to facilitate communication between the emergency department and a remote neurologist for rapid thrombolytic decision-making for stroke. Time from arrival to decision for thrombolytic administration was significantly reduced (median 5 [IQR 3–8] vs. 6 [IQR 4–10] minutes, p=0.016) compared to pre-implementation.^103^ A study from Guatemala used an integrated intervention approach, including patient health coaching and home blood pressure monitoring in rural communities, and reported fewer problems with antihypertensive medication availability.^80^

#### 3.3.4 Team-based care strategies in LAC

Two RCTs within LAC utilized multi-professional team-based approaches for delivering remote interventions. An RCT (n=91) in Brazil found that a 16-week multi-professional remote intervention, including weekly telephone calls and educational materials, significantly reduced the prevalence of positive screening for mental health disorders (37.0% vs. 57.8%, p=0.04) and diabetes-related emotional distress (21.7% vs. 42.2%, p=0.03) compared to standard care among adults with type 2 diabetes.^75^ Another RCT (n=47) evaluated the effects of a 16-week multi-professional remote intervention with a nurse compared to an in-person multi-professional intervention without a nurse on body composition parameters among obese adults in Brazil.^73^ The remote intervention group had significant decreases in waist circumference (p<0.001) and blood glucose (p=0.014) compared to in-person care.

#### 3.3.5 Remote care strategies outside the LAC region

We identified additional insights regarding intervention effectiveness in studies from countries outside LAC. For example, remote diabetes care delivered by primary care physicians in the USA achieved better medication adherence compared to non-physician providers (83% vs 77%).^104^ Additionally, HbA1c control rate of remote and in-person care was similar in urban settings (63% vs 65%), but lower in rural areas (56% vs 63%).^105–107^ Other studies indicated that rural areas face more barriers to engagement with virtual care options than urban areas.^108–110^ In contrast, some studies found that telemedicine increased access and utilization for rural residents and other underserved populations in Canada,^106^ Saudi Arabia,^91^ and other countries.^92^ Other barriers included insufficient technological literacy or skills and discomfort with remote monitoring devices.^111^ Furthermore, internet availability, costs, reimbursement considerations, privacy or data concerns, and practice workflow challenges emerged as common themes.^111^ One Australian study reported initial cost increases to the Medicare Benefits Schedule for general practitioner consultations after expansion of public telehealth coverage (pre-pandemic: $545 million per month; March 2020 – December 2021: $592 million per month; p=0.0001). After this initial increase, there was no significant change in costs.^105^

### 3.4 Mental health support for providers in LAC

In Uruguay,^98^ an online mindfulness program reported improved stress scores among 15 healthcare workers. A brief remote cognitive behavioral therapy intervention decreased anxiety and depression symptoms in 26 Mexican healthcare workers, though time constraints and workload impeded participation.^112^ In Brazil, ^99^ a program providing mental health support for 395 hospital-based healthcare workers showed that workers appreciated the support and connectedness from the virtual interventions. An RCT (n=120) in Brazil showed that oral cannabidiol (CBD) plus standard care vs standard care alone reduced emotional exhaustion over a month. CBD also reduced anxiety and depression symptoms, but five serious adverse events occurred in the CBD group.^74^

### 3.5 Mental health support for providers outside LAC

An RCT in Turkey showed reduced stress and work-related strain and improved psychological well-being among 52 nurses after mindfulness-based breathing and music therapy.^113^ Similarly, a mindfulness-based course lowered workforce distress, emotional exhaustion, fear of COVID-19, and physical stress symptoms, including sleep difficulties in Italy.^114^ In France, a psychological support hotline was rapidly implemented to provide support for hospital workers during the COVID-19 outbreak. After 26 days, the hotline had received 149 calls, helped assess symptoms, and referred staff to additional psychosocial support.^115^

### 3.6 Other interventions

#### 3.6.1 Interventions within LAC

A digital COVID-19 screening protocol among over 500 healthcare workers in Mexico promoted workforce wellness and NCD prevention and control by identifying previously undiagnosed hypertension, diabetes, and obesity.^57, 82^A 10-month medication home delivery program through partnerships with local pharmacies for over 800 older adults in Brazil resulted in 25% fewer in-person visits to the pharmacy. Among the program users, 67% and 21% received medications for hypertension and diabetes, respectively, and 92% of users considered the program satisfactory. ^116^

#### 3.6.2 Interventions outside LAC

In the USA, an analysis of electronic health records for 2,479 cardiometabolic patients showed that pharmacist-led medication review interventions (e.g., simplifying complex regimes) helped conserve supplies, reduce staff exposure, and maintain patient safety.^117^ In Portugal, a cross-sectional study of 603 patients found that community pharmacies taking on dispensing of typically hospital-only medications improved therapy adherence and lowered patient travel and absenteeism.^118^

In Saudi Arabia, a prospective interventional study delivered pharmacy-based education to 54 patients with diabetes, which improved disease knowledge, self-care, medication adherence, and diabetes control compared to usual care.^119^ In the USA, a community-based participatory approach providing online lifestyle education to 105 participants in underserved communities significantly improved health behaviors, self-reported health, and mental health scores.^120^ Integration of diabetes screening into routine COVID-19 testing facilities for 923 low-income individuals in a cross-sectional study identified high rates of previously undiagnosed diabetes and connected patients to follow-up care.^121^ Similarly, an RCT using non-clinical patient coaches increased cancer screening rates among 119 patients compared to usual care.

### 3.7 Recommended Actions

Based on the findings from this scoping review and feedback from the World Bank, PAHO, and our expert panel, we list potential actions relevant to LAC in **Table 3**. While their relevance will vary across LAC countries, they aim to contribute to strengthening data infrastructure to evaluate care disruptions in real-time and improve the resilience and equity of healthcare systems.

**Table 3.** Recommended actions based on review findings.

| Domain | Potential Actions |
| --- | --- |
| **System**<br>(Sub-domains: Governance & Leadership, Healthcare Financing, Population Health Needs, Surveillance, Innovation & Learning) | • Invest in telehealth infrastructure and equitable, virtual health outreach<br>• Create “learning communities” to share best practices between countries, for example in relation to telehealth adoption and chronic disease management<br>• Invest in nationwide healthcare data systems and qualified personnel, to enable real-time analysis allowing for earlier detection of disruptions to non-communicable disease care<br>• Establish standardized measures and reporting for monitoring health indicators, including service utilization and health outcomes, and metric critical to detecting health inequities, to enable benchmarking, comparisons across geographies and time, economic analysis, and relevant and responsive systems changes<br>• Strengthen cross-regional coordination and collaboration on health system resilience planning through agencies like the Pan-American Health Organization |
| **Input**<br>(Sub-domains: Drugs & Supplies, Facility Infrastructure, Information Systems, Workforce, Funds) | • Implement mental health promotion initiatives, screening programs, counseling and forums to support workforce resiliency<br>• Institutionalize evidence-based self-care skills training to aid coping mechanisms among healthcare workers<br>• Integrate team-based care strategies into care models (e.g. involving pharmacists, community health workers, lay health workers)<br>• Create a roster of retired healthcare workers for emergency situation<br>• Develop policy specifying expanded roles of healthcare workers in emergency situations, including those working in the private healthcare sector<br>• Diversify import sources and increase local production of drugs and medical supplies to avoid shortages during crises<br>• Enable cross-national collaborations on sourcing, stockpiling and distribution of essential medicines, e.g. antihypertensives and diabetes medications |
| **Service Delivery**<br>(Sub-domain: Population Health Management, Facility Organization & Management, Access, Availability of Effective Healthcare Services) | • Scale remote primary care strategies like telemedicine and related essential infrastructure to maintain continuity when in-person care is disrupted<br>• Expand reach of essential services into hard-to-access and rural communities through telehealth<br>• Leverage user-friendly, inclusive digital tools like telehealth consultations and mHealth applications to increase community awareness and equitable access to care<br>• Implement patient-centered treatment models tailored to community priorities and resources to improve engagement in self-care behaviors<br>• Harness digital health tools to increase access to medications and lifestyle change support needed to optimize treatment success |

The domains of Output and Outcomes are not included in this table since they are downstream of System, Input, and Service Delivery and responses are likely to aim at these three upstream domains.

## 4 Discussion

Our scoping review of 388 articles found some evidence that hypertension and diabetes care services in LAC were disrupted during the COVID-19 pandemic and indicated potential pathways that may explain these disruptions. Workforce (including the mental health of healthcare workers) and medication supplies were the most frequently reported disrupted in LAC. Disruptions to the healthcare environment were exacerbated at the patient level, resulting in adverse lifestyle changes, financial constraints, and fear of contracting COVID-19.

Our review also identified several promising mitigating interventions. Remote care strategies, including telemedicine (e.g., telephone calls, text messaging, video conferences), teleophthalmology, medication home delivery programs, and mental health care of healthcare workers were the two most studied interventions in LAC. Remote care strategies facilitated healthcare service delivery, clinical support of healthcare workers, and patient education, and they appeared feasible and acceptable. However, knowledge gaps remain in long-term benefits and cost-effectiveness of remote care in LAC. Findings reveal a breadth of innovations from several countries to mitigate pandemic disruptions, but rigorous evaluation is needed, particularly on sustainability and integration within health systems.

Given the number of studies identified in our scoping review, it was surprising that only a few evaluated changes in hypertension or diabetes control during the pandemic. This was likely a product of limited capacity for immediate assessment of relevant data in care delivery settings and the redirection of staff and system’s focus on the COVID response. Introducing data infrastructure and sufficient workforce capacity to allow for NCD treatment and control surveillance and data analysis in real-time could address these gaps.

Our review revealed substantial pandemic impacts on health workforces across LAC (e.g., burnout and mental distress), with disproportionate effects on some types of workers, e.g., worse for nurse assistants compared to physicians.^35^ It is natural for healthcare systems to focus on patient needs, but functional healthcare systems require a healthy workforce. Relevant also in non-LAC countries,^31^ this scoping review signaled several areas LAC countries could consider for strategies to improve the well-being of healthcare workers, such as monitoring provider distress, workload redistribution with task sharing policies, and increasing access to mental health resources.^35^ Dedicated efforts will also be required to build robust emergency response capacities, such as developing rosters of retired workers to be mobilized for emergencies and policies enabling extended task-shifting across provider cadres in emergencies.^124^

While medication shortages occurred initially in some settings, supplies rebounded quickly in others, contrasting with more prolonged global supply chain disruptions.^125^ These differences may be related to domestic pharmaceutical production capacity and supply chain resilience. Countries like Brazil reported pronounced insulin shortages,^128^ while Peru’s rebound aligned with policies fortifying local manufacturing after past shortages.^45^ Diversifying import sources, localized production and patient-centered delivery models can help countries prepare supply chains to withstand future crises. Cross-national regional collaborations on sourcing, stockpiling, and distribution may also be highly effective.

The pandemic introduced several unique circumstances leading to changes in patient behavior potentially detrimental to hypertension and diabetes management. For example, fear of COVID-19 infection posed a major barrier to care-seeking. Patients also faced intensified financial constraints and occupational disruptions (e.g., unemployment), that may have impacted care-seeking and self-management.^130^ Studies also revealed worsening mental health, diet, and health behaviors during lockdowns - all likely exacerbating existing illness. Remote care addressed some of these patient-level factors and indeed emerged as an adaptive strategy, as detailed below.

Most studies in LAC suggested that remote care strategies helped maintain continuity and access across a spectrum of clinical care needs. More specifically, telehealth enabled remote monitoring for clinical decision-making and helped reduce potential COVID-19 exposure. Patient acceptance was generally high, whereas provider acceptance was lower, highlighting the need for more telemedicine training in medical education. Although a few studies reported that telehealth performs comparably to in-person primary care,^76,101,131^ the evidence remains limited regarding its clinical effectiveness, cost-effectiveness, and long-term sustainability.^132,133^ Also, structural barriers around health literacy and digital skills (particularly among elderly patients), insufficient availability of technology, limited infrastructure, and suboptimal connectivity (particularly in rural and poorer regions) can be issues. There may also be cost-related barriers (e.g., out-of-pocket costs).^71,80,134^ These findings highlight the value of patient-centered telehealth in addressing these structural barriers.^110^ While global collaborations can accelerate learnings on how to optimize remote care strategies, optimal design (e.g., telehealth modalities like video conferences) of remote care and evaluation frameworks tailored to LAC contexts will be essential for scaling up remote care models in the region.^135^

### 4.1 Proposed modification to the conceptual framework

After completing this scoping review, we perceived screening and lab testing in the ‘service delivery’ domain as closely linked to the domain of “outputs,” along with other diagnostic tests (e.g., imaging). Regarding patient factors, our initial conceptual framework only specified patient perception, whereas other patient-related elements (e.g., patient financial situation) are relevant. Thus, we propose modifying our conceptual framework for future studies, as depicted in **Figure 4**.

**Figure 4.**
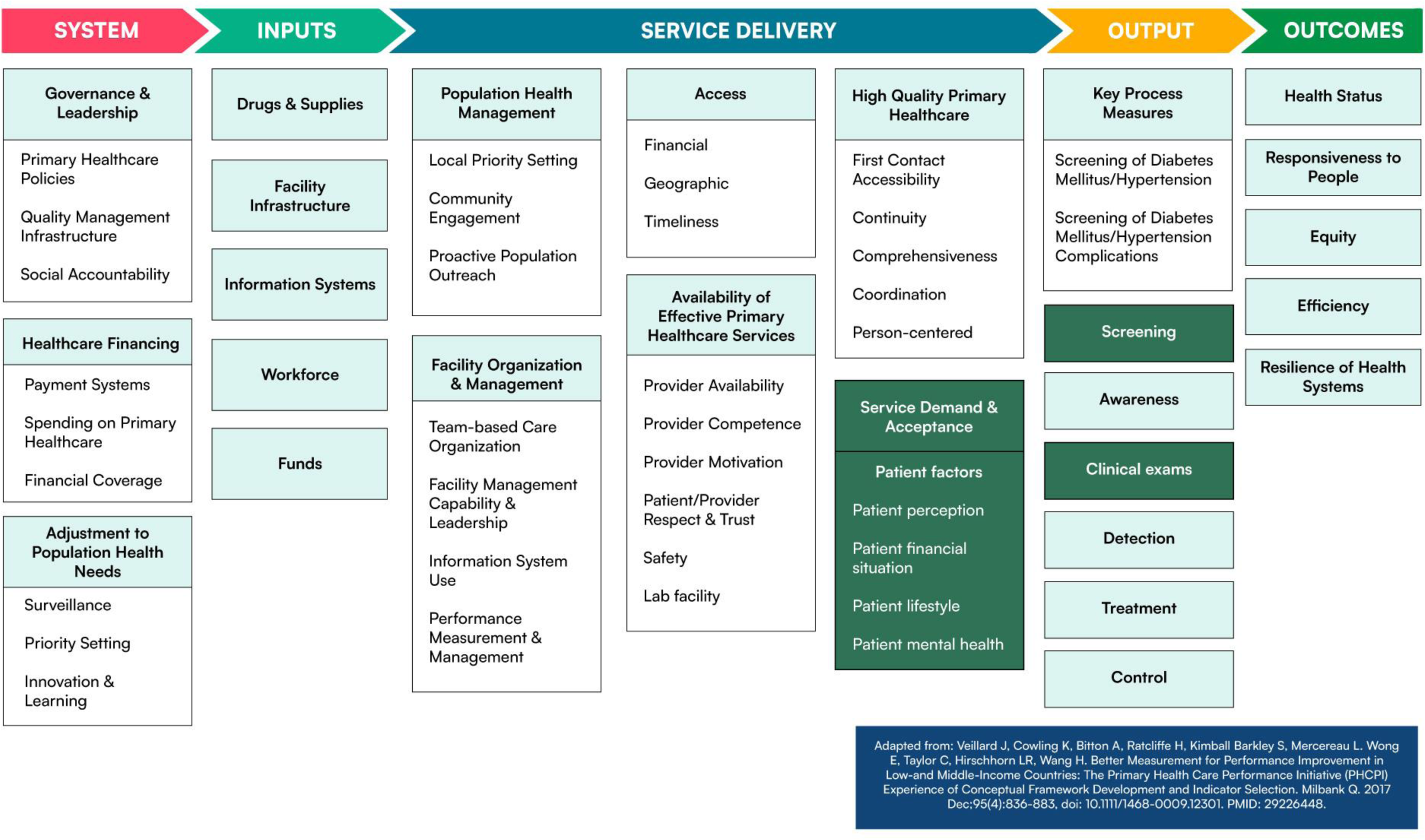
Second modification of the Primary Health Care Performance Initiative Conceptual Framework based on scoping review findings.

### 4.2 Limitations

Despite its comprehensiveness and rigor, the present scoping review has limitations. First, the limited data on interventions likely reflect the challenges of conducting rigorous research during the COVID-19 pandemic, especially in settings with limited research and data infrastructure, as well as the timing of our study (i.e., <3 years into the pandemic). Second, as noted above, grey literature was not systematically searched. Third, we did not consider the timing of included studies with the spread of COVID-19 across specific LAC countries. Finally, the review was restricted to studies published in English, Spanish, or Portuguese, while a few LAC countries have other languages as their primary language.

## 5 Conclusions

This review revealed concerning disruptions to essential NCD services, including diabetes and hypertension care across LAC. Workforce capacity and well-being, medication supplies, and patient perception were most reported as sources of the disruption. While this review identified promising strategies for adapting service delivery, predominantly remote care strategies, gaps remain in their cost-effectiveness and sustainability in the region. Our findings offer crucial insights that may inform health leaders in building resilient, equitable NCD care in LAC.

## Contributors

The scope of the review was conceived by the Pan American Health Organization and the World Bank. The concept sheet and study plan were developed by SYA, EWG, and KM, shared with PAHO and The World Bank, and revised as needed. OO, SBJ, RM, SA, EG, KM, LR, and DB iteratively developed the search strategy. Literature screening and data abstraction were conducted by OO, SBJ, RM, JM, DB, SH, ECB, FI, IAQ, KL, YL, YW, AR, HL, AN, AE, IN, JL, NM, IJ, TA, SN, MFFM, YYK, CS, JF, RBD, and TT; LA, SYA, EWG, KM provided input and oversight during the stages of the review process. Data adjudication and synthesis were undertaken by OO, SBJ, RM, JM, ECB, FL, AR, JL, NM, and IB, with input and iterative discussion with SYA, EWG, and KM. Tables, figures, and maps were developed by OO, SBJ, RM, JM, DB, SH, ECB, FI, IAQ, KL, AR, JL, NM, IJ, YYK, CS, JF, and RBD. OO, SBJ, RM, SYA, EWG, and KM drafted the first iteration of the manuscript and made substantial contributions to the critical review, editing, and revision. All authors approved the final version of the manuscript. All authors had full access to all the data in the study and had final responsibility for the decision to submit for publication.

## Data sharing

Researchers wishing to undertake additional analyses of the data or access original estimates are invited to contact the corresponding author (OO). The study protocol and analysis plan are available.

## Declaration of interests

We established a working consortium of investigators and policymakers from John Hopkins University, the Royal College of Surgeons in Ireland, the World Bank, the Pan American Health Organization, and experts from the LAC region. All other authors declare no competing interests.

## Data Availability

All data produced in the present work are contained in the manuscript.

## Acknowledgments

This project was supported by the World Bank. We appreciate the opportunity to contribute to this synthesis of published literature.

We thank individuals from the World Bank: Cristian A Herrera, Rialda Kovacevic, Francisca Corona Ravest, and Pan-America Health Organization (PAHO): Anselm Hennis, Silvana Luciani, Roberta Caixeta, who provided support in various ways. We thank our Regional Expert Committee members selected by PAHO, who assisted in locating grey literature and interpretation and contextualization of findings, providing both early and later input: Carlos A. Aguilar-Salinas (Mexico), Jaime Miranda (Peru although currently in Australia), Kenneth Connell (Barbados), Maria Inês Schmidt (Brazil).

## Notes

### Competing Interest Statement

The authors have declared no competing interest.

### Clinical Protocols

https://pubmed.ncbi.nlm.nih.gov/38262656/

