## Appendix 1 for "Disruption to diabetes and hypertension care during the COVID-19 pandemic in Latin America and the Caribbean and mitigation approaches: A Scoping Review"

**Table e1: Adapted PRISMA-ScR Checklist**^1^

| Section | Item | PRISMA-ScR Checklist Item | Page # reported on |
| --- | --- | --- | --- |
| **TITLE** |  |  |  |
| Title | 1 | Identify the report as a scoping review. | 1 |
| **ABSTRACT** |  |  |  |
| Structured Summary | 2 | Provide a structured summary that includes (as applicable) background, objectives, eligibility criteria, sources of evidence, charting methods, results, and conclusions that relate to the review questions and objectives. | 2 |
| **INTRODUCTION** |  |  |  |
| Rationale | 3 | Describe the rationale for the review in the context of what is already known. Explain why the review questions/objectives lend themselves to a scoping review approach. | 4 |
| Objectives | 4 | Provide an explicit statement of the questions and objectives being addressed with reference to their key elements (e.g., population or participants, concepts, and context) or other relevant key elements used to conceptualize the review questions and/or objectives. | 4 |
| **METHODS** |  |  |  |
| Protocol and registration | 5 | Indicate whether a review protocol exists; state if and where it can be accessed (e.g., a Web address); and if available, provide registration information, including the registration number. | 4 |
| Eligibility criteria | 6 | Specify characteristics of the sources of evidence used as eligibility criteria (e.g., years considered, language, and publication status), and provide a rationale. | 4, appendix p.6 |
| Information sources | 7 | Describe all information sources in the search (e.g., databases with dates of coverage and contact with authors to identify additional sources), as well as the date the most recent search was executed. | 4 |
| Search | 8 | Present the full electronic search strategy for at least 1 database, including any limits used, such that it could be repeated. | appendix p.4-5 |
| Selection of sources of evidence | 9 | State the process for selecting sources of evidence (i.e., screening and eligibility) included in the scoping review. | 4 |
| Data charting process | 10 | Describe the methods of charting data from the included sources of evidence (e.g., calibrated forms or forms that have been tested by the team before their use, and whether data charting was done independently or in duplicate) and any processes for obtaining and confirming data from investigators. | 4-5, appendix p.7-8 |
| Data items | 11 | List and define all variables for which data were sought and any assumptions and simplifications made. | appendix p.7-8 |
| Critical appraisal of individual sources of evidence | 12 | If done, provide a rationale for conducting a critical appraisal of included sources of evidence; describe the methods used and how this information was used in any data synthesis (if appropriate). | 4-5 |
| Summary measures | 13 | Not applicable for scoping reviews. | N/A |
| Synthesis of results | 14 | Describe the methods of handling and summarizing the data that were charted. | 4-5 |
| Risk of bias across studies | 15 | Not applicable for scoping reviews. | N/A |
| Additional analyses | 16 | Not applicable for scoping reviews. | N/A |
| **RESULTS** |  |  |  |
| Selection of sources of evidence | 17 | Give numbers of sources of evidence screened, assessed for eligibility, and included in the review, with reasons for exclusions at each stage, ideally using a flow diagram. | Figure 1 |
| Characteristics of sources of evidence | 18 | For each source of evidence, present characteristics for which data were charted and provide the citations. | Appendix p.14-23, appendix p.26-40 |
| Critical appraisal within sources of evidence | 19 | If done, present data on critical appraisal of included sources of evidence (see item 12). | appendix p.9-12 |
| Results of individual sources of evidence | 20 | For each included source of evidence, present the relevant data that were charted that relate to the review questions and objectives. | 8-19 |
| Synthesis of results | 21 | Summarize and/or present the charting results as they relate to the review questions and objectives. | Tables 2, appendix p.24-64 |
| Risk of bias across studies | 22 | Not applicable for scoping reviews. | N/A |
| Additional analyses | 23 | Not applicable for scoping reviews. | N/A |
| **DISCUSSION** |  |  |  |
| Summary of evidence | 24 | Summarize the main results (including an overview of concepts, themes, and types of evidence available), link to the review questions and objectives, and consider the relevance to key groups. | 5-11 |
| Limitations | 25 | Discuss the limitations of the scoping review process. | 14 |
| Conclusions | 26 | Provide a general interpretation of the results with respect to the review questions and objectives, as well as potential implications and/or next steps. | 12-14 |
| **FUNDING** |  |  |  |
| Funding | 27 | Describe sources of funding for the included sources of evidence, as well as sources of funding for the scoping review. Describe the role of the funders of the scoping review. | 15 |

**Figure e1. Primary Health Care Performance Initiative Conceptual Framework modified for this scoping review. The red box indicates subdomains modified for this project.**

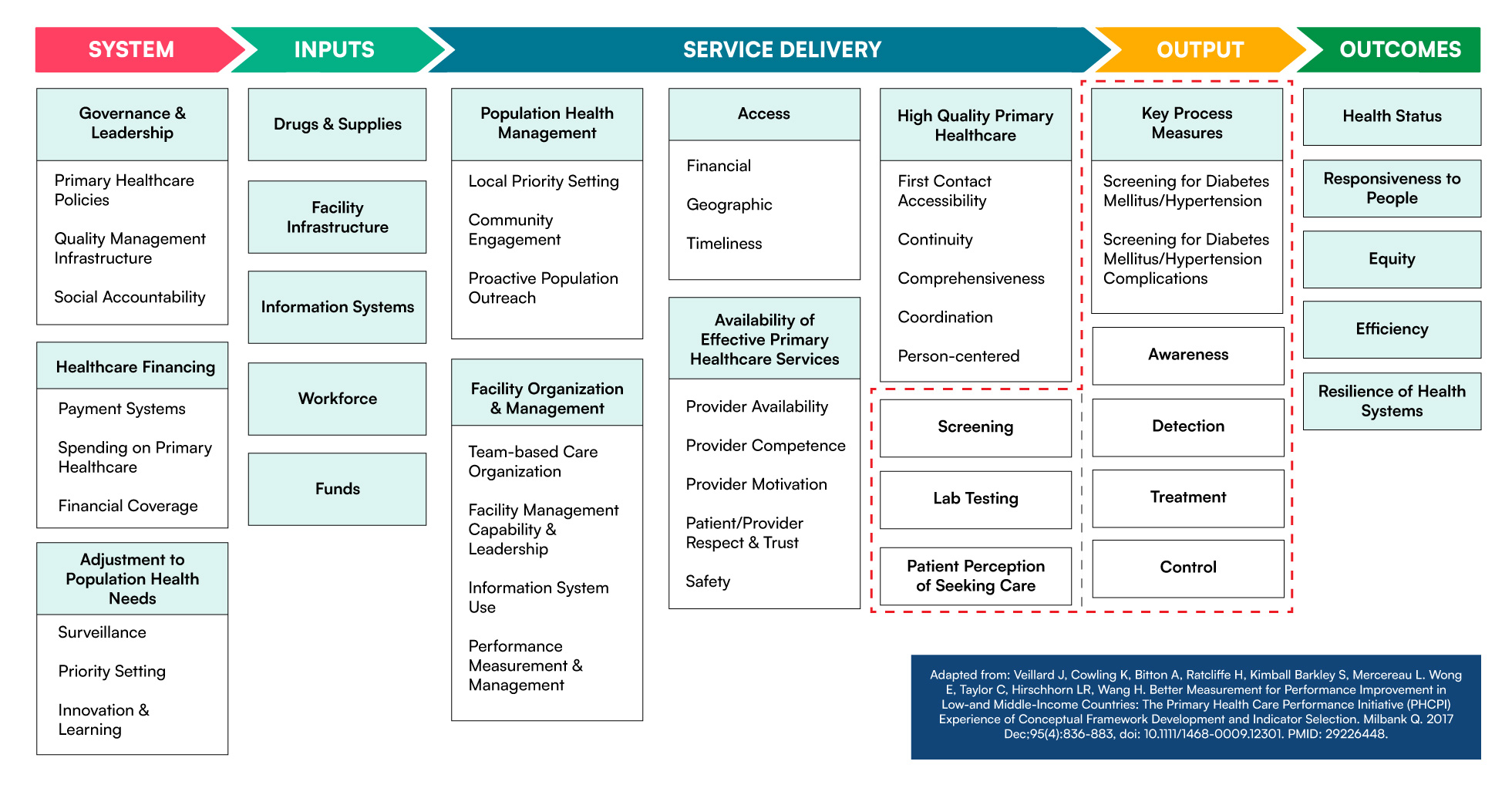

**Table e2: Population, Concept and Context (PCC) of this scoping review**

| **PCC item** | **Definition** | **Example** |
| --- | --- | --- |
| Population | Individuals for whom guidelines recommend screening or access to diabetes or hypertension care | Patients with type 2 diabetes |
| Concept | Disruption to the delivery of primary care services, with a particular focus on diabetes and hypertension care (awareness, detection, treatment and control), and recovery of those services where interruptions took place | Disruption: Cancellation of diabetes appointments due to perceived risk of contracting SARS-CoV-2 infection  Intervention - Recovery or mitigation: Offering virtual hypertension consultations to continue routine care |
| Context | Latin America and Caribbean region during the COVID-19 pandemic (2020-2022) | Colombia; Mexico City; Venezuelan border region |

**Table e3: Search strategy and search results from December 12, 2022**

| #1 | ("Coronavirus Infections"^2^ OR "COVID-19"^2^ OR "SARS-CoV-2"[Mesh] OR "Coronavirus"[Mesh] OR "Betacoronavirus"[Mesh] OR "COVID-19 Testing"[Mesh] OR "COVID-19 Vaccines"[Mesh] OR ("COVID-19"[tw] OR "COVID 19"[tw] OR "COVID19"[tw] OR "COVID2019"[tw] OR "COVID 2019"[tw] OR "COVID-2019"[tw] OR "novel coronavirus"[tw] OR "new coronavirus"[tw] OR "novel corona virus"[tw] OR "new corona virus"[tw] OR "SARS-CoV-2"[tw] OR "SARSCoV2"[tw] OR "SARS-CoV2"[tw] OR "2019nCoV"[tw] OR "2019-nCoV"[tw] OR "2019 coronavirus"[tw] OR "2019 corona virus"[tw] OR "coronavirus disease 2019"[tw] OR "corona virus disease 2019"[tw] OR "severe acute respiratory syndrome coronavirus 2"[tw] OR "sars-coronavirus-2"[tw] OR "SARS Coronavirus 2"[tw] OR "coronavirus disease 19"[tw] OR "corona virus disease 19"[tw]) OR (coronavirus[tw] OR "corona virus"[tw] OR coronavirinae[tw] OR coronaviridae[tw] OR betacoronavirus[tw] OR nCoV[tw] OR "CoV 2"[tw] OR CoV2[tw] OR sarscov2[tw] OR "novel CoV"[tw] OR "wuhan virus"[tw]) OR ((wuhan[tw] OR hubei[tw] OR huanan[tw]) AND ("severe acute respiratory"[tw] OR pneumonia[tw]) AND (outbreak[tw]))) |
| --- | --- |
| #2 | "Primary Health Care"[Mesh] OR "General Practice"[Mesh] OR "General Practitioners"[Mesh] OR "Health Services"[Mesh] OR "Primary Health Care"[Mesh] OR "Physicians, Primary Care"[Mesh] OR "Ambulatory Care"[Mesh] OR "Continuity of Patient Care"[Mesh] OR "Health Policy"[Mesh] OR "Delivery of Health Care"[Mesh] OR "first level of care"[tiab] OR "primary care"[tiab] OR "primary health care"[tiab] OR "general practice"[tiab] OR "general practitioner*"[tiab] OR "health service*"[tiab] OR "ambulatory care"[tiab] OR "continuity of patient care"[tiab] OR "continuity of care"[tiab] OR "healthcare system*"[tiab] OR "health care system*"[tiab] OR "health policy"[tiab] OR "health policies"[tiab] OR "healthcare policy"[tiab] OR "healthcare policies"[tiab]  OR  "Disease Management"[Mesh] OR "Self-Management"[Mesh] OR "Self Care"[Mesh] OR "disease management"[tiab] OR "self-manag*"[tiab] OR "self-care"[tiab] OR "care, self"[tiab] OR "self-treat*"[tiab] OR "self-test*"[tiab] OR "self-medicat*"[tiab] OR "self-monitor*"[tiab] OR "self-diagnos*"[tiab] OR "self-efficacy"[tiab] OR "self-competen*"[tiab]  OR  "Quality of Health Care"[Mesh] OR "Patient Acceptance of Health Care"[Mesh] OR "Patient Care Management"[Mesh] OR "Universal Health Care"[Mesh] OR "Facilities and Services Utilization"[Mesh] OR "Management Information Systems"[Mesh] OR "Quality of Health Care"[tiab] OR "healthcare quality"[tiab] OR "health care quality"[tiab] OR "Patient Acceptance of Health Care"[tiab] OR "Patient Acceptance of Healthcare"[tiab] OR "healthcare utilization"[tiab] OR "health care utilization"[tiab] OR "patient compliance"[tiab] OR "Facilities Utilization"[tiab] OR "Services Utilization"[tiab] OR "Patient Care"[tiab] OR "point of care"[tiab] OR "Universal Health Care"[tiab] OR "Universal Healthcare"[tiab] OR "Information Systems"[tiab]  OR  "Health Facilities"[Mesh] OR "Health Personnel"[Mesh] OR "Health Workforce"[Mesh] OR "Health Information Systems"[Mesh] OR "Budgets"[Mesh] OR "Financial Management, Hospital"[Mesh] OR "health facility"[tiab] OR "health facilities"[tiab] OR "health personnel"[tiab] OR "health care provider*"[tiab] OR "healthcare provider*"[tiab] OR "health care worker*"[tiab] OR "healthcare worker*"[tiab] OR "health care professional*"[tiab] OR "healthcare professional*"[tiab] OR "health workforce"[tiab] OR nurse*[tiab] OR pharmacist*[tiab] OR physician*[tiab] OR medication[tiab] OR "drug supply"[tiab] OR "drug supplies"[tiab] OR health facilit*[tiab] OR medical facilit*[tiab] OR "facility infrastructure"[tiab] OR outpatient*[tiab] OR budget*[tiab] OR financial*[tiab]  OR  "resilient health system*"[tiab] OR "sustainable health system*"[tiab] OR "health system sustainability"[tiab] OR "health system resilience"[tiab] OR "resilient health systems"[tiab] OR "patient centered care" [tiab] OR "patient acceptance"[tiab] |
| #3 | "Glucose Metabolism Disorders"[Mesh] OR "Glycogen Storage Disease"[Mesh] OR "Insulin Resistance"[Mesh] OR "Blood Glucose"[Mesh] OR "Glucose Tolerance Test"[Mesh] OR diabetes[tiab] OR diabetic*[tiab] OR "glucose intoler*"[tiab] OR "glucose toler*"[tiab] OR "blood glucose"[tiab] OR "blood sugar*"[tiab] OR "elevated serum glucose level*"[tiab] OR hyperglycemia*[tiab] OR hyperglucemia*[tiab] OR hyperglycaemia*[tiab] OR IDDM[tiab] OR MODY[tiab] OR NIDDM [tiab] OR T1DM[tiab] OR T2DM[tiab] OR prediabet*[tiab] OR IIDM[tiab] OR "glycogen storage disease*"[tiab] OR glycogenoses[tiab] OR glycogenosis[tiab] OR "glycosylation disorder*"[tiab] OR "insulin coma*"[tiab] OR "insulin resistan*"[tiab] OR "insulin sensitivit*"[tiab] OR "insulin shock*"[tiab] OR "insulin independ*"[tiab] |
| #4 | "hypertension"[Mesh] OR "Prehypertension"[Mesh] OR "Blood Pressure"[Mesh] OR "Noncommunicable Diseases"[Mesh] OR "Antihypertensive Agents"[Mesh] OR "hypertension*"[tiab] OR "hyper tension*"[tiab] OR hypertensive*[tiab] OR "blood pressure*"[tiab] OR prehypertens*[tiab] OR systolic[tiab] OR systole[tiab] OR diastolic[tiab] OR diastole[tiab] OR "pulse pressure*"[tiab] OR "Noncommunicable Diseases"[tiab] OR "Non communicable Diseases"[tiab] OR "Antihypertensive*"[tiab] |
| #5 | ("Latin America"[Mesh] OR "Latin America*"[tw] OR "Caribbean Region"[Mesh] OR Caribbean*[tw] OR Aruba*[tw] OR Curacao*[tw] OR "Sint Maarten*"[tw] OR "West Indies*"[tw] OR Montserrat*[tw] OR Turks*[tw] OR Caicos*[tw] OR Cayman*[tw] OR Antigua*[tw] OR Barbuda*[tw] OR Bahamas*[tw] OR Barbados*[tw] OR "British Virgin Island*"[tw] OR Cuba*[tw] OR Dominica*[tw] OR Grenada*[tw] OR Guadeloupe*[tw] OR Haiti*[tw] OR Jamaica*[tw] OR Martinique*[tw] OR "Puerto Rico*"[tw] OR "Puerto Rican*"[tw] OR "Saint Kitts*"[tw] OR "St Kitts*"[tw] OR Nevis*[tw] OR "Saint Lucia*"[tw] OR "St Lucia*"[tw] OR "Saint Vincent*"[tw] OR "St Vincent*"[tw] OR Grenadines*[tw] OR Trinidad*[tw] OR Tobago*[tw] OR "Virgin Island*"[tw] OR "Saint Bart*"[tw] OR "St Bart*"[tw] OR "Central America"[Mesh] OR "Central America*"[tw] OR Belize*[tw] OR "Costa Rica*"[tw] OR "El Salvador*"[tw] OR Salvadorean*[tw] OR Salvadoran*[tw] OR Salvadorian*[tw] OR Guatemala*[tw] OR Honduras*[tw] OR Nicaragua*[tw] OR Panama*[tw] OR "South America"[Mesh] OR "South America*"[tw] OR Argentina*[tw] OR Argentine*[tw] OR Argentinian*[tw] OR Bolivia*[tw] OR Brazil*[tw] OR Brasil*[tw] OR Chile*[tw] OR Colombia*[tw] OR Ecuador*[tw] OR Guyana*[tw] OR Guiana*[tw] OR Paraguay*[tw] OR Peru*[tw] OR Suriname*[tw] OR Surinam*[tw] OR Uruguay*[tw] OR Venezuela*[tw] OR "Mexico"[Mesh] OR Mexico*[tw] OR Mexican*[tw] OR "Mexican Americans"[Mesh] OR "Hispanic or Latino"[Mesh] OR Hispanic*[tw] OR Hispano*[tw] OR Latin[tw] OR Latina*[tw] OR Latine*[tw] OR Latinu*[tw] OR Latino*[tw] OR Latinx*[tw] OR "Spanish speak*"[tw] OR "Spanish America*"[tw] OR Boricua*[tw] OR Chicana*[tw] OR Chicano*[tw]) |
| #6 | #1 AND (#2 OR #3 OR #4) AND #5 AND 2020 – present |
| #7 | "Health Care Reform"[Mesh] OR "health system recovery"[tiab] OR "post-pandemic"[tiab] OR "post-pandemic recovery"[tiab] OR "post-COVID-19 recovery"[tiab] OR "recovery plan*"[tiab] OR "health strategy*"[tiab] OR "reform"[tiab] OR "disruption recovery"[tiab] |
| #8 | #1 AND (#2 OR #3 OR #4) AND #7 AND 2020 – present |
| #9 | #6 OR #8 Filters: from 2020 - 3000/12/12 |

**Table e4: Extracted outcome data items**

| **Item for data extraction** | **Description/Examples** |
| --- | --- |
| ***Study findings*** | |
| Continuity or disruption of diabetes care | Levels of diabetes care (i.e., diabetes awareness, detection, treatment and control, or management of complications) offered or provided in LAC during the pandemic. Examples include number of routine consultations, access to insulin, rate of blood glucose measurements, funding of diabetes services, etc. |
| Continuity or disruption of hypertension care | Levels of hypertension care (i.e., hypertension awareness, detection, treatment and control, or management of complications) offered or provided in LAC during the pandemic. Examples include fractions of individuals receiving blood pressure (BP) control or hypertension treatment, mean systolic BP/diastolic BP levels, number of anti-hypertensive medications dispensed, affordability of medication, etc. |
| Continuity or disruption of other primary care | Changes in provision of other primary care services in LAC during the pandemic |
| Diabetes/hypertension interventions | Interventions to recover potential disruptions in diabetes or hypertension care in LAC (or other regions) during or after the pandemic |
| Other primary care interventions | Interventions to recover potential disruptions in NCD care more generally in LAC during or after the pandemic |
| Domains of services that were disrupted | Drug and supplies; facility infrastructure; information systems; workforces; funds; community engagement; patients’ perceptions (e.g., fear of visiting clinics) |
| Care utilization or patient-related indicators (i.e., outcomes) | Frequency of clinic visits; adherence to treatment; diabetes and hypertension management cascade (frequency or rate of screening, detection/diagnosis, treatment, and control); mean BP levels; hypertension control; glycemic control |
| Type of interventions | Implementing/expanding telemedicine; promoting task-sharing; introducing longer prescription periods; funding |
| ***General study information*** | |
| Title | Study title |
| Author(s) | Study author(s) |
| Country/countries | Country or countries where study’s data were collected |
| Region(s) | Detail whether study is nationally representative, or otherwise detail region(s) of data collection |
| City | City or cities of data collection, if not nationally or regionally representative |
| Study objective | Briefly mention study objective, e.g. “to describe use of hypertension care services during the pandemic” |
| ***Methodological details*** | |
| Study design | Briefly mention study design and details of data collection methods, for example pre-post data collection for intervention studies |
| Setting and sector of healthcare system | (public/private/both/other) |
| Population | (urban/rural/both/other) |
| Clinical conditions | Diabetes; hypertension; other - mental health; other - chronic respiratory conditions (excluding COVID-19); other - other cardiometabolic conditions (e.g., obesity); other - liver disease; other - cancer screening (excluding established cancer care); other - maternal health |
| Data collection dates | Dates when study collected data |
| Participant description (if applicable) | Healthcare workers; patients |

| **Figure e2: Risk of bias assessment and summary for qualitative studies** | |
| --- | --- |
| 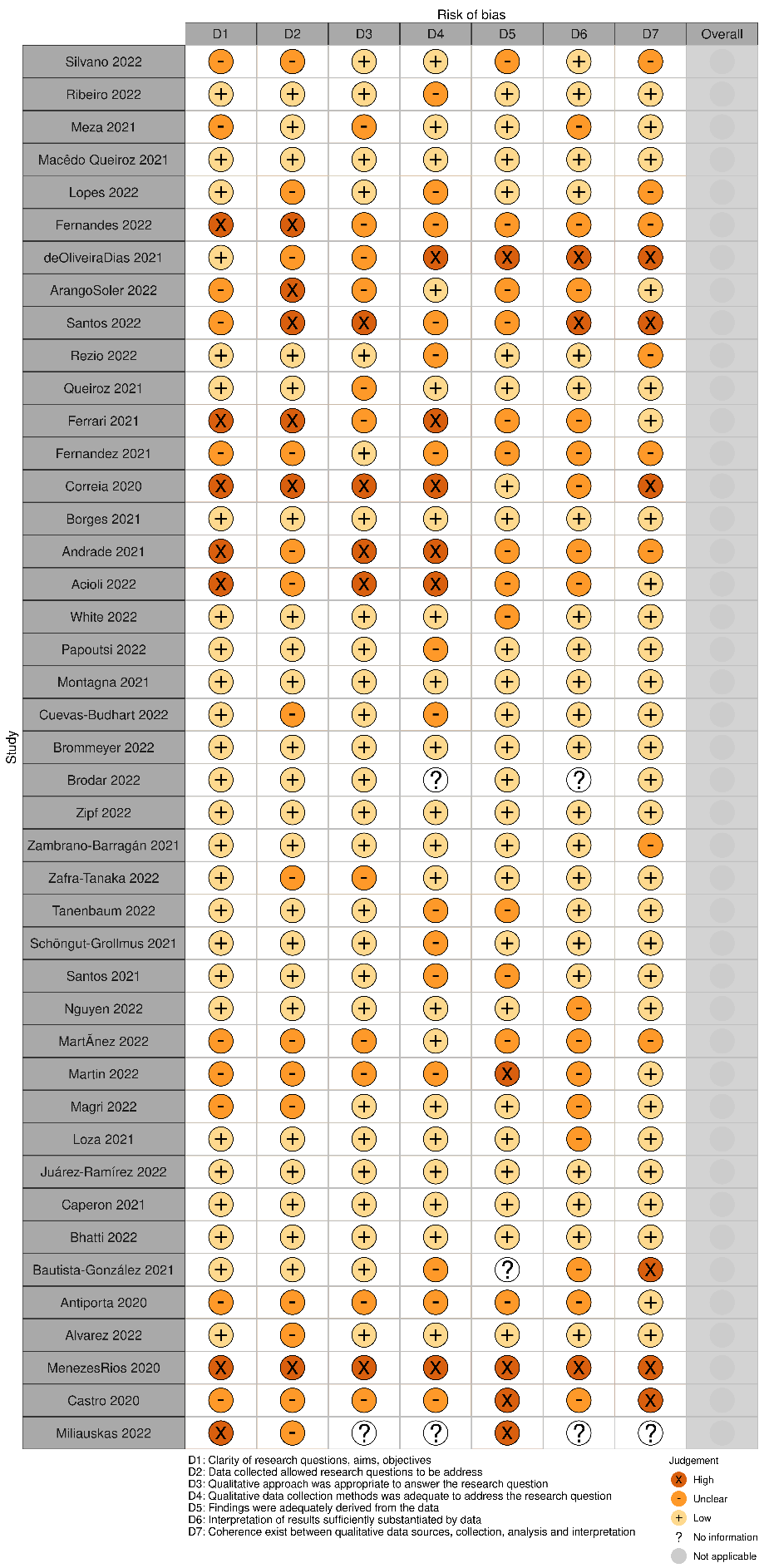 | 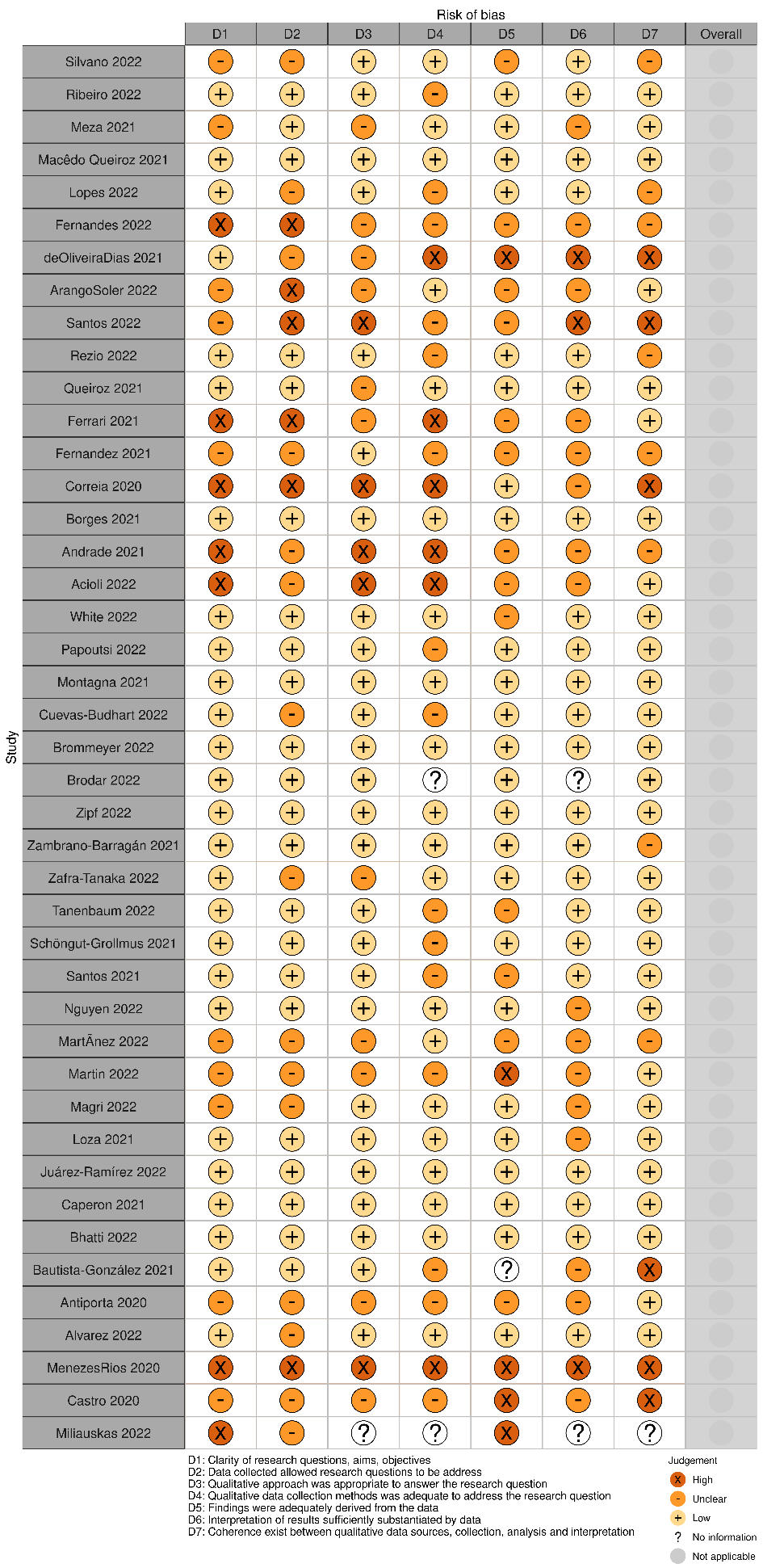 |

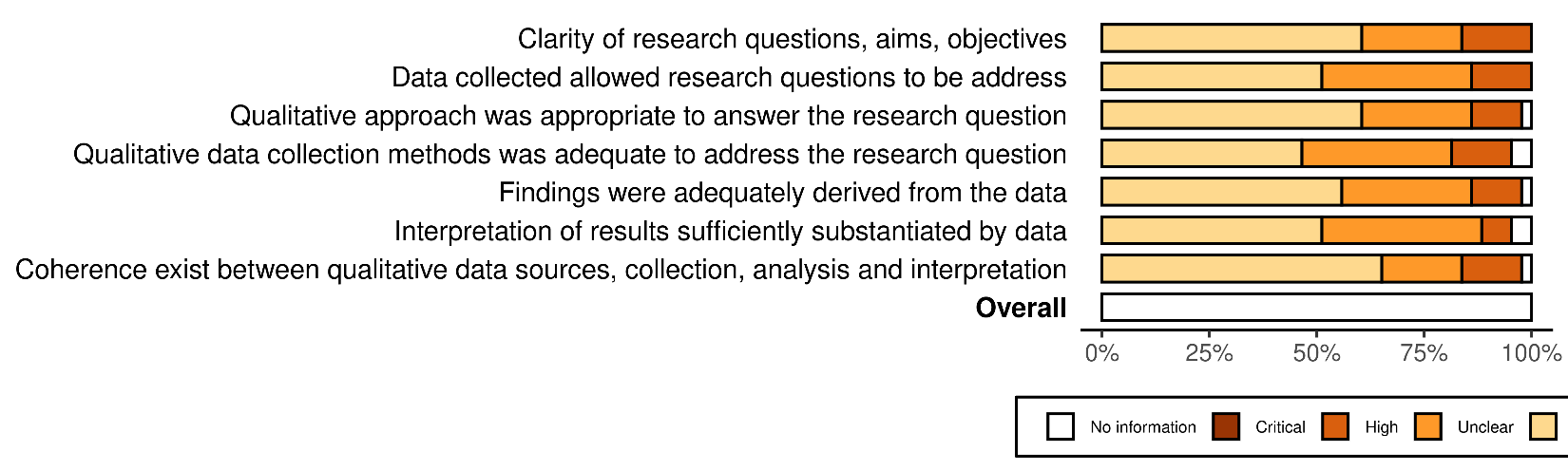

**Figure e3: Risk of bias assessment and summary for quantitative randomized studies**
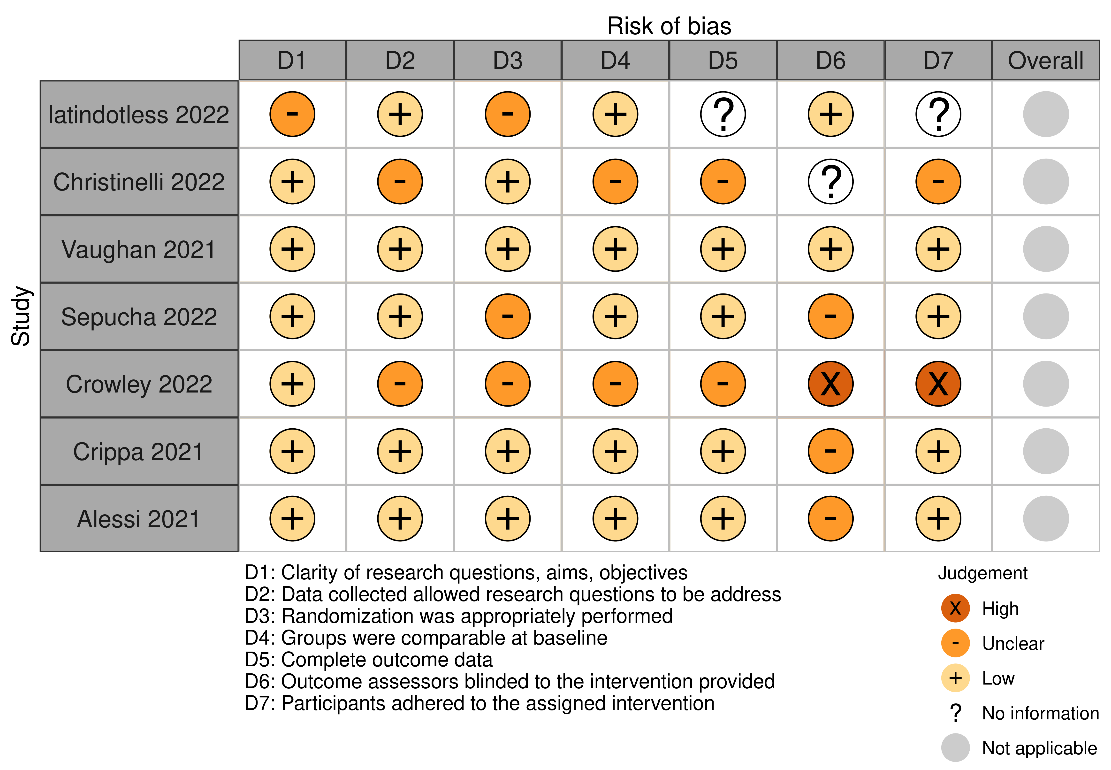

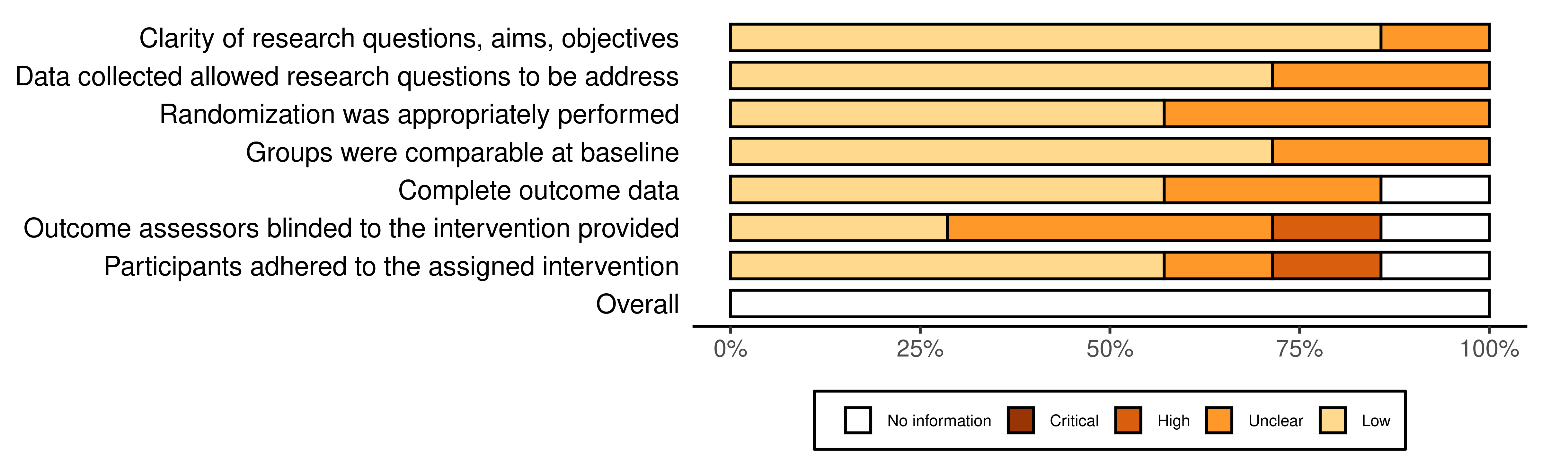

| **Figure e4: Risk of bias assessment summary for quantitative non-randomized studies (e.g. quasi, pre-post, etc.)** | |
| --- | --- |
| 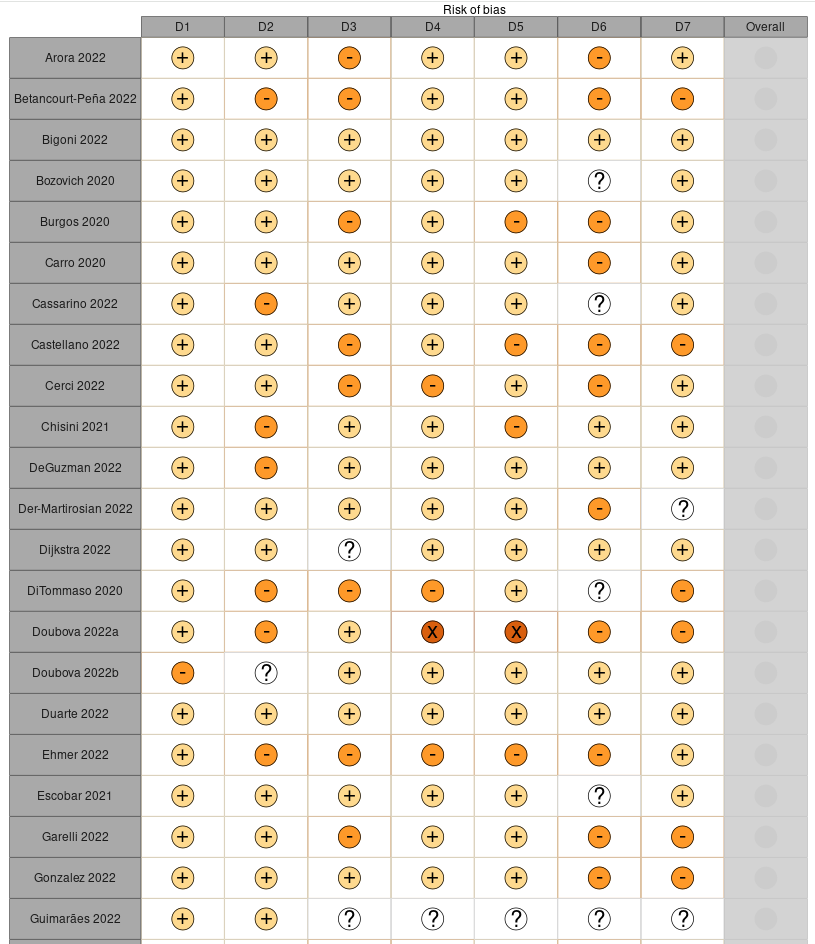 | 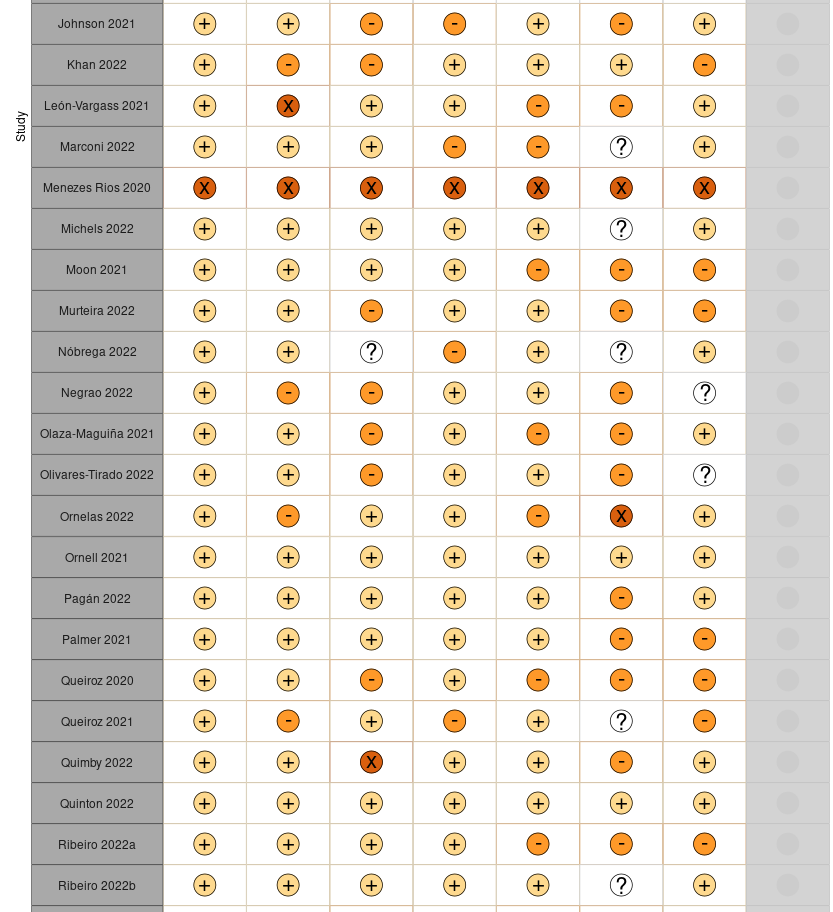 |
| 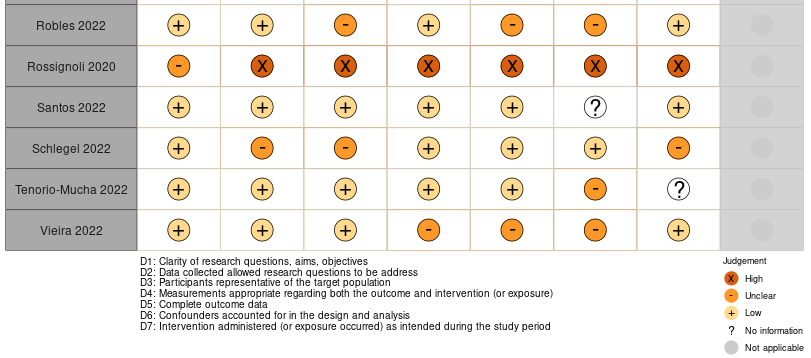 | |

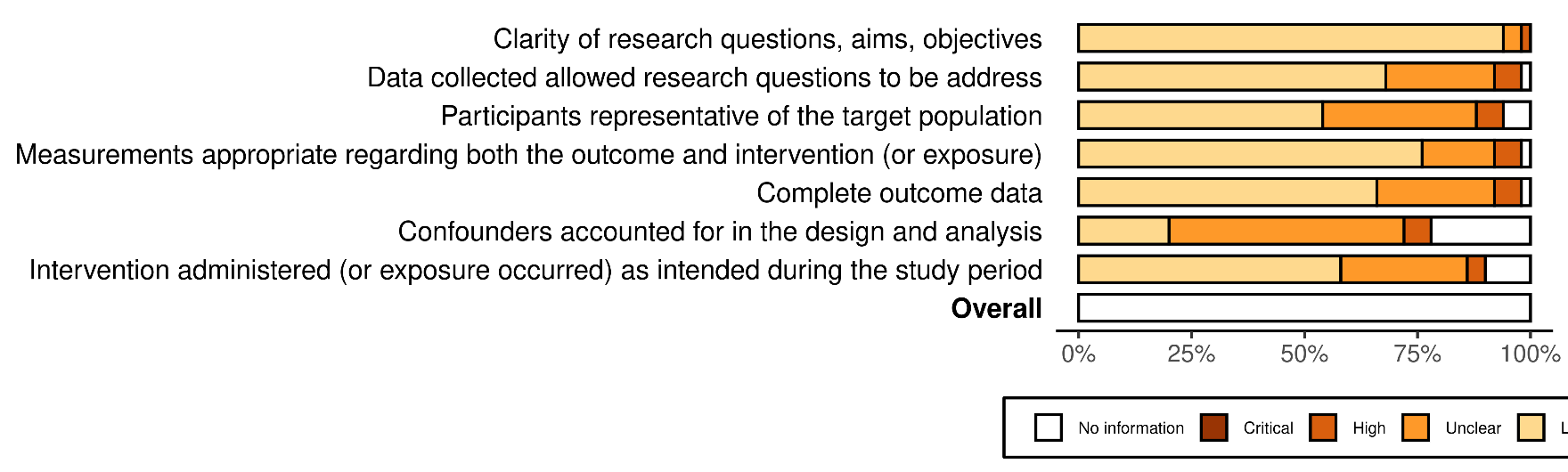

| **Figure e5: Risk of bias assessment and summary for quantitative descriptive studies (e.g., cross-sectional, etc.)** | |
| --- | --- |
| 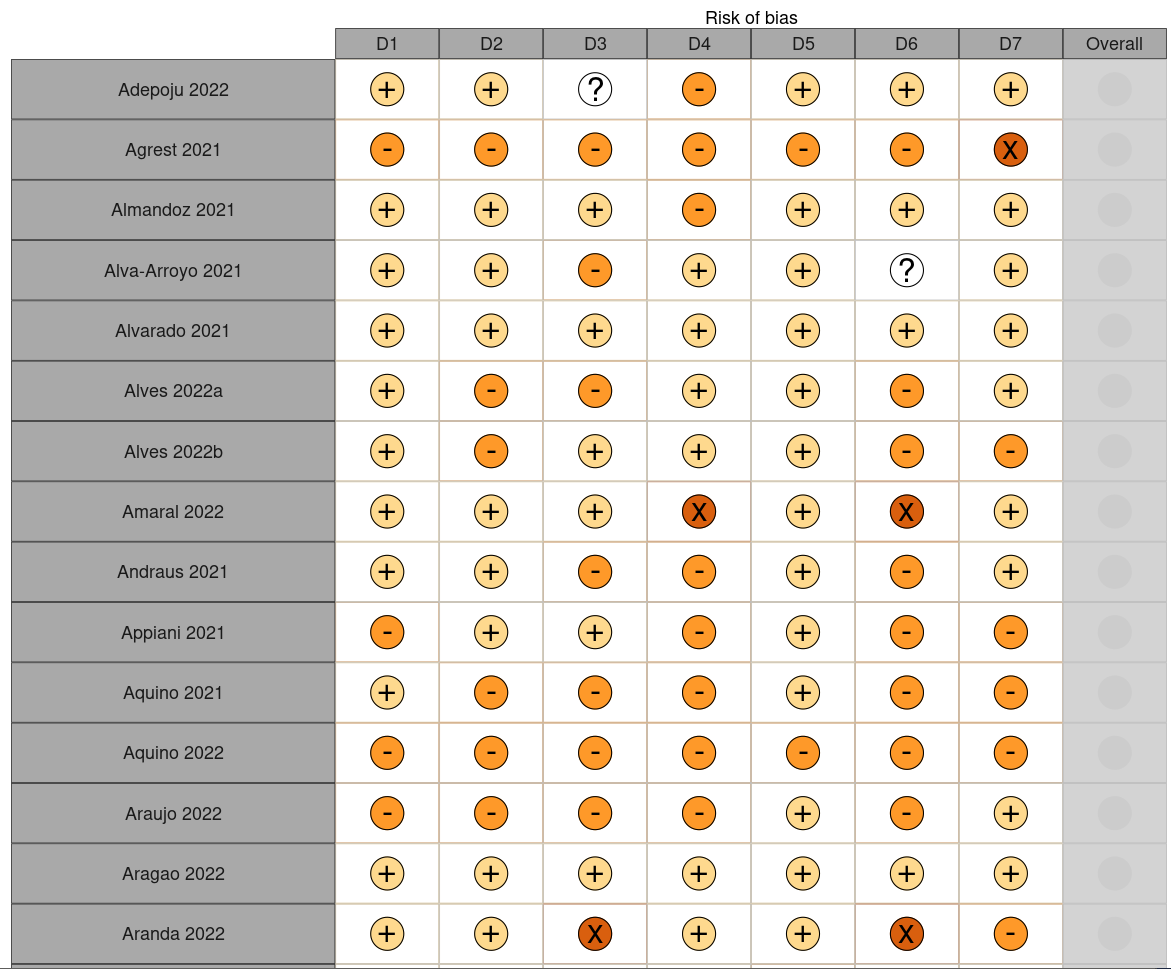 | 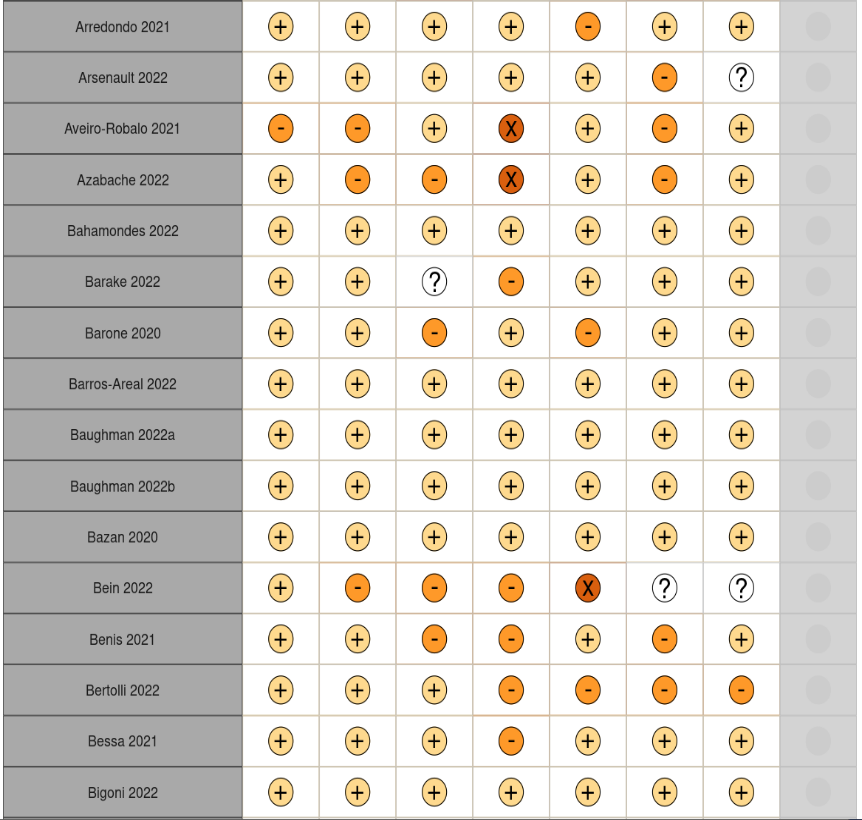 |
| 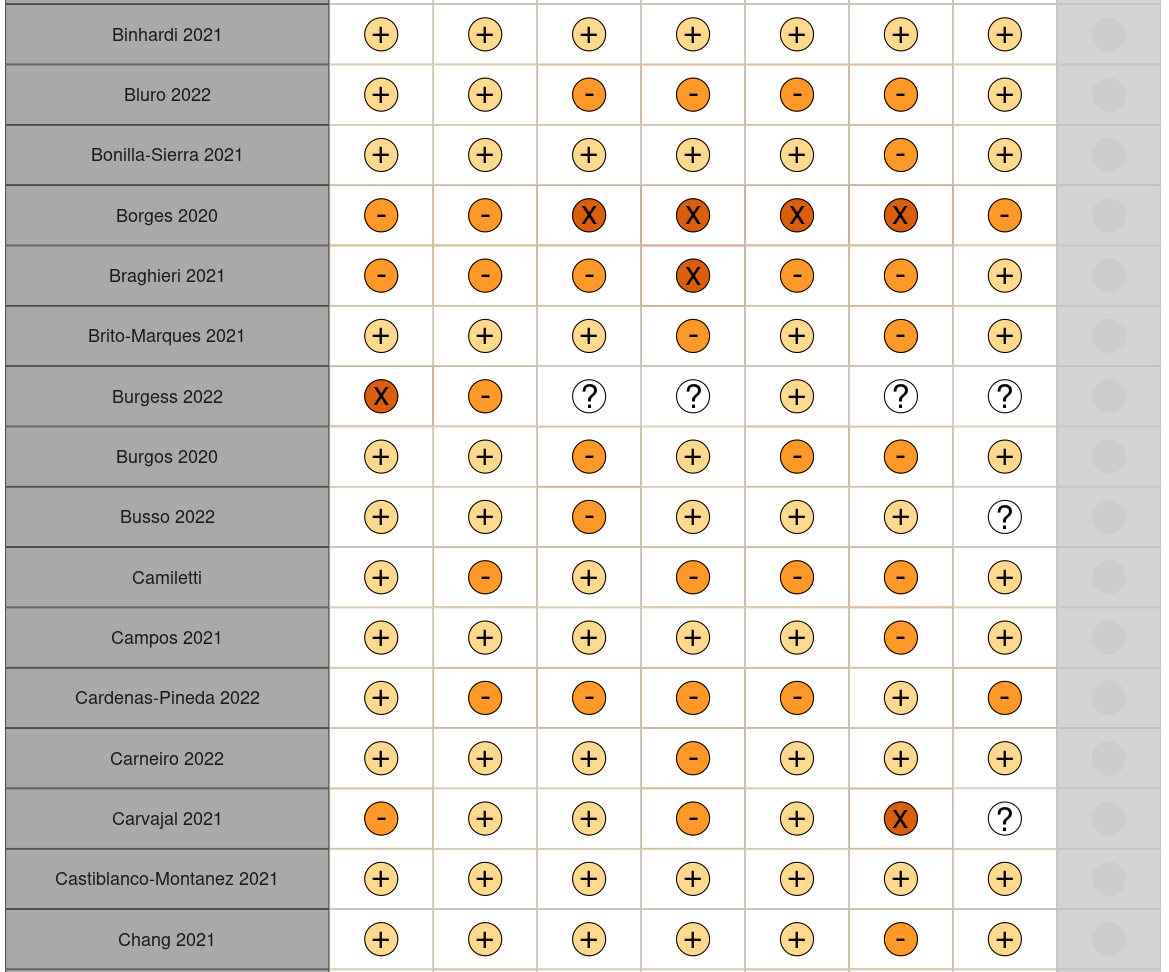 | 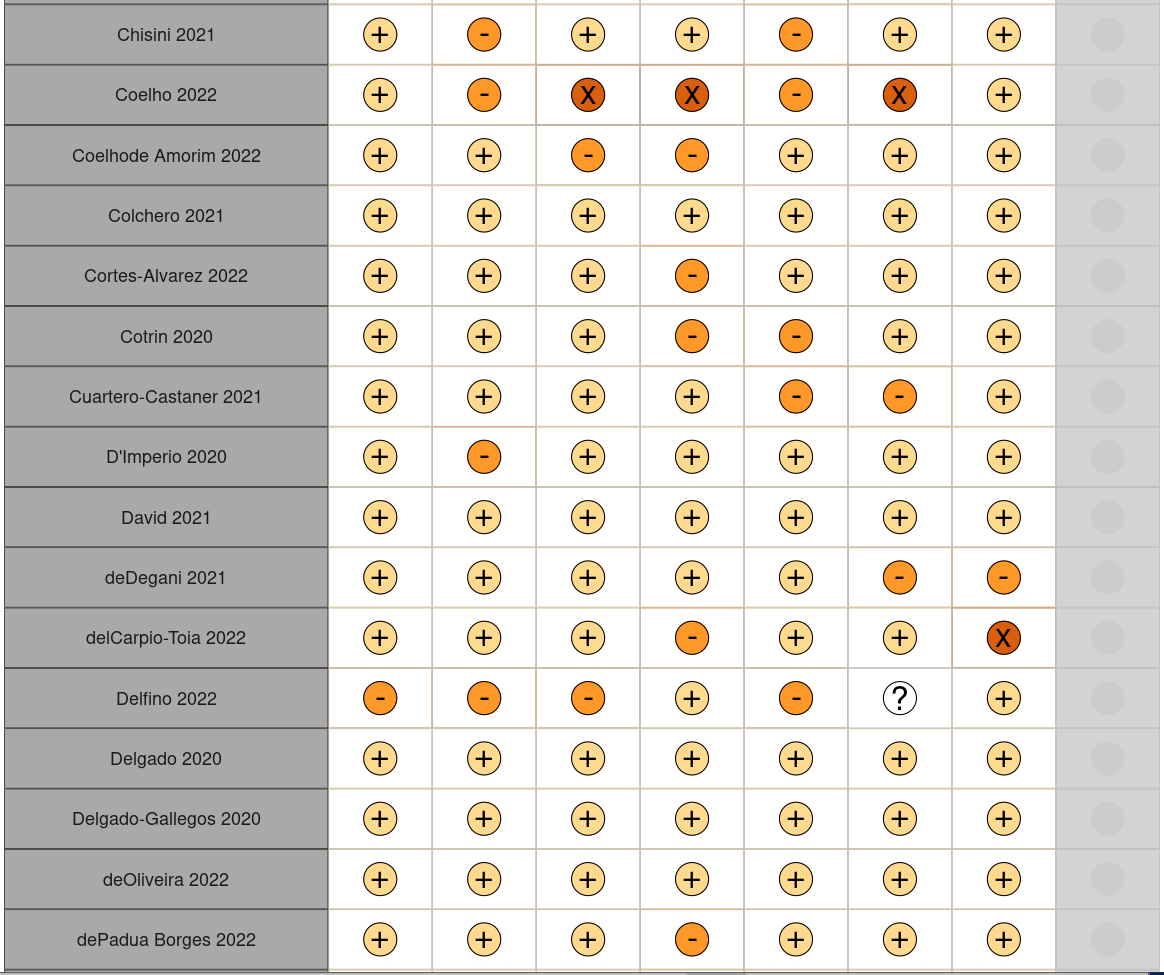 |
| 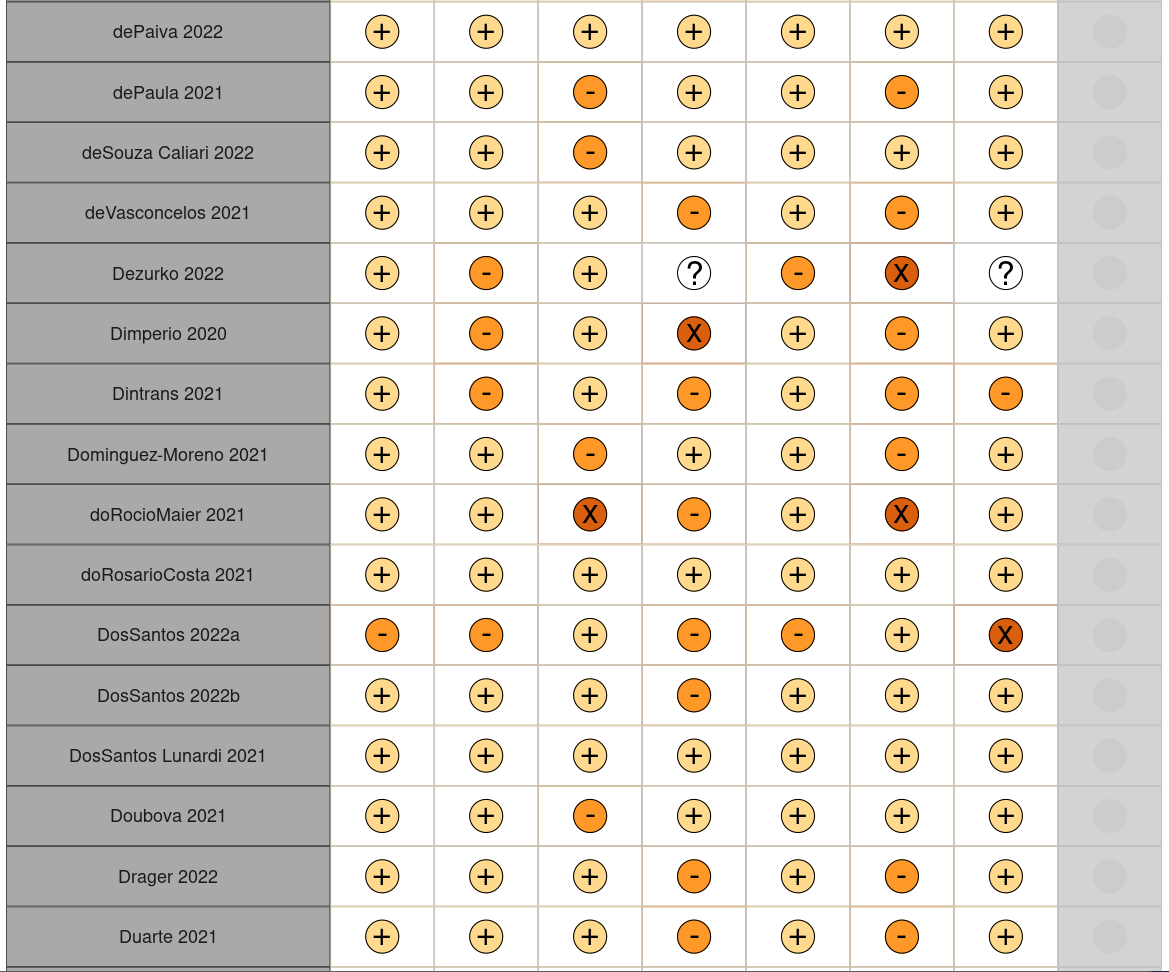 | 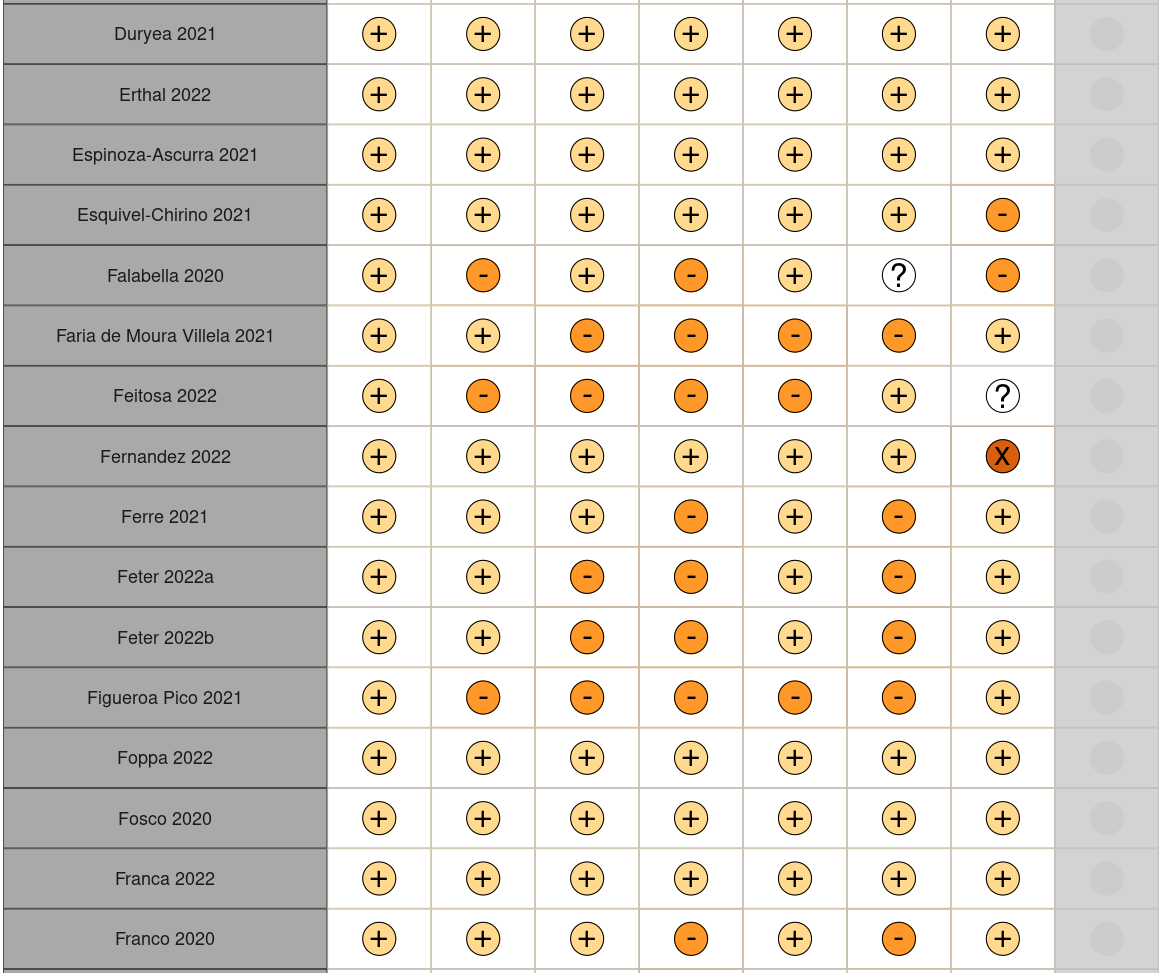 |
| 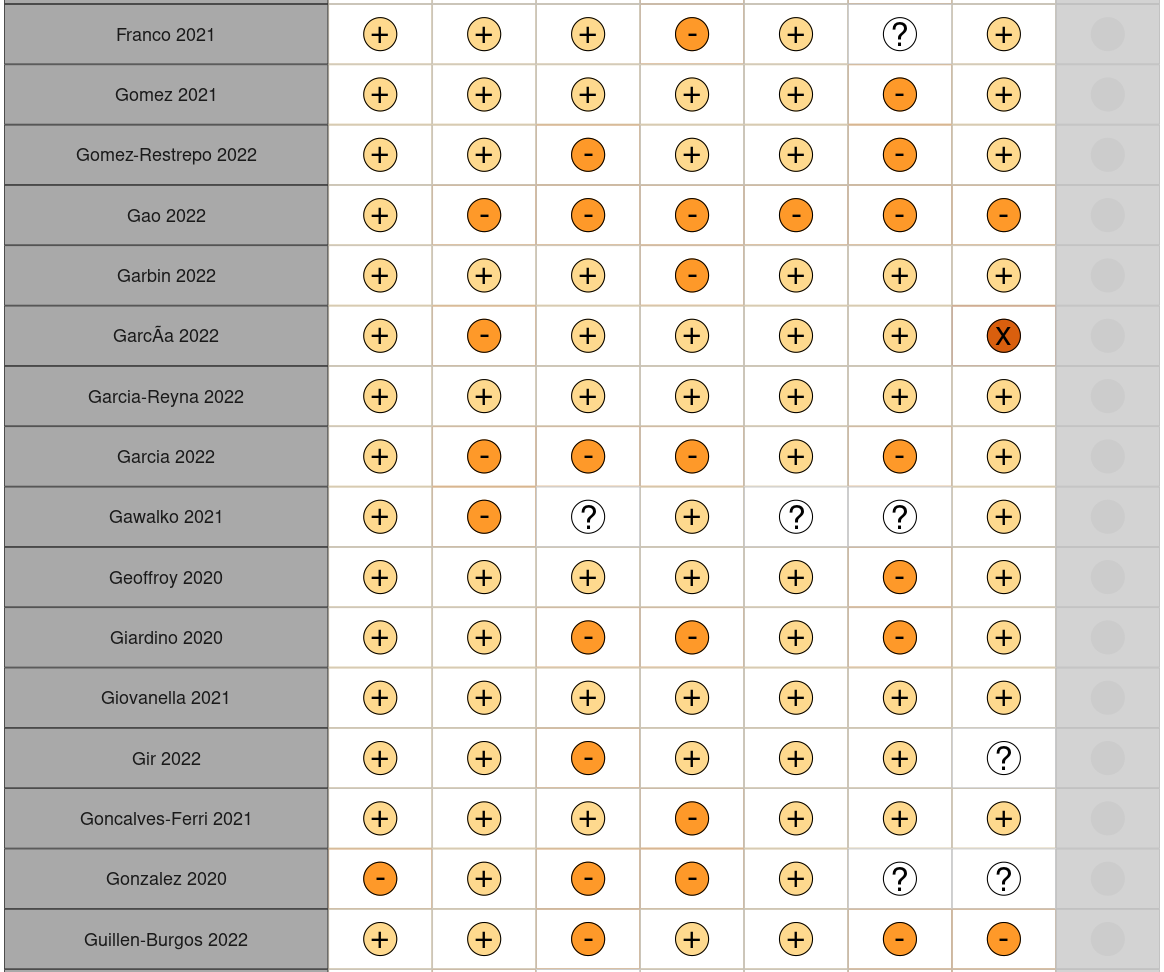 | 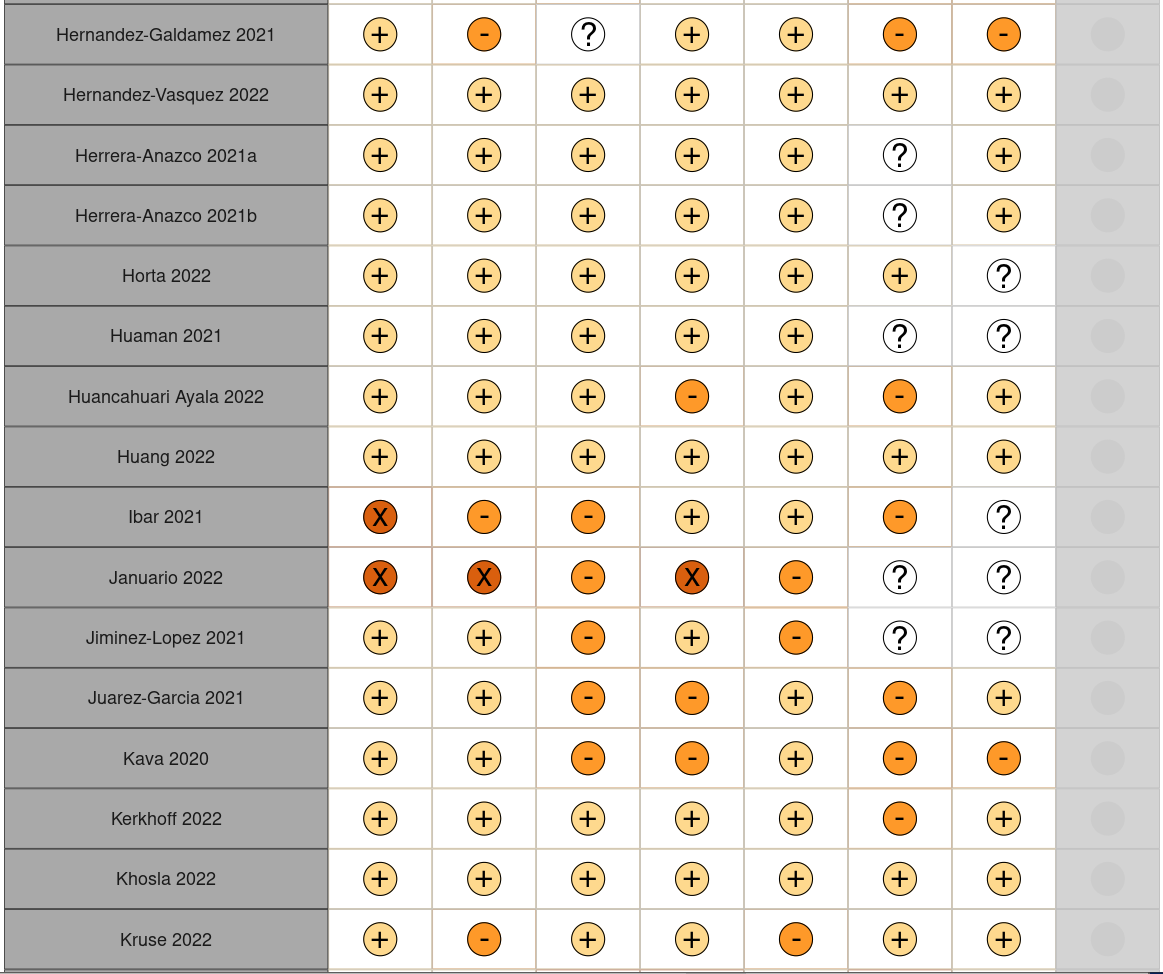 |
| 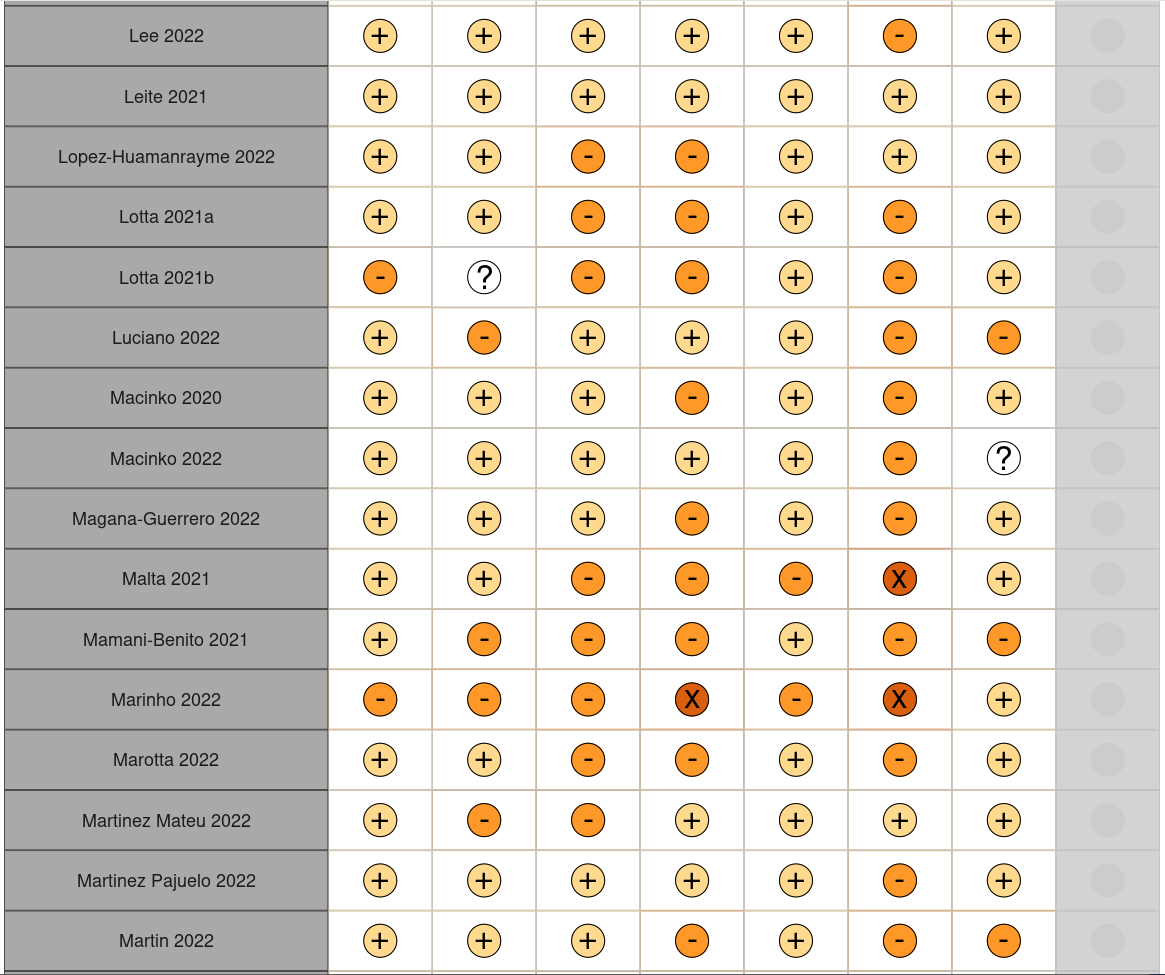 | 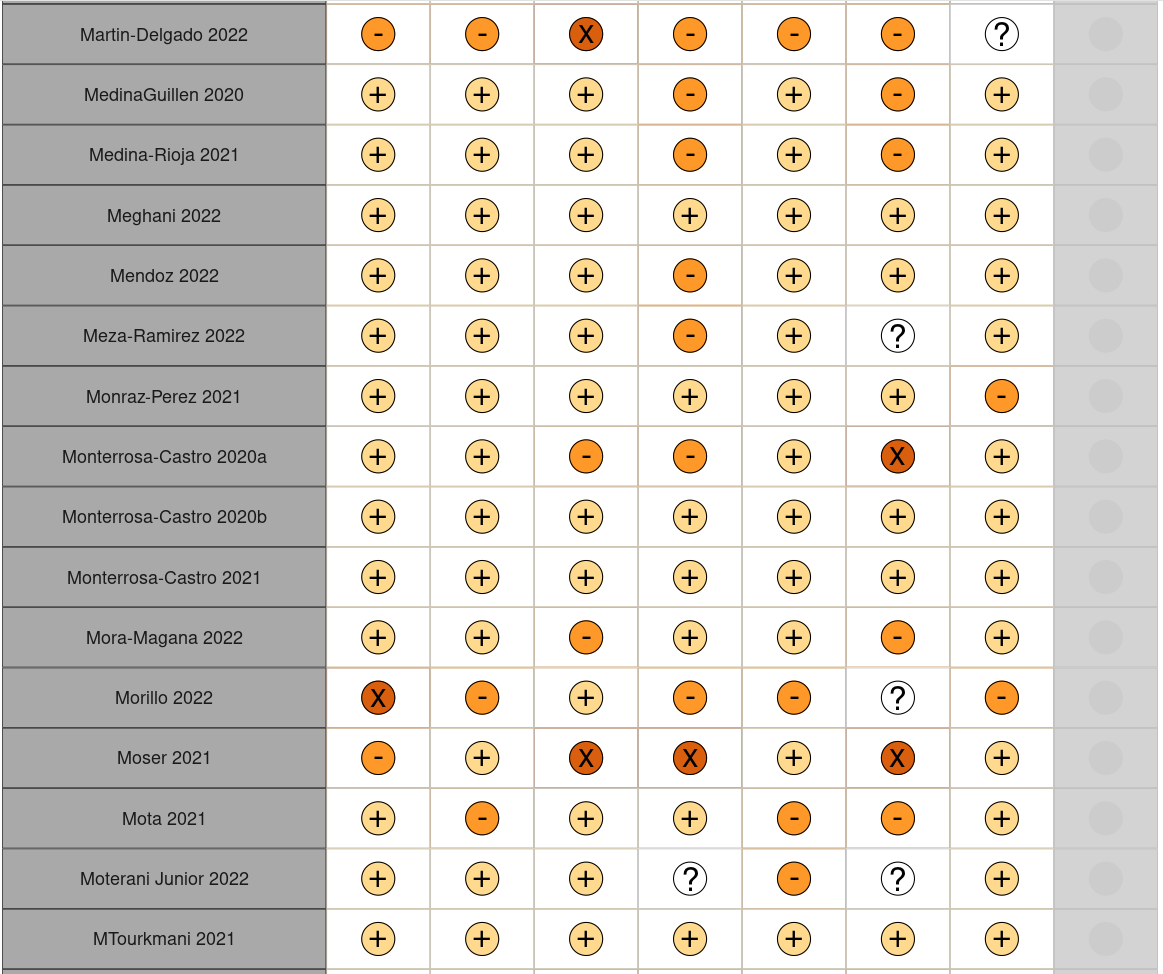 |
| 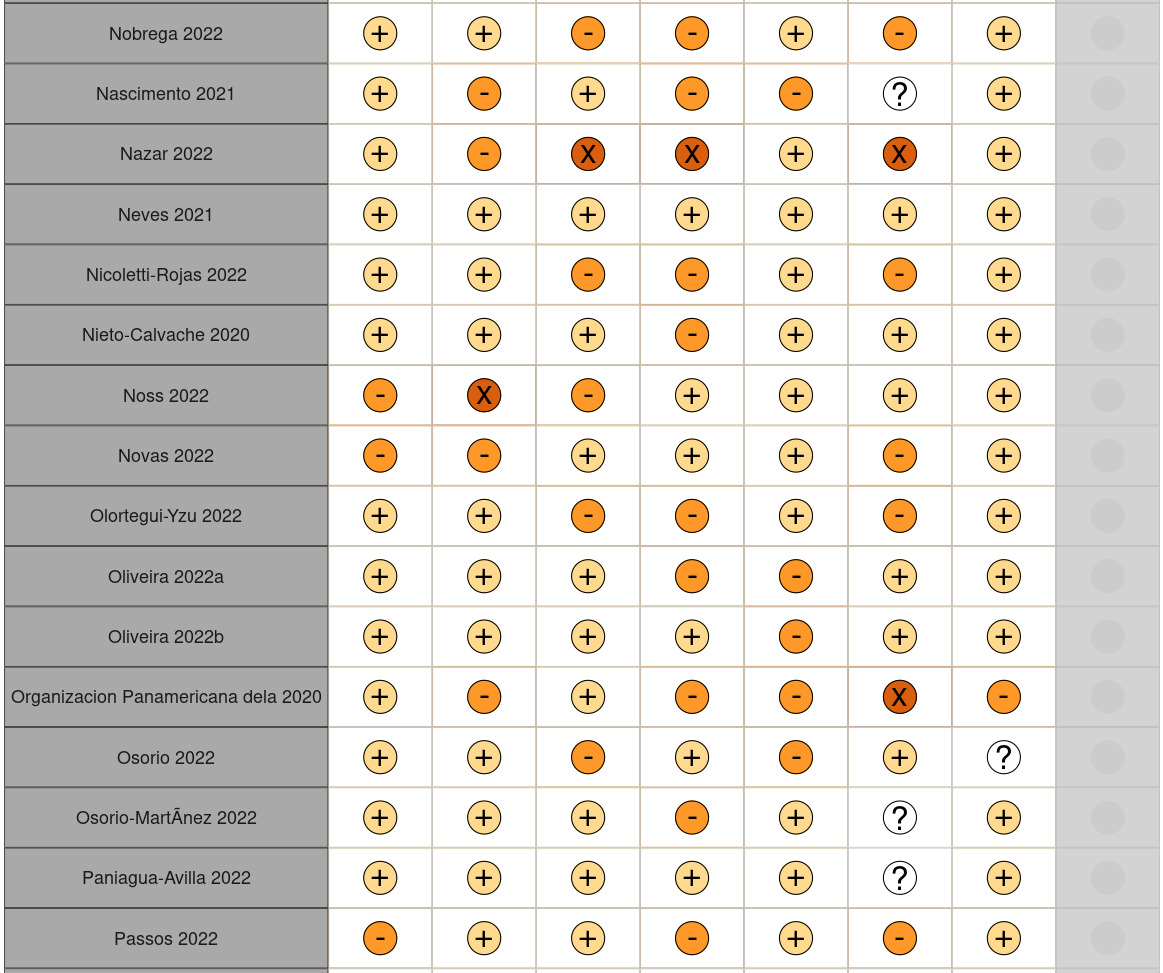 | 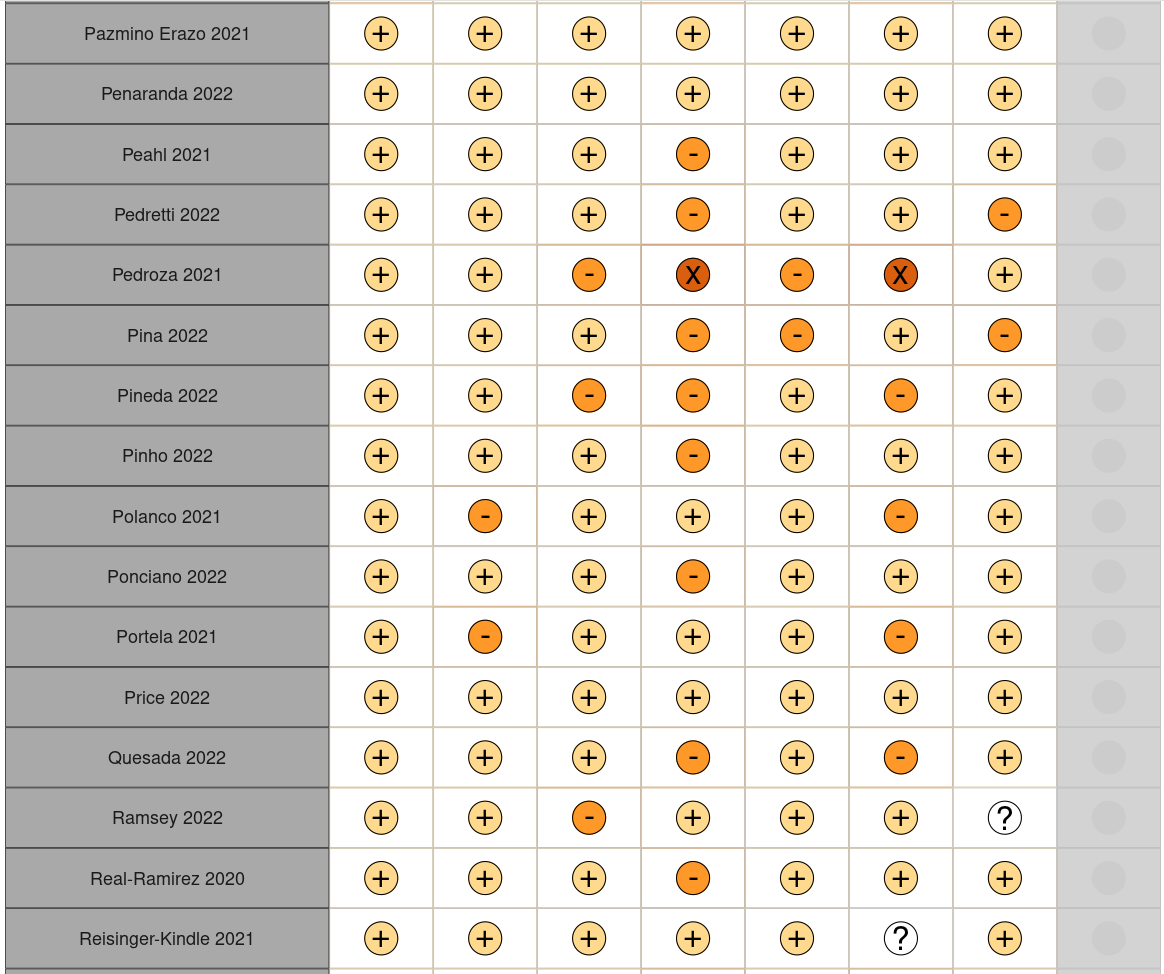 |
| 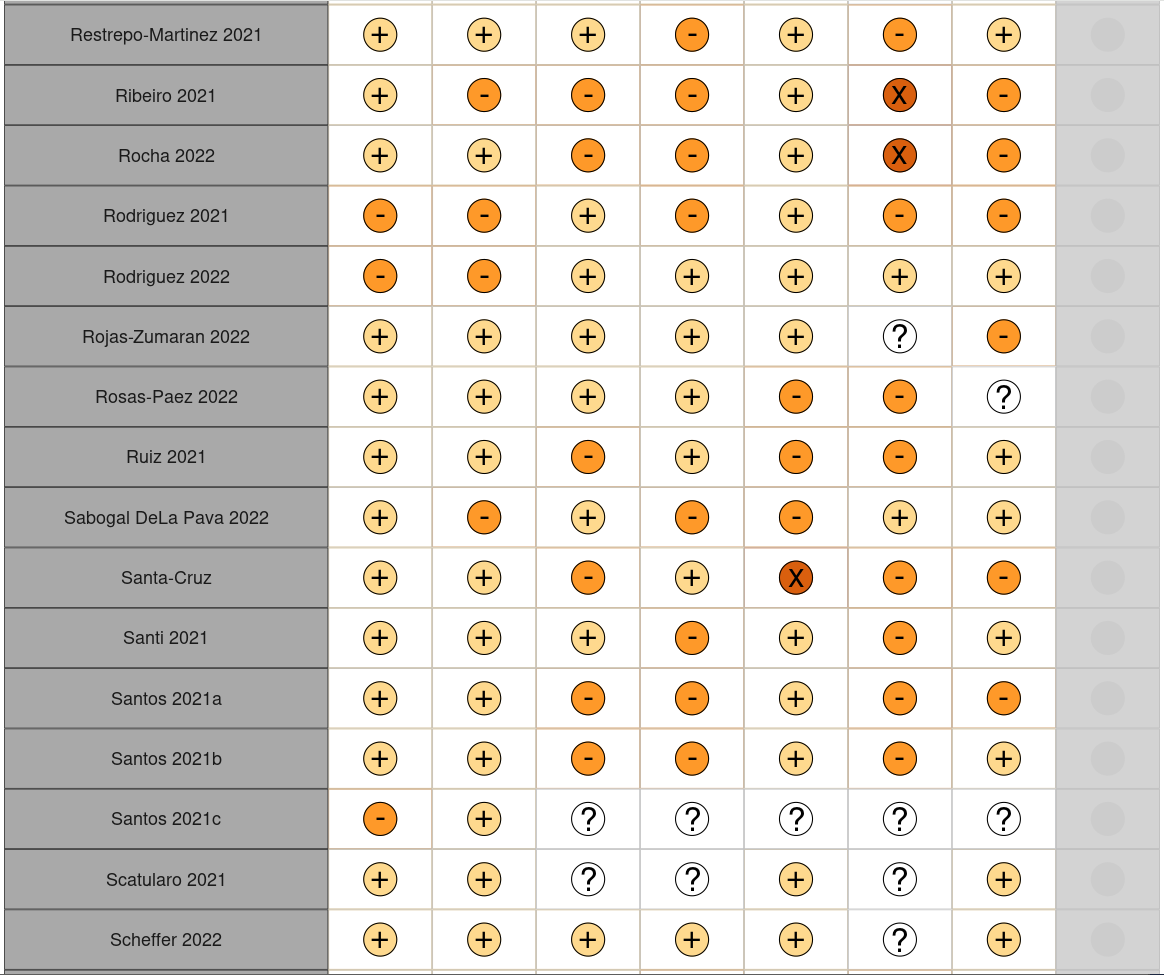 | 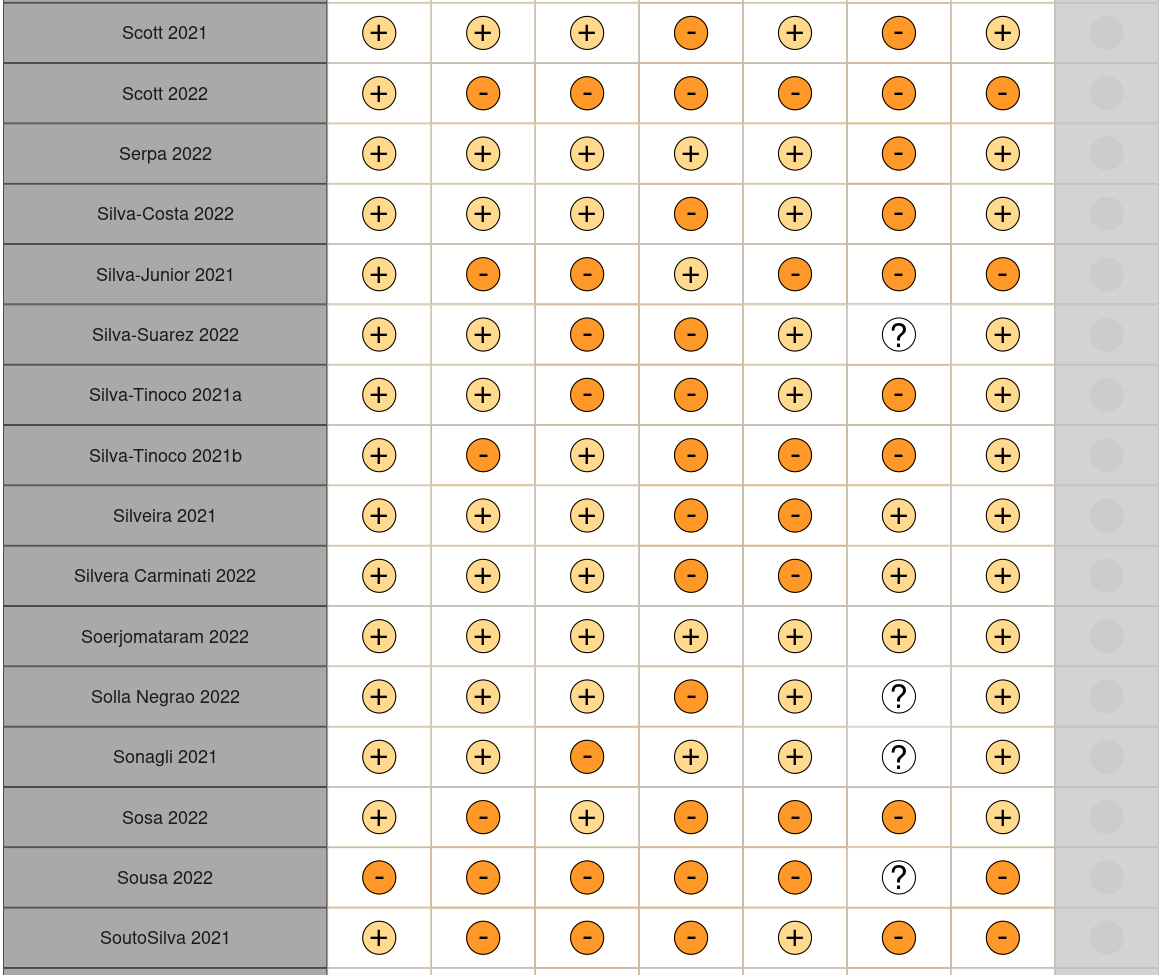 |
| 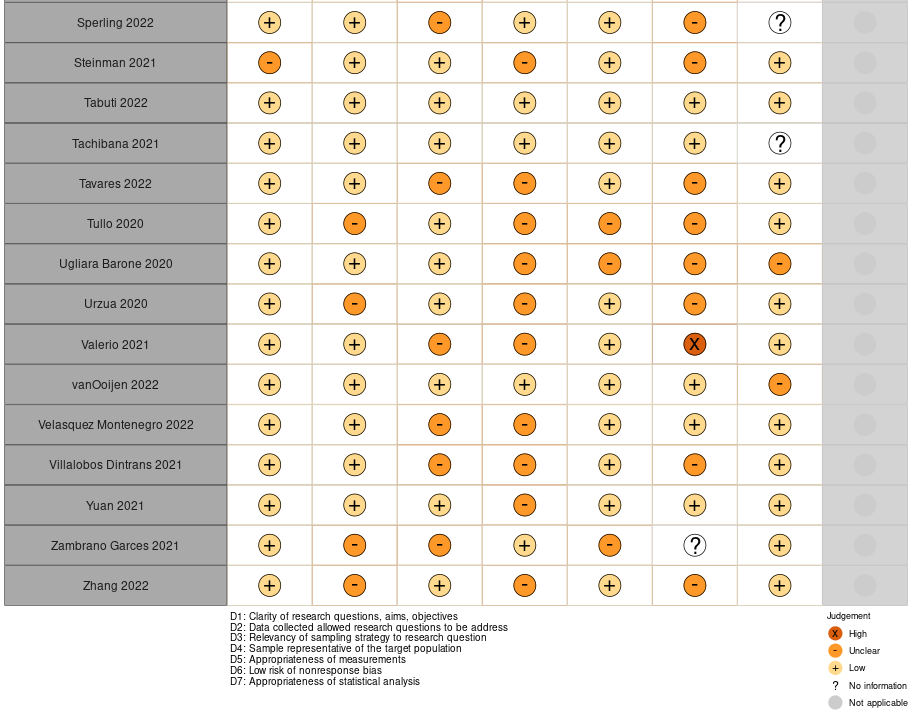 | |

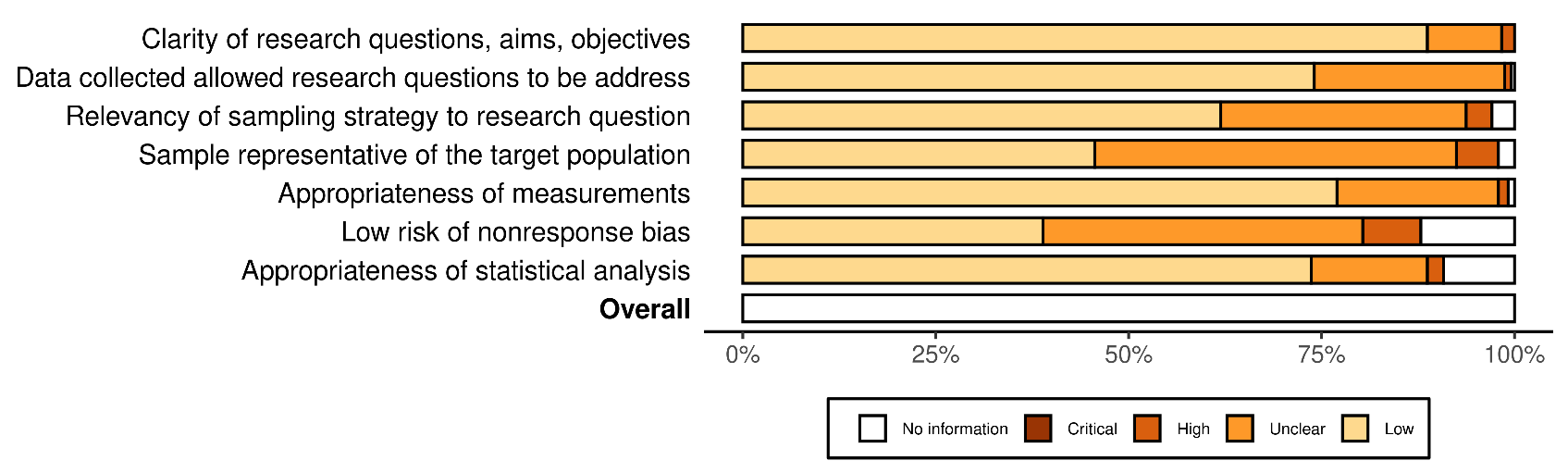

**Figure e6: Risk of bias assessment summary for mixed methods studies**

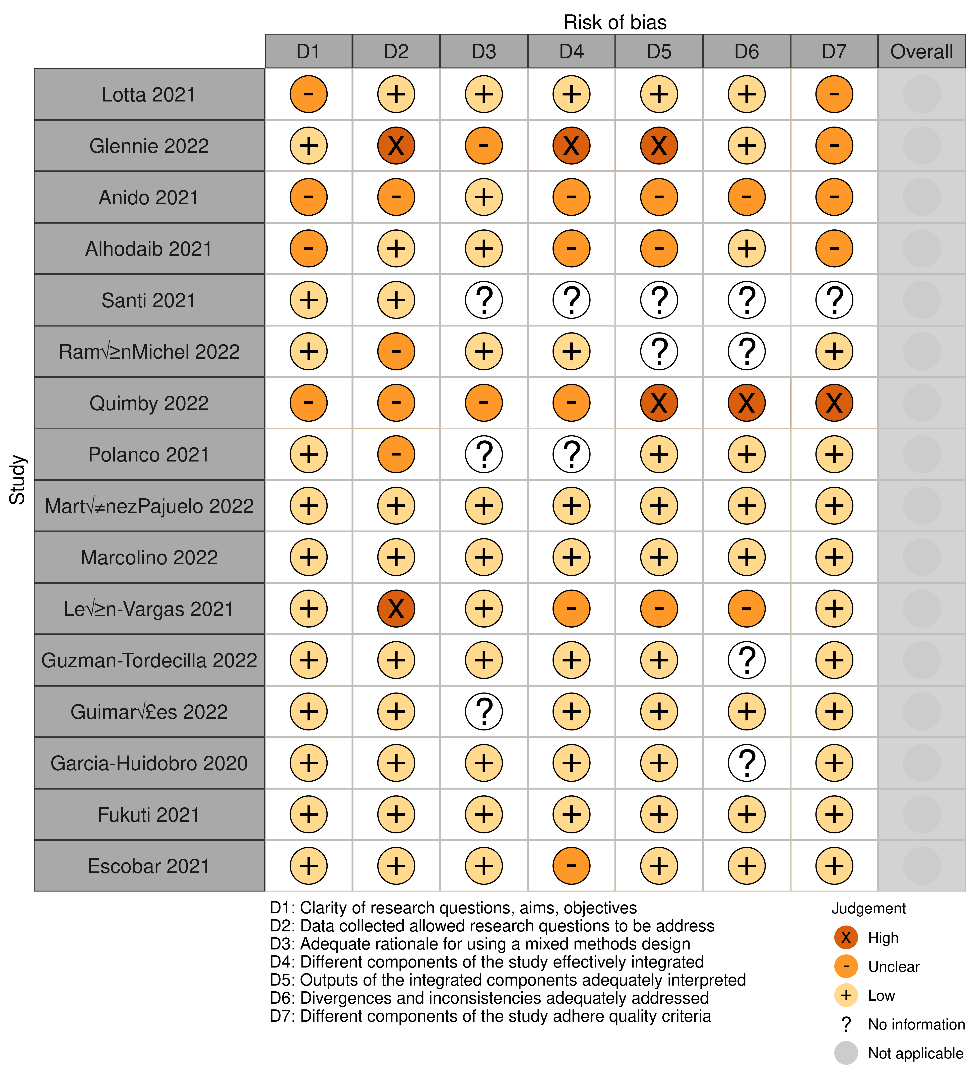

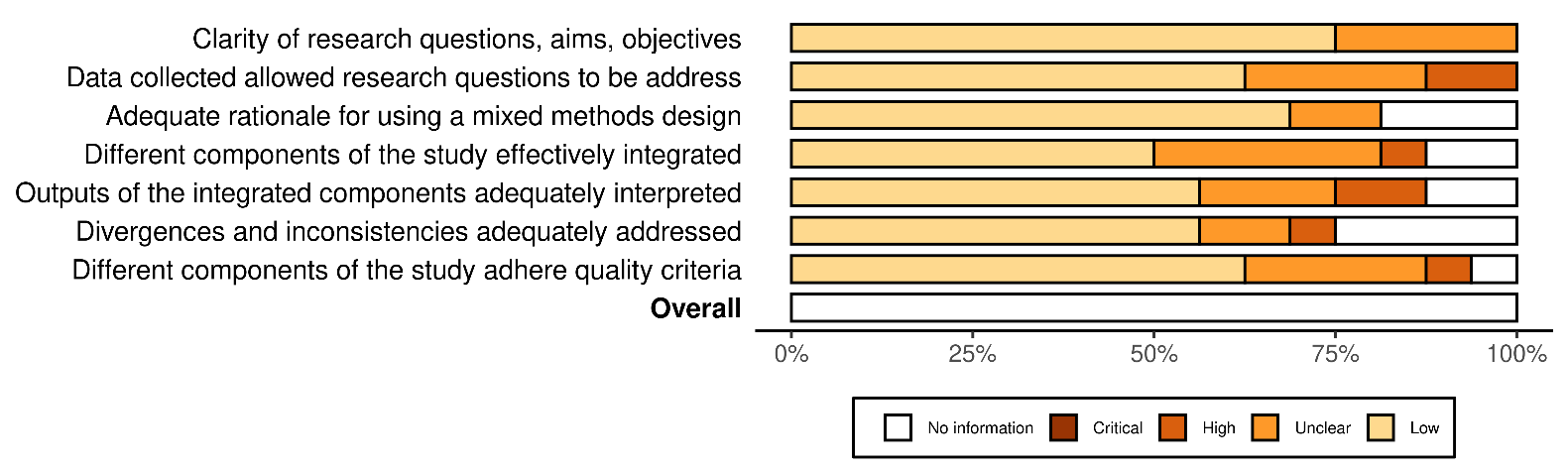

**Figure e7. Clinical condition addressed, by thematic area of interest (disruptions vs. intervention)**

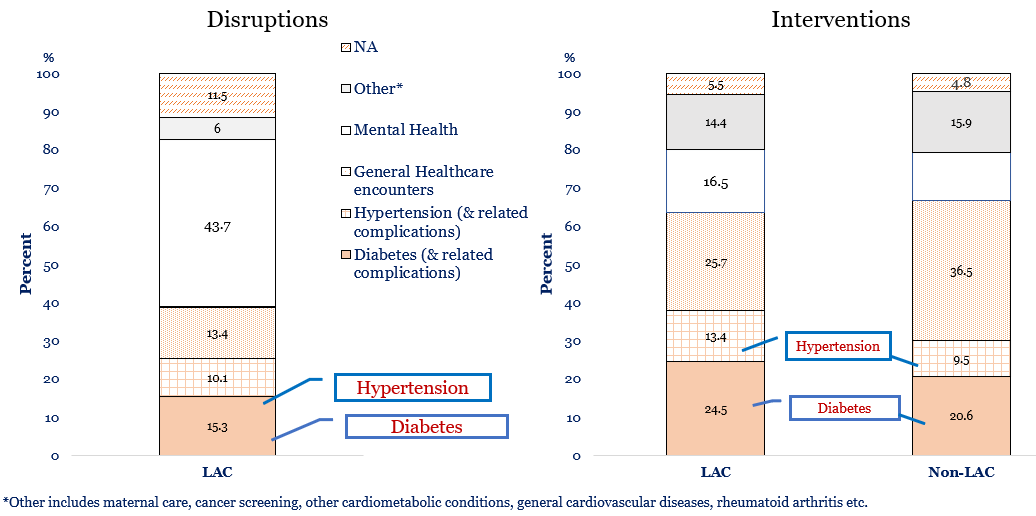

**Figure e8. Geographical representation of all included studies**

**A**

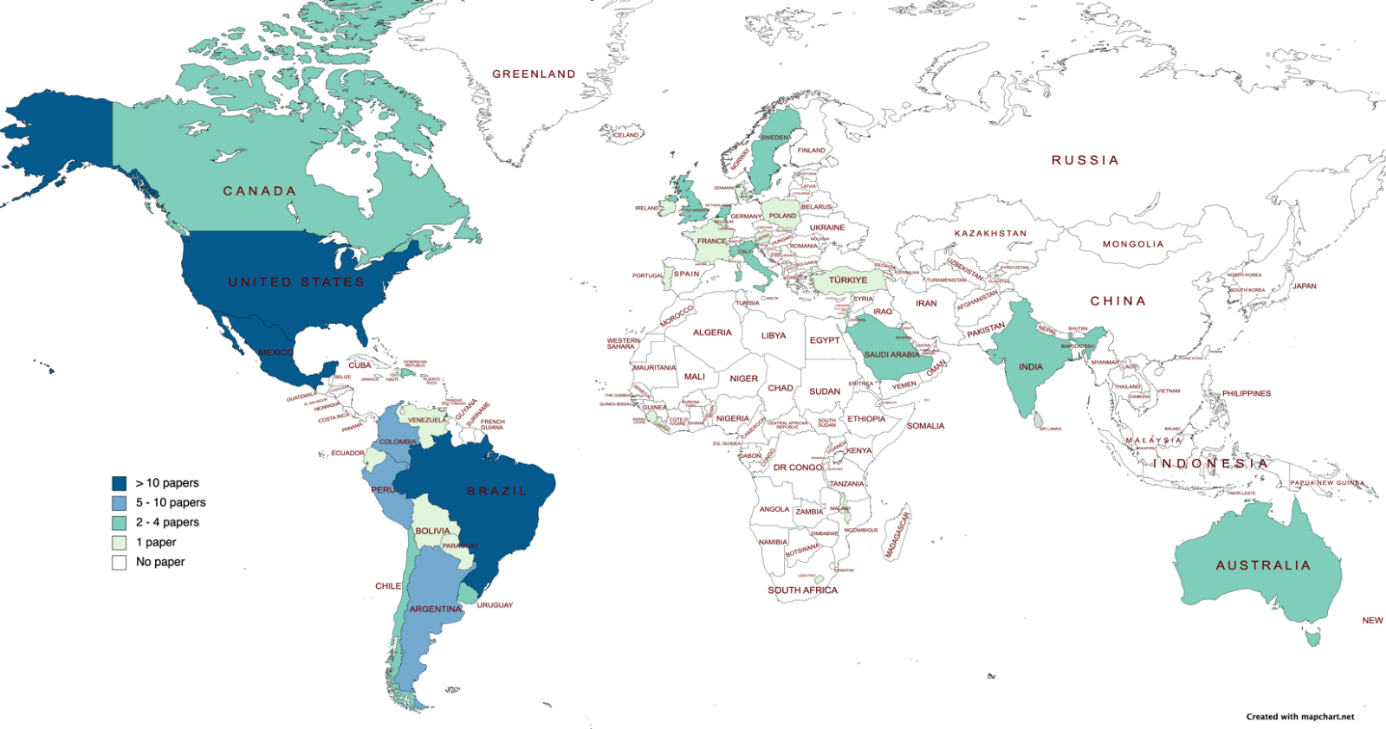

**Figure e9. Disruption: Clinical condition addressed, by Latin America and Caribbean (LAC) countries represented**

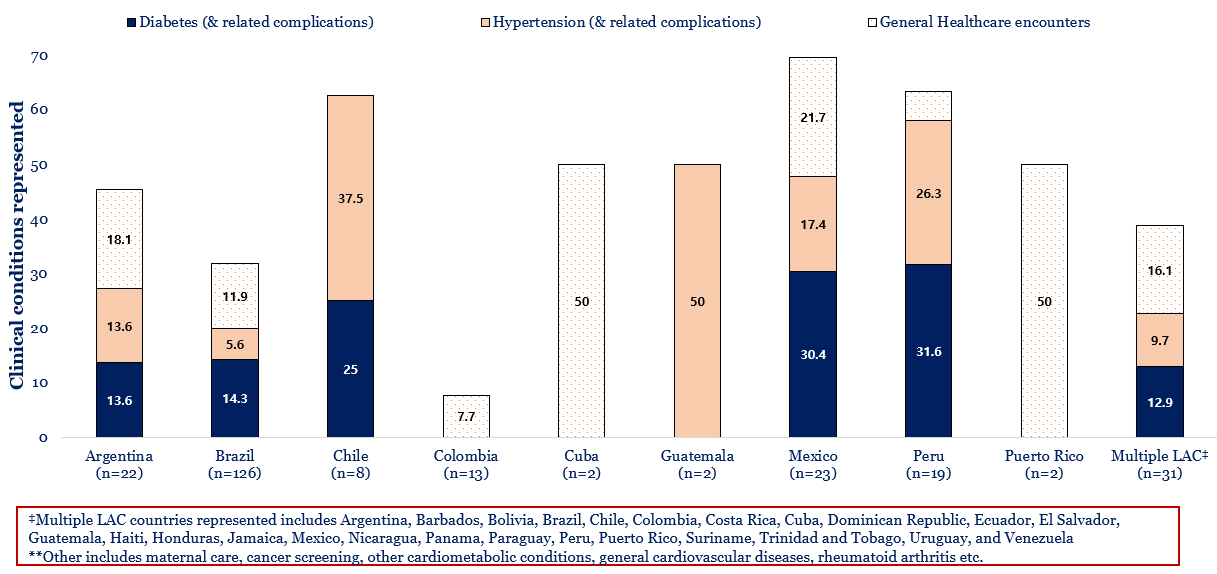

**Table e5. Characteristics of studies, primary care services domains impacted during the COVID-19 pandemic in Latin America and the Caribbean (LAC)**

| **Author, year** | **Country** | **Participants** | **System-level domains** | | | | | | **Service delivery** | | | | | | **Practice performance** | | | | | **Consequences** | | | | | **Condition** | | | | | **Outcomes** | |  |  |  |
| --- | --- | --- | --- | --- | --- | --- | --- | --- | --- | --- | --- | --- | --- | --- | --- | --- | --- | --- | --- | --- | --- | --- | --- | --- | --- | --- | --- | --- | --- | --- | --- | --- | --- | --- |
|  |  |  | Governance/leadership | Finance/Funding | Drug/Supplies | Infrastructure | Information System | Workforce | Pop Health Mgt | Facility Org | Access/Availability | Quality | Screening/Lab | Patient perception Perception | Process measures | Awareness | Detection/Diagnosis | Treatment | Control | Health status | Responsiveness | Equity | Efficiency | Resilience | Diabetes | Hypertension | | Complications | Other |  | | Negative | No Change | Positive |
| **Mesoamerica** |  |  |  |  |  |  |  |  |  |  |  |  |  |  |  |  |  |  |  |  |  |  |  |  |  | |  |  |  | |  |  |  |  |
| Quesada 2022 | Costa Rica | Patients |  |  |  | • |  |  |  |  | • | • | • |  |  |  | • |  |  | • | • | • | • | • | • | |  |  |  | | Death from Diabetes | * |  |  |
| Rodriguez 2021 | Honduras | Physicians, Nurses | • | • |  |  |  | • | • |  | • | • |  | • | • |  |  | • | • |  |  |  | • | • |  | |  |  | • | | Anxiety and Depression | * |  |  |
| Paniagua-Åvila 2022 | Guatemala | Healthcare workers |  |  |  |  | • | • |  |  |  |  |  |  |  |  |  |  |  |  |  |  |  | • |  | |  |  | • | | Mental Distress & depression | * |  |  |
| Hernandez-Galdamez 2021 | Guatemala | Patients |  |  | • |  |  | • |  |  | • |  |  | • |  |  |  | • | • |  |  |  | • | • |  | | • |  |  | | Medication Access & Adherence | * |  |  |
| Caperon 2021 | El Salvador | Patients; HCW | • |  |  |  |  |  | • |  | • |  |  | • |  |  | • |  |  |  |  |  | • | • |  | |  | • |  | | NCD awareness & Digital Access | * |  |  |
| Silva-Tinoco 2021 | Mexico | Patients |  |  |  |  |  |  |  |  |  |  |  |  |  |  |  | • |  |  |  |  |  |  | • | |  |  |  | | Coping with diabetes  Selfcare activities |  |  | *  * |
| Real-Ramirez 2020 | Mexico | HCW |  |  |  |  |  | • |  |  |  |  | • |  |  |  | • |  |  |  |  |  |  |  |  | |  |  | • | | Burnout | * |  |  |
| Pineda 2022 | Mexico | Patients |  |  |  |  |  | • |  |  |  |  |  | • |  |  |  | • | • |  |  | • |  |  | • | |  |  |  | | Treatment Adherence |  |  |  |
| Jimenez-Lopez 2021 | Mexico | HCW |  |  |  |  |  | • |  | • | • | • | • | • |  |  |  | • | • |  | • |  | • | • |  | |  |  | • | | Mental health diagnosis | * |  |  |
| Meza-Ramirez 2022 | Mexico | General practitioners |  |  |  |  |  | • |  | • |  | • |  |  |  |  |  |  | • |  |  |  |  | • |  | |  |  | • | | Depression | * |  |  |
| Meza 2021 | Mexico | HCWs | • | • |  | • |  | • |  | • | • |  |  |  | • |  |  |  |  |  | • | • |  |  |  | |  |  | • | | Clinical practice model (from client-based to public health approach) |  |  | * |
| Medina-Rioja 2021 | Mexico | Patients; Physicians |  |  | • | • |  |  |  |  |  |  | • |  |  |  | • | • |  |  | • |  | • | • |  | |  |  | • | | Stroke diagnosis  Stroke management | * |  | * |
| Cuevas-Budhart 2022 | Mexico | Patients |  |  |  | • |  |  |  |  | • |  |  | • |  |  |  | • |  |  |  |  | • | • |  | |  | • |  | | Coping & psychological symptoms | * |  |  |
| Rosas-Paez 2022 | Mexico | Physicians; Nurses |  |  |  |  |  | • |  |  |  |  |  |  |  |  |  |  |  |  |  |  |  | • |  | |  | • |  | | Burnout  Work engagement | *  * |  |  |
| Mora-Magana 2022 | Mexico | HCW | • | • |  | • |  |  |  | • |  | • | • |  | • |  |  | • |  | • | • | • | • |  |  | |  |  | • | | Anxiety, depression, fear of COVID | * |  |  |
| Magana-Guerrero 2022 | Mexico | HCWs |  |  |  |  |  |  |  |  |  |  |  |  |  |  |  |  |  |  |  |  |  |  |  | |  |  | • | | COVID-19 among HCWs | * |  |  |
| Garcia-Reyna 2022 | Mexico | HCW | • |  |  | • | • |  | • | • | • |  |  |  |  |  | • | • |  |  |  | • |  |  |  | |  |  | • | | Fear of COVID-19 | * |  |  |
| Esquivel-Chirino 2021 | Mexico | HCW |  |  |  |  |  | • |  | • | • |  |  |  | • |  |  |  |  | • |  |  |  |  |  | |  |  | • | | COVID-19 cases, deaths among HCWs | * |  |  |
| Doubova 2022 | Mexico | HCW | • |  |  | • | • | • | • | • | • | • |  |  |  |  |  |  | • | • | • | • | • | • | • | | • |  | • | | Post-COVID-19 recovery of MCH and NCDs services | * |  | * |
| Doubova 2021 | Mexico | NA |  |  |  | • |  |  | • |  |  |  |  |  | • |  | • | • | • | • |  | • | • | • | • | | • |  | • | | Health services disruption | * |  |  |
| Delgado-Gallegos 2020 | Mexico | Patients |  |  |  | • |  | • | • | • |  |  |  |  | • |  |  |  |  | • |  |  | • | • |  | |  |  | • | | Mental health- stress, xenophobia, fear of infection with Covid-19 | * |  |  |
| Cortes-Álvarez 2022 | Mexico | Nurses |  |  |  |  |  | • |  |  | • |  |  |  | • |  |  |  |  |  |  |  | • |  |  | |  |  | • | | Mental health |  |  |  |
| Juarez-Ramirez 2022 | Mexico | HCW |  |  | • |  |  | • |  | • | • |  |  |  |  |  |  | • | • |  |  | • |  | • | • | | • | • | • | | Equipment shortages  Remote monitoring  Care continuity | *  * |  | * |
| Colchero 2021 | Mexico | Patients |  |  |  |  |  |  |  |  | • |  | • | • |  |  | • | • |  |  |  | • |  |  |  | |  |  | • | | Screening and detection  Care seeking | *  * |  |  |
| Arredondo 2021 | Mexico | Patients |  | • |  |  |  |  |  |  | • |  |  |  | • |  |  |  |  |  |  |  |  | • | • | |  |  |  | | Epidemiological burden | * |  |  |
| Alonso 2021 | Mexico | Patients; HCW |  |  | • |  |  | • |  | • |  |  |  | • |  |  |  | • | • |  |  |  | • | • |  | |  |  | • | | Mental health outcomes | * |  |  |
| Juarez-Garcia 2021 | Mexico | HCW |  |  |  |  |  | • |  |  | • | • |  |  | • |  |  |  |  |  |  |  | • |  |  | |  |  | • | | Mental Health Symptoms | * |  |  |
| Cassarino 2022 | Uruguay | Patients | • |  |  |  |  |  |  | • | • |  | • |  |  |  | • | • |  | • | • |  | • |  |  | |  |  | • | | Cervical cancer screening, treatment |  | * |  |
| Silvera Carminati 2022 | Uruguay | Nurses |  |  |  |  |  | • |  |  | • |  |  |  |  |  |  |  |  |  |  |  |  | • |  | |  |  | • | | Physical and mental burden | * |  |  |
| **The Caribbean** |  |  |  |  |  |  |  |  |  |  |  |  |  |  |  |  |  |  |  |  |  |  |  |  |  | |  |  |  | |  |  |  |  |
| Valdés García 2022 | Cuba | Patients |  |  |  | • |  |  |  | • |  |  | • |  |  |  | • | • |  |  |  |  |  | • |  | |  |  | • | | Health service disruption | * |  |  |
| Vázquez-Garay 2021 | Cuba | HCW |  |  |  | • |  | • |  |  | • |  |  |  |  |  |  | • |  |  |  |  | • |  |  | |  |  | • | | Mental Health | * |  |  |
| Alvarez 2022 | Puerto Rico | Nurses; Other HCW |  |  |  |  |  | • |  | • |  | • |  |  |  |  |  |  | • | • |  |  |  | • |  | |  |  | • | | Burnout & Mental health symptoms | * |  |  |
| Silva-Suárez 2022 | Puerto Rico | Pharmacists |  |  |  |  |  | • |  |  | • | • |  |  |  |  |  | • | • |  |  |  |  | • |  | |  |  | • | | Mental health and professional disruptions | * |  |  |
| **Andean Area** |  |  |  |  |  |  |  |  |  |  |  |  |  |  |  |  |  |  |  |  |  |  |  |  |  | |  |  |  | |  |  |  |  |
| Zhang 2022 | Bolivia | HCW; first responders |  |  |  | • |  | • | • |  |  |  |  |  | • |  |  |  |  |  |  | • | • | • |  | |  |  | • | | Retention of HCWs | * |  |  |
| Nicoletti-Rojas 2022 | Chile | Patients |  |  |  | • |  |  | • |  | • |  |  |  |  | • |  | • |  | • |  | • |  |  | • | | • | • | • | | Healthcare access  NCD Self-management | *  * |  |  |
| Urzua 2020 | Chile | HCWs |  |  |  |  |  | • |  |  |  | • |  |  | • |  |  |  |  |  | • |  |  | • |  | |  |  | • | | Mental health symptoms | * |  |  |
| OrtizContreras 2022 | Chile | HCWs |  |  |  |  | • | • | • | • | • | • |  |  | • |  |  |  |  |  | • |  |  | • |  | |  |  | • | | Maternal health initiatives |  |  | * |
| Carvajal 2021 | Chile | Patients | • |  |  | • |  |  |  |  | • |  |  | • | • | • |  |  |  |  | • | • |  | • |  | |  |  | • | | Healthcare access | * |  |  |
| Schongut-Grollmus 2021 | Chile | Patients | • | • | • | • |  |  |  |  | • |  |  | • |  |  |  | • |  |  | • | • | • |  |  | |  |  | • | | Healthcare access (financial) | * |  |  |
| Olivares-Tirado 2022 | Chile | Physicians; Nurses; HCW |  |  |  |  |  | • |  |  | • | • |  |  | • |  |  |  |  |  |  |  | • | • |  | |  |  | • | | Mental health symptoms | * |  |  |
| Barake 2022 | Chile | Patients | • |  |  | • |  |  | • |  |  |  |  |  | • |  |  | • | • |  |  |  |  | • |  | | • |  |  | | Hypertension treatment and control | * |  |  |
| Alvarado 2021 | Chile | Physicians; Other healthcare Workers |  | • |  |  | • | • |  | • | • | • |  |  |  |  | • |  |  | • |  |  | • |  |  | |  |  | • | | Mental health symptoms | * |  |  |
| ArangoSoler 2022 | Colombia | Physicians; Nurses | • | • | • |  |  | • |  |  | • |  |  |  |  |  |  |  |  | • | • |  |  | • |  | |  |  | • | | Physical and mental symptoms | * |  |  |
| Monterrosa-Castro 2021 | Colombia | General practitioners |  |  |  |  |  | • |  | • | • |  |  |  |  | • | • |  |  |  |  |  |  | • |  | |  |  | • | | Mental health symptoms | * |  |  |
| Guillen-Burgos 2022 | Colombia | General practitioners; Physicians; Nurses | • | • | • | • |  | • | • | • |  | • | • |  | • | • |  |  |  | • | • | • | • | • |  | |  |  | • | | Mental health symptoms | * |  |  |
| Sabogal De La Pava 2022 | Colombia | Other |  |  | • |  |  |  | • |  | • | • |  |  | • |  |  | • |  | • |  |  | • |  |  | |  |  | • | | Medication shortage | * |  |  |
| Fernandez 2022 | Colombia | HCW |  |  |  |  |  | • |  | • | • |  |  |  |  |  |  |  | • |  |  |  |  | • |  | |  |  | • | | Burnout and anxiety | * |  |  |
| Castiblanco-Montanez 2021 | Colombia | Patients |  |  | • | • | • |  | • |  | • |  |  |  | • |  |  |  | • | • | • |  | • | • |  | |  |  | • | | Telehealth vs In-person visits |  | * |  |
| Restrepo-Martinez 2021 | Colombia | HCW |  |  |  |  |  | • |  |  | • |  |  |  | • |  |  |  |  |  |  |  | • |  |  | |  |  | • | | Anxiety and depression | * |  |  |
| Penaranda 2022 | Colombia | HCW | • | • |  |  |  |  | • | • | • |  |  | • | • |  |  |  | • |  |  |  | • | • |  | |  |  | • | | Mental health symptoms | * |  |  |
| Nieto-Calvache 2020 | Colombia | Patients; General practitioners |  |  | • |  |  |  |  |  | • |  |  |  | • |  |  |  |  |  |  |  |  | • |  | |  |  | • | | Maternal health – blood donation & availability of blood products | * |  |  |
| Monterrosa-Castro 2020 | Colombia | General practitioners |  |  |  |  |  | • |  | • | • |  |  |  |  |  |  |  |  |  |  |  |  | • |  | |  |  | • | | Mental health symptoms - anxiety | * |  |  |
| Gomez-Restrepo 2022 | Colombia | Patients |  |  |  | • | • | • | • | • | • | • |  | • | • |  | • | • | • | • | • |  | • | • |  | |  |  | • | | Mental healthcare access | * |  |  |
| HernandezRincon 2021 | Colombia | Not Applicable | • |  |  |  |  |  | • | • |  |  |  |  | • |  | • | • | • |  | • |  | • | • |  | |  |  | • | | Primary healthcare resilience |  | * |  |
| Pazmino Erazo 2021 | Ecuador | HCWs |  |  |  |  |  | • |  |  | • |  |  |  | • |  |  |  |  |  |  | • | • |  |  | |  |  | • | | Mental health symptoms | * |  |  |
| Franco 2020 | Ecuador | Nurses |  |  |  |  |  | • |  |  | • |  |  |  |  |  |  |  |  |  |  |  | • |  |  | |  |  | • | | Mental health symptoms | * |  |  |
| Cuartero-Castaner 2021 | Ecuador | Not Reported |  |  |  | • |  |  |  | • | • | • |  |  | • |  |  |  |  |  | • |  | • |  |  | |  |  | • | | Burnout syndrome | * |  |  |
| Bonilla-Sierra 2021 | Ecuador | Medical Residents |  |  |  |  |  | • |  |  | • |  |  |  | • |  | • |  |  |  |  |  | • | • |  | |  |  | • | | Mental health symptoms | * |  |  |
| Figueroa Pico 2021 | Ecuador | Nurses; Other HCW |  |  |  |  |  | • |  | • |  | • |  |  | • | • |  |  |  |  |  |  | • | • |  | |  |  | • | | Mental health symptoms | * |  |  |
| Zambrano Garces 2021 | Ecuador | HCWs |  |  |  |  |  | • |  | • | • |  |  |  |  |  |  |  |  |  |  |  | • | • |  | |  |  | • | | Mental health symptoms | * |  |  |
| Herrera-Anazco 2021 | Peru | Other: database analysis | • | • | • |  |  |  |  |  | • | • |  |  |  |  |  | • |  |  |  | • | • | • | • | | • |  |  | | Drug shortages | * |  |  |
| Cardenas-Pineda 2022 | Peru | Patients |  |  |  | • |  |  | • | • | • |  | • |  | • |  |  |  |  | • |  |  |  | • |  | |  |  | • | | Maternal care | * |  |  |
| Azabache 2022 | Peru | HCWs |  |  |  |  |  | • |  |  | • |  |  |  | • |  |  |  |  |  |  |  | • | • |  | |  |  | • | | Mental health symptoms | * |  |  |
| Velasquez Montenegro 2022 | Peru | Patients |  |  | • |  |  |  |  |  | • |  | • | • |  |  |  |  | • |  | • |  | • |  |  | | • |  |  | | Medication non-adherence | * |  |  |
| Huaman 2021 | Peru | HCW and non-HCW |  |  | • | • |  |  |  |  | • | • |  |  | • |  |  |  |  |  |  |  | • | • |  | |  |  | • | | Mental health symptoms | * |  |  |
| delCarpio-Toia 2022 | Peru | HCWs |  |  |  | • |  | • | • |  | • |  |  |  | • |  |  |  | • |  |  |  | • | • |  | |  |  | • | | Burnout syndrome | * |  |  |
| Rojas-Zumaran 2022 | Peru | Patients |  |  |  | • |  |  | • | • | • |  | • |  | • | • | • |  |  | • |  |  |  | • |  | |  |  | • | | Cancer screening rates | * |  |  |
| Osorio-Martinez 2022 | Peru | HCWs |  |  |  |  |  | • |  | • | • |  |  |  |  | • | • |  |  |  |  |  |  | • |  | |  |  | • | | Mental health symptoms | * |  |  |
| Olortegui-Yzu 2022 | Peru | HCW |  |  |  |  |  | • | • |  | • |  |  |  | • | • | • |  | • |  | • | • | • | • |  | |  |  | • | | Mental health symptoms | * |  |  |
| Mamani-Benito 2021 | Peru | HCWs |  |  |  |  |  | • |  | • | • |  |  |  | • |  | • |  |  | • |  |  | • |  |  | |  |  | • | | Mental health symptoms | * |  |  |
| Zafra-Tanaka 2022 | Peru | HCWs |  |  | • | • | • |  |  | • | • |  |  |  |  |  |  | • | • |  |  |  | • | • | • | |  |  |  | | Healthcare access | * |  |  |
| Tenorio-Mucha 2022 | Peru | NA |  |  | • |  |  |  |  |  | • |  |  |  |  |  |  | • |  |  |  |  | • | • | • | | • |  |  | | Medication availability  Medication delivery | *  * |  |  |
| Martinez Pajuelo 2022 | Peru | Patients |  | • |  |  |  | • | • | • | • |  |  |  | • | • | • |  |  |  | • |  | • | • |  | |  |  | • | | Mental health symptoms | * |  |  |
| Hernandez-Vasquez 2022 | Peru | Patients | • | • |  |  |  | • | • | • |  |  |  |  | • |  | • |  |  | • | • |  |  |  | • | |  |  |  | | Hospital admissions  In-patient mortality | * |  | * |
| Lopez-Huamanrayme 2022 | Peru | Patients; Physicians |  |  |  | • | • |  |  | • | • |  |  |  |  |  |  | • | • |  |  |  | • | • | • | |  |  |  | | In-person healthcare visits  Telehealth visits | * |  | * |
| Espinoza-Ascurra 2021 | Peru | HCWs |  |  |  |  |  | • |  | • | • |  |  |  |  |  |  |  | • |  |  |  | • | • |  | |  |  | • | | Depression | * |  |  |
| Ruiz 2021 | Venezuela | HCWs |  |  |  |  |  | • |  |  |  |  | • |  |  |  | • |  |  |  |  |  | • | • |  | |  |  | • | | Generalized Anxiety Disorder | * |  |  |
| **Southern Cone** |  |  |  |  |  |  |  |  |  |  |  |  |  |  |  |  |  |  |  |  |  |  |  |  |  | |  |  |  | |  |  |  |  |
| Agrest 2021 | Argentina | HCWs |  |  |  |  |  | • |  |  | • |  |  |  | • |  |  |  |  |  |  |  |  | • |  | |  |  | • | | Distress & Uncertainty  Strengthening of HCW Team | * |  | * |
| Carro 2020 | Argentina | Patients |  |  |  |  | • |  |  | • | • |  |  |  |  |  | • | • | • |  | • |  |  | • |  | |  | • |  | | Medical visits | * |  |  |
| Marconi 2022 | Argentina | HCWs |  |  |  |  |  | • |  | • | • |  |  |  | • |  |  |  |  |  |  |  | • |  |  | |  |  | • | | Mental health – psychiatric sick leave | * |  |  |
| Ardila-Gomez 2021 | Argentina | HCWs |  | • |  |  |  |  |  |  | • |  |  |  | • |  |  |  |  |  |  | • |  | • |  | |  |  | • | | Mental health services | * |  |  |
| Fosco 2020 | Argentina | Patient |  |  | • |  |  |  |  |  | • | • | • |  |  |  | • | • | • | • |  |  | • | • |  | | • |  |  | | Severe arterial hypertension | * |  |  |
| Bozovich 2020 | Argentina | Physicians; Medical directors of 31 private centers |  |  |  | • |  |  |  | • | • | • |  |  | • |  |  | • |  |  |  |  |  | • |  | |  |  | • | | Hospital visits | * |  |  |
| Bluro 2022 | Argentina | Public (no HCW) |  |  |  |  |  |  |  |  |  |  |  | • |  |  |  | • |  |  | • |  |  | • |  | |  |  | • | | Medical consultations | * |  |  |
| Appiani 2021 | Argentina | Physicians; Residents |  |  |  |  |  | • |  |  | • |  |  |  | • |  |  |  |  |  |  |  |  | • |  | |  |  | • | | Mental health symptoms | * |  |  |
| D‚ÄôImperio 2020 | Argentina | Health centers |  |  |  | • |  |  |  | • |  |  |  |  |  |  |  |  |  |  |  |  |  | • |  | |  |  | * | | Remote healthcare  Admissions |  |  | *  * |
| Martínez Mateu 2022 | Argentina | Paediatric Patients |  | • | • |  |  |  | • |  | • |  |  | • |  |  |  | • | • | • |  | • | • |  | • | |  |  |  | | Metabolic control | * |  |  |
| Gonçalves-Ferri 2021 | Brazil | Maternity Hospitals |  |  | • | • |  |  |  | • | • |  |  | • | • | • |  |  | • | • |  |  | • | • |  | |  |  | • | | Breastfeeding support for mothers with Covid-19 infection | * |  |  |
| Januario 2022 | Brazil | HCW |  |  |  |  |  | • |  |  |  |  | • |  |  |  | • | • |  |  |  |  | • | • |  | |  |  | • | | Depression | * |  |  |
| Lotta 2021 | Brazil | Nurses; Community health workers |  |  |  |  |  | • |  | • |  |  |  |  |  |  |  |  | • |  |  |  |  | • |  | |  |  | • | | HCWs access to resources | * |  |  |
| Franco 2021 | Brazil | Patients |  |  |  |  |  |  |  |  |  |  |  |  |  |  |  | • |  |  |  |  |  |  | • | |  |  |  | | Treatment & self-care adherence | * |  |  |
| Anido 2021 | Brazil | HCW | • |  |  |  |  | • |  | • |  |  |  |  | • | • |  |  |  |  |  |  | • | • |  | |  |  | • | | Mental health symptoms | * |  |  |
| Zipf 2022 | Brazil | Nurses |  |  | • |  |  | • |  | • | • |  |  |  |  |  |  |  |  |  |  |  |  | • | • | |  |  |  | | Mental health symptoms | * |  |  |
| Ugliara Barone 2020 | Brazil | Patients |  |  | • |  |  |  | • |  |  |  |  | • |  |  |  | • | • | • |  |  |  |  | • | |  |  |  | | Blood glucose or variability | * |  |  |
| Serpa 2022 | Brazil | HCWs |  |  |  |  |  | • |  |  | • |  |  |  |  |  |  |  |  |  |  |  |  | • |  | |  |  | * | | Healthcare worker distress | * |  |  |
| Pinho 2022 | Brazil | HCWs |  |  |  |  |  | • |  | • | • |  |  |  | • |  |  |  |  |  |  |  | • | • |  | |  |  | • | | Burnout | * |  |  |
| Martin 2022 | Brazil | Patients |  |  |  | • |  |  |  |  |  | • |  |  |  |  |  |  |  |  | • |  |  |  |  | |  |  | * | | Maternal care |  | * |  |
| Luciano 2022 | Brazil | Patients |  | • | • | • |  |  |  |  | • | • | • |  | • |  | • | • |  | • |  |  | • |  | • | |  |  |  | | New onset type 1 diabetes | * |  |  |
| Horta 2022 | Brazil | Other: Public |  |  |  | • |  |  |  |  | • |  |  | • |  |  |  |  |  |  | • | • | • | • |  | |  |  |  | | Healthcare utilization | * |  |  |
| Meira-Silva 2022 | Brazil | HCW |  |  |  |  |  | • | • |  | • |  |  |  |  | • |  |  |  | • |  |  | • |  |  | |  |  | • | | Mental health -Burnout | * |  |  |
| Negrao 2022 | Brazil | Patient |  |  |  | • |  |  |  |  | • | • | • |  | • |  | • |  |  |  | • |  |  | • |  | |  |  | • | | Breast Cancer Screening  Time to mammography from symptom onset  Proportion with palpable masses at diagnosis | * | * |  |
| Duarte 2022 | Brazil | Patients |  |  |  | • |  |  |  |  |  |  | • |  | • |  | • |  |  |  | • |  |  |  |  | |  |  | • | | Breast and cervical cancer screening | * |  |  |
| Guimarães 2022 | Brazil | Patients |  | • |  | • | • |  |  | • | • |  | • |  | • |  | • | • | • |  |  | • | • |  | • | |  |  |  | | Hospitalization rate |  |  | * |
| Foppa 2022 | Brazil | Patients |  |  |  |  |  |  |  |  | • |  |  |  |  |  |  | • | • | • |  |  |  | • | • | |  |  |  | | Quality of care | * |  |  |
| Gir 2022 | Brazil | HCW | • | • |  | • |  | • |  | • | • |  |  | • | • |  |  |  |  | • | • |  | • |  |  | |  |  | • | | Use of psychoactive substances by HCWs | * |  |  |
| Feitosa 2022 | Brazil | Patients |  |  |  |  |  |  |  |  |  |  |  |  |  |  | • | • |  |  |  |  |  |  |  | | • |  |  | | Medication adherence |  | * |  |
| França 2022 | Brazil | Patients, HCW |  |  |  |  |  | • |  |  |  |  |  |  | • |  |  |  |  |  |  |  | • | • |  | |  |  | • | | Mental health – depression symptoms | * |  |  |
| Erthal 2022 | Brazil | Patients |  | • |  | • |  |  | • |  |  |  |  | • |  |  |  | • | • | • | • |  |  |  | • | |  |  |  | | Health and lifestyle of patients with diabetes  Eating habits, physical activity, & perceived glycemic control | * | * |  |
| De Vasconcelos 2021 | Brazil | Patients |  |  |  |  |  |  |  |  |  |  |  |  |  |  |  |  | • |  |  |  |  |  | • | |  |  |  | | Physical activity | * |  |  |
| De Padua Borges 2022 | Brazil | Patients |  |  |  | • | • |  |  | • | • |  |  |  |  |  | • | • | • | • |  |  | • |  | • | |  |  |  | | Mental health symptoms | * |  |  |
| CoelhodeAmorim 2022 | Brazil | Patients |  |  | • | • |  |  |  | • | • |  |  |  |  |  |  |  | • | • |  |  | • |  |  | |  |  | • | | Continuity of care | * |  |  |
| Chisini 2021 | Brazil | Patients |  |  |  | • |  | • |  |  | • |  |  |  |  |  |  | • | • |  |  |  |  | • | • | |  |  |  | | Gestational diabetes care | * |  |  |
| Binhardi 2021 | Brazil | Patients |  |  | • | • | • |  |  |  | • |  | • |  |  |  |  | • | • |  |  | • | • | • | • | |  |  |  | | Medication access  Physical activity  Glucose monitoring | *  * |  | * |
| Bessa 2021 | Brazil | Patients | • |  |  |  |  |  |  |  | • |  | • | • | • |  | • |  |  | • |  | • |  | • |  | |  |  | • | | Cancer screening rates  Palpable lump detection | *  * |  |  |
| Bertolli 2022 | Brazil | Patients |  |  | • |  |  |  |  |  | • |  |  | • |  |  |  | • |  |  |  |  |  |  |  | |  |  | • | | Medication access  Home medication delivery  In-person medication delivery | *  * |  | * |
| Bazan 2020 | Brazil | HCW | * |  |  |  |  |  |  | * |  |  |  |  | • |  | * |  |  |  |  |  | • |  |  | |  |  | • | | Psychological distress | * |  |  |
| Santos 2021 | Brazil | Nurses | • | • |  | • |  | • |  |  | • | • | • |  | • |  |  |  |  |  | • |  | • | • |  | |  |  | • | | Workplace changes  Mental health - fear | * |  | * |
| Barros-Areal 2022 | Brazil | HCW |  |  |  |  |  | • |  | • | * |  |  |  | • |  |  |  |  |  |  |  | * | • |  | |  |  | • | | Mental health related work absences | * |  |  |
| Araujo 2022 | Brazil | Public |  |  |  |  |  |  |  |  |  |  |  | • | • |  |  |  |  |  |  | • | • |  |  | |  |  | * | | Trust in primary care services | * |  |  |
| Andraus 2021 | Brazil | Patients; Nurses; caregivers |  |  | • | • |  |  |  |  | • | • |  |  |  |  |  | • | • |  |  |  | • |  |  | |  |  | • | | Worsening symptoms  Timely medication access | *  * |  |  |
| Oliveira 2022 | Brazil | Nurses |  |  |  |  |  | • |  |  | • |  |  |  | • |  |  |  |  |  |  |  | • |  |  | |  |  | • | | Mental health: Major depressive episodes, suicidal ideation, minor psychiatric symptoms | * |  |  |
| Aragão 2022 | Brazil | Physicians, Medical Residents and Fellows | • | • |  | • |  | • |  | • | • |  | • | • | • | • | • |  | • | • |  |  | • | • |  | |  |  | • | | Mental health: stress levels of residents in surgical rotation | * |  |  |
| Leite 2021 | Brazil | Patients |  |  |  | • |  |  |  |  | • |  |  | • | • |  |  |  |  | • | • |  |  |  | • | | • |  | • | | Access to medications  Avoidance of seeking healthcare  Management of NCD | *  *  * |  |  |
| Alvarado 2021 | Chile | Physicians, HCW |  | • |  |  | • | • |  | • | • | • |  |  |  |  | • |  |  | • |  |  | • |  |  | |  |  | • | | Mental health symptoms | * |  |  |
| Nicoletti-Rojas 2022 | Chile | Patients |  |  |  |  |  |  | • |  | • |  |  | • |  | • |  | • |  | • |  |  |  |  | • | | • |  | • | | NCD self-management | * |  |  |
| Dos Santos Lunardi 2021 | Brazil | Patients |  |  | • | • |  |  |  |  | • |  |  |  |  |  |  | • | • |  | • | • | • |  |  | |  |  | • | | Epilepsy care | * |  |  |
| do Rosario Costa 2021 | Brazil | CHW | • |  | • |  |  | • | • | • |  |  |  | • |  | • |  |  |  |  |  | • | • |  |  | |  |  | • | | Perception of safety at work | * |  |  |
| dePaula 2021 | Brazil | HCW & non HCW |  |  |  |  |  | • |  | • | • | • |  |  | • |  |  |  |  |  |  |  | • |  |  | |  |  | • | | Quality of life: Physical  Social relations  Psychological | **  * |  |  |
| dePaiva 2022 | Brazil | HCW |  |  |  |  |  | • |  |  | • |  |  |  | • |  |  |  |  | • |  |  | • |  |  | |  |  | • | | Sickness absenteeism | * |  |  |
| Cualhete 2022 | Brazil | Not applicable | • |  |  |  |  |  | • |  | • |  |  |  |  |  |  |  |  |  |  | • |  |  |  | |  |  | • | | Homeless people access to hygeine and assistive services  Access of people with disabilities to information and Covid-19 protective measures | *  * |  |  |
| deOliveira 2022 | Brazil | Physicians |  | • |  |  |  | • |  | • |  |  |  |  |  |  |  |  |  |  |  |  | • | • |  | |  |  | • | | Workload (Public sector doctors)  Earnings (Public sector doctors) | * |  | * |
| David 2021 | Brazil | Nurses | • |  |  |  |  | • |  | • | • |  |  |  | • |  |  |  |  | • |  |  |  |  |  | |  |  | • | | Covid-19 infection rate and death | * |  |  |
| Cotrin 2020 | Brazil | Physicians, Nurses, Dentists |  |  |  |  |  | • |  | • |  |  |  |  |  | • |  |  |  |  |  |  |  | • |  | |  |  | • | | HCW income, workload, and anxiety | * |  |  |
| Campos 2021 | Brazil | HCW |  |  |  |  |  | • | • |  | • |  |  |  | • |  |  |  |  |  |  | • | • |  |  | |  |  | • | | Mental health – Depression, Anxiety | * |  |  |
| Ca Queiroz 2021ho 2022 | Brazil | HCW |  |  |  |  |  | • |  |  |  |  |  |  |  |  |  |  |  |  |  |  | • | • |  | |  |  | • | | Mental health and Work issues | * |  |  |
| Coelho de Amorim 2022 | Brazil | Patients |  |  | • | • |  |  |  | • | • |  |  |  |  |  |  | • | • | • |  | • | • |  |  | |  |  | • | | Medication use, health promotion activities (discontinued) | * |  |  |
| Brito-Marques 2021 | Brazil | HCW |  |  |  |  |  | • | • |  |  |  |  |  | • |  |  |  |  | • |  |  |  |  |  | |  |  | • | | Mental Health – Sleep quality, Insomnia, Anxiety and Depression | * |  |  |
| Ribeiro 2022 | Brazil | Patients |  | • |  | • | • |  |  |  | • |  | • | • | • | • | • | • | • | • |  |  | • | • |  | |  | • |  | | CVD Hospitalizations  Length of hospital stay | * | * |  |
| Bigoni 2022 | Brazil | NA |  | • |  | • |  | • |  | • |  |  |  |  | • |  |  |  |  |  |  |  |  | • |  | |  |  | • | | Health system functionality- screenings, diagnostic & treatment procedures, physician appointment  HCW jobs | * |  | * |
| Michels 2022 | Brazil | Patients |  | • |  | • |  |  |  | • | • |  | • | • | • |  |  | • |  | • | • |  | • |  |  | |  |  | • | | Maternal health | * |  |  |
| Bahamondes 2022 | Brazil | Policy makers, Health Post managers | • |  |  | • |  |  |  |  | • |  |  |  | • |  |  |  |  |  | * |  | • | * |  | |  |  | * | | Sexual reproductive health – access and quality of care | * |  |  |
| Araujo 2022 | Brazil | Physicians; Medical Residents and Fellows | • | • |  | • |  | • |  | • | • |  | • | • | • | • |  |  | • | • |  |  | • | • |  | |  |  | • | | Stress  Workload | * |  | * |
| Rodrigues de Queiroz | Brazil | Patients |  |  |  | • |  |  |  |  | • |  | • |  |  |  | • |  |  |  | • |  |  | • |  | |  |  | • | | Cancer diagnosis | * |  |  |
| deOliveira 2022 | Brazil | Physicians |  | • |  |  |  | • |  | • |  |  |  |  |  |  |  |  |  |  |  |  | • | • |  | |  |  | • | | Working hours and income of physicians in private hospitals  Working hour and income of Physicians in public hospitals | * |  | * |
| Macinko 2022 | Brazil | Older adults |  |  |  | • |  |  | • | • | • |  |  |  | • |  | • | • |  |  |  | • | • |  |  | |  |  | • | | Health-care utilization  Gender- disparity in health utilization | *  * |  |  |
| Alves 2022 | Brazil | Nurses |  |  |  |  |  | • |  |  |  |  |  |  |  |  | • |  |  |  |  |  |  | • |  | |  |  | • | | Mental health – symptom of psychopathology | * |  |  |
| Tavares 2022 | Brazil | Nurses | • |  |  | • | • | • |  |  |  |  | • |  |  |  | • | • | • |  |  |  | • | • |  | |  |  | • | | Minor Psychiatric disorders- burnout, | * |  |  |
| Souto Silva 2021 | Brazil | HCW | • | • |  |  |  | • |  |  | • |  | • | • | • |  | • | • |  | • | • | • |  | • |  | |  |  | • | | Mental health – Stress and Psychological symptoms | * |  |  |
| Ribeiro 2022 | Brazil | Nurses |  |  |  |  |  | • |  |  | • |  |  |  | • |  |  |  |  | • |  |  |  |  |  | |  |  |  | | Impact on personal, professional life, | * |  |  |
| Macedo Queiroz 2021 | Brazil | Nurses |  |  |  |  |  | • |  |  | • |  |  |  |  | • |  |  |  |  | • |  | • |  |  | |  |  | • | | Mental health | * |  |  |
| Oliveira Lopes 2022 | Brazil | Patients |  |  |  | • |  |  | • |  |  |  |  |  |  | • |  |  | • | • |  |  | • |  |  | | • |  |  | | Impact of social network and support on treatment compliance |  |  | * |
| Garcia 2022 | Brazil | HCW | • | • |  | • |  | • | • |  | • | • |  | • | • | • | • |  | • | • |  | • | • | • |  | |  |  | • | | Mental health – Depression, Anxiety | * |  |  |
| Camacho | Brazil | HCW | • |  |  |  |  | • |  | • |  |  |  |  | • |  |  |  |  |  | • |  | • | • |  | |  |  | • | | Mental health – Fear, Anxiety, Emotional exhaustion | * |  |  |
| Marques 2021 | Brazil | Patients | • | • |  |  |  |  |  |  | • |  |  |  | • |  |  |  |  | • | • |  |  | • |  | |  |  | • | | Cancer diagnosis | * |  |  |
| Montagna 2021 | Brazil & Ireland | Junior Physicians |  |  |  |  |  | • |  |  | • |  |  |  | • |  |  |  |  |  |  |  | • |  |  | |  |  | • | | Professional socialization | * |  |  |
| Tullo 2020 | Paraguay | Not Applicable |  |  |  |  |  |  |  | • | • |  |  |  |  |  |  |  | • |  | • |  | • | • | • | |  |  | • | | Medical consultations for type II DM  Other medical consultations | * |  | * |
| Tachibana 2021 | Brazil | Patients |  |  |  | • |  | • |  |  |  |  | • |  |  |  | • |  |  |  |  |  |  | • |  | |  |  | • | | Breast cancer diagnosis | * |  |  |
| Portela 2021 | Brazil | Patients | • | • | • | • |  |  | • | • |  |  | • |  | • |  | • | • |  |  |  |  | • | • |  | |  |  | • | | Hospitalizations  ICU admissions  Mortality | *  * |  | * |
| Nóbrega 2022 | Brazil | Nurses | • | • |  |  |  | • | • |  | • |  | • |  | • | • |  |  |  | • |  |  | • | • |  | |  |  | • | | Mental health | * |  |  |
| Lotta 2021 | Brazil | HCW |  |  |  | • | • | • |  | • | • | • |  |  | • |  |  |  |  |  | • | • | • | • |  | |  |  | • | | Mental health, Equality, Workforce protection and Vulnerabilities | * |  |  |
| deOliveira 2022 | Brazil | Patients | • |  | • | • |  |  | • |  | • |  | • |  | • |  | • | • |  | • | • |  |  | • |  | |  |  | • | | Preventive and Treatment services | * |  |  |
| Nascimento 2021 | Brazil | Patients |  |  |  | • |  |  | • | • | • |  |  |  | • |  |  |  |  | • |  |  |  |  |  | |  |  | • | | Hospitalization rates from coronary events  Mortality from coronary events | * |  | * |
| De Souza Caliari 2022 | Brazil | Nurses, Nursing technicians and assistants |  |  |  |  |  | • |  |  |  |  |  |  |  |  |  |  |  |  |  |  |  |  |  | |  |  | • | | Quality of Life – physical, environment, psychological domain  Quality of Life – social domain  Workload, Stress and workplace tension | *  * | * |  |
| Chang 2021 | Argentina | Patients | • |  |  |  | • |  | • | • | • |  |  |  |  |  |  | • | • |  | • | • |  | • |  | |  | • |  | | Rehabilitation post discharge from stroke unit |  |  | * |
| Adila-Gomez 2021 | Argentina | HCW |  |  |  |  | • |  |  | • | • |  |  |  |  |  |  | • | • |  | • |  |  | • |  | |  |  |  | | Availability of mental health services for HCWs during pandemic | * |  |  |
| Setinmnan 2021 | Brazil | Patients |  |  |  |  |  |  | • |  |  | • |  | • | • |  | • |  |  | • |  |  |  | • |  | |  |  | • | | Non-SARS COV-2 emergencies; trend and mortality |  | * |  |
| Osorio 2022 | Brazil | HCW |  |  |  | • |  | • |  |  |  |  |  |  | • |  |  |  |  |  |  |  | • |  |  | |  |  | • | | Anxiety  Depression | * | * |  |
| Santos 2022 | Brazil | Patients | • |  |  |  |  |  |  |  |  |  |  | • | • |  |  |  |  | • | • |  |  | • |  | |  | • |  | | CVD hospitalization  Death | *  * |  |  |
| Oliveira 2022 | Brazil | Patients |  |  |  |  |  |  |  |  |  |  |  | • |  |  |  |  | • |  |  |  |  |  | • | |  |  |  | | Healthy lifestyle:  eating habits  physical activity | *  * |  |  |
| Reizo 2022 | Brazil | Nurses | • | • |  |  |  | • | • | • |  |  |  |  | • |  |  |  |  |  | • |  | • |  |  | |  |  | • | | Disvalue, loss of social labor rights | * |  |  |
| Scatularo 2021 | Argentine | Physicians, Nurses and other HCWs |  |  |  |  |  | • |  |  | • |  |  |  | • |  |  |  |  |  |  |  | • |  |  | |  |  | • | | Anxiety  Minor depression  Sleep quality  Alcohol consumption and smoking | *  *  *  * |  |  |
| Gonzalez 2020 | Argentine | Patients |  |  | • |  |  | • |  | • | • |  |  | • |  | • |  |  | • | • |  | • | • |  |  | | • |  |  | | Health care access  Medication access  Unreported to Health care system | *  *  * |  |  |
| Gonzalez 2022 | Argentine | Patients |  |  |  |  |  |  |  | • | • |  |  |  |  |  |  | • |  | • |  |  | • |  |  | |  | • |  | | Delay in stroke evaluation | * |  |  |
| Dezurko 2022 | Argentine | Patients |  | • | • |  |  |  |  |  | • |  |  | • |  |  |  | • | • |  | • | • | • | • |  | | • |  |  | | Impact of Social isolation on control of HTN by income:  Low income  High income | * |  | * |
| D’Imperio 2020 | Argentina | Physicians |  |  |  | • |  |  |  | • | • |  |  |  | • |  |  |  |  |  |  | • | • | • |  | |  | • |  | | 1.Insitution size, structural capabilities  2. Organization and outpatient setup  3. CCU admission and MI treatment | *  * |  | * |
| Bein 2022 | Argentine | Patients |  |  | • |  | • |  |  |  | • | • |  | • |  |  |  |  | • | • | • |  | • | • |  | |  |  | • | | Healthcare coverage  Obtaining medication  Frequency of seizure | * |  |  |
| Loza 2021 | Argentine | Patients |  |  | • |  | • |  |  |  | • | • |  |  | • | • |  |  |  | • |  |  | • | • |  | |  |  | • | | 1.Access to regularly scheduled consultations  2.Access to chronic medication  3. Acute and emergent consultation  4. Use of information technology | *  *  * |  | * |
| Ibar 2021 | Argentine | HCWs |  | • |  | • |  | • |  |  | • | • |  | • | • | • | • |  |  | • |  |  | • | • |  | |  |  | • | | Emotional exhaustion and burnout | * |  |  |
| Giardino 2020 | Argentine | HCWs |  |  |  |  |  | • |  | • |  |  |  |  | • |  |  | • |  | • |  |  |  |  |  | |  |  |  | | 1.Lack of sleep, exhaustion, and depression related symptoms due to COVID-19  2.Use of sleep medicine | * |  | * |
| Fosco 2020 | Argentina | Patients |  |  | • |  |  | • |  |  | • |  | • |  |  |  | • | • |  | • |  |  |  | • |  | | • |  |  | | Mandatory lockdown and arterial HTN:  Unavailability of medicines  Increase in alcohol consumption | * |  | * |
| DeDegani 2021 | Argentine | Patients |  |  |  | • |  |  |  |  |  |  | • |  |  |  | • |  |  |  | • |  |  | • |  | |  |  | • | | Cancer screening | * |  |  |
| Bozovich 2020 | Argentine | Physicians; Medical Directors |  |  |  | • |  |  |  | • | • |  |  |  | • |  |  | • |  |  |  |  |  | • |  | |  | • | • | | NCDs Car | * |  |  |
| Bluro 2022 | Argentine | Public (excluding HCWs) |  |  |  |  |  |  |  |  |  |  |  | • |  |  |  | • |  |  | • |  |  | • |  | |  |  | • | | Fear of seeing a physician, difficulty navigating information system due to pandemic |  |  | * |
| Argest 2021 | Argentine | Physicians; Medical Residents |  |  |  |  |  | • |  |  | • |  |  |  | • |  |  |  |  |  |  |  |  | • |  | |  |  | • | | Stress, exhaustion, anxiety, etc |  |  | * |
| Feter 2022 | Brazil | Other: general population | • | • | • |  |  |  |  | • | • | • |  | • | • | • | • | • | • | • | • |  |  | • | • | | • |  |  | | Management of chronic disease |  |  | * |
| Dos Santos 2022 | Brazil | Nurses, nursing assistants, technicians |  |  |  |  |  | • |  | • |  |  |  |  | • |  |  |  |  |  |  |  | • |  |  | |  |  | • | | Burnout, exhaustion |  |  | * |
| Santos 2021 | Brazil | Patients |  |  |  | • |  |  |  |  | • |  | • | • |  |  | • |  |  |  | • |  |  | • |  | |  |  | • | | Cancer screening / access | * |  |  |
| Rocha 2022 | Brazil | Nurses | • |  |  |  |  | • |  |  | • | • | • |  | • |  |  |  |  |  |  |  |  |  |  | |  |  | • | | Health workers quality of life during the COVID-19 pandemic | * |  |  |
| Rezio 2022 | Brazil | Nurses | • | • |  |  |  | • | • | • |  |  | • | • |  |  |  |  | • |  | • | • | • | • |  | |  |  | * | | Mental health | * |  |  |
| Macedo Queiroz 2021 | Brazil | Nurses |  |  |  |  |  | • |  |  | • |  |  |  | • |  |  |  |  |  |  |  |  | • |  | |  |  | • | | Mental health | * |  |  |
| Passos 2022 | Brazil | Nurses |  |  |  |  |  |  |  |  |  |  |  |  |  |  |  |  |  |  |  |  |  |  |  | |  |  | • | | Mental health |  |  |  |
| Nobrega 2022 | Brazil | Ecological data on Community Health Workers | • |  |  |  |  | • | • | • | • |  |  |  | • |  |  |  |  |  | • |  |  | • |  | |  |  | • | | Changes in work process of community health agent | * |  |  |
| Moser  2021 | Brazil | Health Workers |  |  |  |  |  | • |  |  |  |  |  |  |  |  |  |  |  |  |  |  |  |  |  | |  |  | • | | Mental health | * |  |  |
| Miliauskas 2022 | Brazil | Ecological data on Mental Health Remote Care |  |  |  | • |  |  |  | • | • |  |  |  |  |  |  | • | • |  |  |  |  | • |  | |  |  | • | | Mental Health Care |  |  | * |
| Lacerda 2022 | Brazil | Healthcare Professionals |  |  |  |  |  | • |  |  |  |  |  |  |  |  |  |  |  |  |  |  |  |  |  | |  |  | • | | Mental Health | • |  |  |
| Ferrari 2021 | Brazil | HCW |  |  |  |  |  | • |  |  |  |  |  |  |  |  |  |  |  |  |  |  |  | • |  | |  |  | • | | Mental Health | • |  |  |
| Fernandez 2021 | Brazil | CHW |  |  |  |  |  | • |  | • |  |  |  |  |  |  |  |  |  |  |  |  |  | • |  | |  |  | • | | Changes in practice | • |  |  |
| Faria 2021 | Brazil | Nurses |  |  |  |  |  | • |  |  |  |  |  |  |  |  |  |  |  |  |  |  |  | • |  | |  |  | • | | Mental Health | • |  |  |
| Duarte 2021 | Brazil | Health Systems | • |  |  |  |  |  | • | • |  |  |  |  | • |  |  |  |  |  |  |  |  | • |  | |  |  | • | | Non-communicable diseases | • |  |  |
| Dimperio 2020 | Argentina | Clinical Centers |  |  |  | • |  |  |  |  | • |  |  |  |  |  | • | • | • |  |  |  |  | • |  | |  |  | • | | Cardiovascular Care | • |  |  |
| Coelho 2022 | Brazil | HCW |  |  |  |  |  | • |  |  |  |  |  |  |  |  |  |  |  |  |  |  |  |  |  | |  |  | • | | Mental Health | • |  |  |
| Braghieri 2021 | Brazil | Patients |  |  | • |  |  |  |  |  | • | • |  |  |  |  |  | • |  |  |  |  |  | • |  | |  | • |  | | Peripheric Artery Disease | • |  |  |
| Borges 2020 | Brazil | Patients |  |  | • |  |  | • |  |  | • |  |  | • |  |  | • | • | • |  |  |  | • | • |  | |  |  | • | | Non-communicable diseases | • |  |  |
| Borges 2021 | Brazil | Nurses |  |  |  |  |  | • |  |  |  |  |  |  |  |  |  |  |  |  |  |  |  | • |  | |  |  | • | | Mental Health | • |  |  |
| Andrade 2021 | Brazil | Patients |  |  |  | • |  |  |  |  |  |  | • | • |  | • | • |  |  |  |  |  |  | • |  | |  |  | • | | Cervical Cancer Screening | • |  |  |
| Alencar 2022 | Brazil | Patients |  |  |  |  |  |  | • |  |  | • |  | • |  |  |  | • |  |  |  |  | • | • | • | |  |  |  | |  | • |  |  |
| Steinman 2021 | Brazil | Patients |  |  | • |  |  |  | • | • |  |  |  | • | • |  |  |  |  | • |  |  | • | • |  | |  |  | • | | Emergency visits due to non SARsCov-2 emergencies | * |  |  |
| **Multiple** |  |  |  |  |  |  |  |  |  |  |  |  |  |  |  |  |  |  |  |  |  |  |  |  |  | |  |  |  | |  |  |  |  |
| Camilletti 2022 | 1 | Patients |  | • | • |  |  |  |  |  | • |  |  |  |  |  |  | • | • |  |  | • |  |  | • | | • |  |  | | Medication Discontinuation  Physical activity | *  * |  |  |
| Santi 2021 | 1 | Patients |  |  | • | • |  |  | • |  | • |  |  |  | • | • |  |  | • | • |  | • | • |  |  | |  |  | • | | Healthcare access | * |  |  |
| Ramón-Michel 2022 | 2 | Patients | • | • | • |  |  | • | • |  | • | • |  | • | • | • |  |  |  | • | • | • | • | • |  | |  |  | • | | Maternal care access | * |  |  |
| Martin-Delgado 2022 | 3 | HCW |  |  |  |  |  | • |  |  | • |  |  |  | • |  |  |  |  |  |  |  |  | • |  | |  |  | • | | Mental health symptoms | * |  |  |
| Kruse 2022 | 4 | Patients; advocacy organizations in LAC serving those with NCDs |  |  | • | • |  |  | • |  | • | • |  |  |  |  |  | • | • |  |  | • | • |  |  | |  |  | • | | Treatment delays  Telemedicine accessibility | *  * |  |  |
| Barone 2020 | 5 | Members of the International diabetes federation in LAC | • |  | * |  | • |  |  |  | • |  |  |  |  | • |  | • |  |  | • |  | • |  | • | |  |  |  | | Medication shortage | * |  |  |
| Arsenault 2022 | 6 | Not Applicable |  |  |  |  |  |  |  |  | • |  |  |  |  |  |  |  |  |  |  |  |  | • |  | |  |  | • | | Cancer screening | * |  |  |
| Knaul 2022 | 7 | Not Applicable | • |  |  |  |  |  | • | • |  |  |  |  |  | • |  |  |  |  | • | • |  |  |  | |  |  | • | | Subnational responses cannot replace coordinated national policy |  |  |  |
| Dintrans 2021 | 8 | The decision-makers and implementers of social and health programs and policies | • | • |  | • |  |  |  | • | • | • |  | • | • |  |  |  | • |  | • | • |  | • |  | |  |  | • | | Continuous service function  Service coverage  Coverage and access to health care  All services compared to pre-pandemic  Health services compared to pre-pandemic | *  *  *  * |  | * |
| Becerra-Posada 2021 | 9 | NA |  | • |  |  |  |  |  |  |  |  |  |  |  |  |  |  |  |  |  |  |  |  |  | |  |  | • | | Financial support for health policy and systems research | * |  |  |
| Aveiro-Robalo 2021 | 10 | Healthcare workers |  |  |  |  |  | • |  | • | • |  |  |  | • |  |  |  |  |  |  |  |  | • |  | |  |  | • | | Perception of mental repercussion | * |  |  |
| Zhang 2022 | 11 | HCW, public | • |  |  |  |  | • |  |  | • |  |  |  |  | • |  |  |  |  |  |  | • | • |  | |  |  | • | | Mental health symptoms | * |  |  |
| Villalobos Dintrans 2021 | 12 | Patients | • | • |  |  |  |  |  | • | • |  |  | • |  | • |  | • |  |  |  |  | • | • |  | |  |  | • | | Relation between the perception of coverage before and after the pandemic |  |  | * |
| Villain 2021 | 13 | Managers/supervisors of cancer screening programs | • |  |  | • |  |  | • | • | • | • | • |  | • | • | • |  |  | • | • |  | • | • |  | |  |  | • | | Cancer screening | * |  |  |
| Piña 2022 | 14 | Physicians | • | • | • | • |  | • | • |  | • | • | • | • |  |  | • | • | • |  |  |  | • | • |  | |  | • |  | | Physicians’ perception about patients’ reluctance for CAD testing  Barrier in imaging, i.e., CT, etc  Barrier to invasive CAD diagnosis | *  *  * |  |  |
| Llop-Gironés 2021 | 15 | Nurses | • |  |  | • | • | • |  | • |  |  |  |  | • |  |  |  |  |  | • | • | • | • |  | |  |  | • | | Vulnerability faced by nurses | * |  |  |
| Delgado 2020 | 16 | Physicians; Nurses |  |  |  |  |  | • |  |  |  |  |  |  |  |  |  |  |  |  |  |  |  |  |  | |  |  | • | | Access to essential personal protective equipment | * |  |  |
| Benis 2021 | 17 | General population | * |  |  |  | • |  |  |  |  |  |  | * |  |  |  |  |  |  |  |  | • |  |  | |  |  | • | | Telemedicine after pandemic |  |  | * |
| Antiporta 2020 | 18 | Ministry of Health focal person for mental health | • |  | • |  |  | • | • |  | • |  |  |  | • |  |  |  |  |  |  |  |  | • |  | |  |  | • | | Mental health burden and needs | * |  |  |
| Giovanella 2021 | 19 | Physicians |  | • |  |  |  |  | • | • |  | • |  |  |  | • | • |  |  | • | • |  | • | • |  | |  |  | • | | Integration of Primary Health Care Strategies | * |  |  |
| Luciani 2022 | 20 | NA | • | • | • | • |  |  | • | • | • | • | • |  | • | • | • | • | • |  | • | • | • | • |  | |  |  | • | | NCD services  Adaptive strategies to mitigate service disruptions | * |  | * |
| PAHO 2020 | 20 | Regional offices |  | • |  |  |  | • |  |  | • |  |  |  | • |  |  |  |  |  |  |  |  | • |  | |  |  | • | | NCD services | * |  |  |
| Zambrano-Barragán 2021 | 21 | Patients | • | • | • | • | • |  | • |  | • | • | • | • | • |  | • |  | • |  |  | • | • | • |  | |  |  | • | | Healthcare access of Migrant workers | * |  |  |
| Soerjomataram 2022 | 22 | Patients |  | • |  |  |  | • |  | • |  |  | • |  |  |  | • |  |  | • |  |  |  | • |  | |  |  | • | | Cancer registries | * |  |  |
| Goncalves 2022 | 23 | Patients |  |  |  |  | • |  |  |  |  | • |  |  |  |  | • |  |  | • |  |  |  |  |  | |  |  | • | | AI in LC screening and diagnosis |  |  | * |
| Miranda-Olivares 2021 | 24 | Patients |  |  |  | • |  |  |  |  | • | • | • |  |  |  | • | • |  |  |  |  |  | • |  | |  |  | • | | Cancer screening and diagnosis | * |  |  |
| Aranda 2022 | 25 | Patients |  | • | • | • |  |  |  |  | • | • |  |  |  |  |  |  |  |  |  |  | • | • |  | |  |  | • | | maternal health services | * |  |  |
| Medina Guillen 2020 | 26 | HCW |  | • | • | • |  | • |  | • | • | • |  |  |  |  |  |  |  |  | • |  | • | • |  | |  |  | • | | Occupational exposure  Occupational challenges | *  * |  |  |

Table Notes: HCW, Health Care Workers,; NA, Not Applicable

1. Costa Rica, Cuba, El Salvador, Guatemala, Mexico, Dominican Republic
2. Argentina, Bolivia, Brazil, Chile, Colombia, Ecuador, Salvador, Peru, Uruguay
3. Argentina, Chile, Colombia, Ecuador
4. Argentina, Bolivia, Brazil, Chile, Colombia, Costa Rica, Cuba, Dominican Republic, Ecuador, El Salvador, Guatemala, Honduras, Mexico, Nicaragua, Panama, Paraguay, Peru, Suriname, Uruguay, and Venezuela
5. Argentina, Chile, Costa Rica, Ecuador, Ecuador-inland, Ecuador-coast, Nicaragua, Puerto Rico, Dominican Republic, and Venezuela were in group 1. In group 2 were: Bolivia, Brazil, Brazil-inland, Cuba, Guatemala, Honduras, Paraguay, Peru, and Uruguay.
6. Mexico, Chile, Haiti
7. Argentina, Bolivia, Brazil, Chile, Colombia, Ecuador, Mexico, and Puerto Rico
8. Argentina, Bolivia, Brazil, Chile, Colombia, Costa Rica, Cuba, Ecuador, El Salvador, Guatemala, Honduras, Mexico, Nicaragua, Panama, Paraguay, Peru, the Dominican Republic, Uruguay, Venezuela
9. Argentina, Brazil, Colombia, Mexico, Peru, Barbados, Jamaica, and Trinidad and Tobago
10. Peru, Chile, Paraguay, Mexico, Colombia, Bolivia, Panama, Ecuador, Costa Rica, El Salvador, Honduras, Guatemala, Venezuela, Argentina, Brazil
11. Latin America, consisting of 33 countries or territories
12. Several LAC
13. Côte d'Ivoire, Cameroon, Rwanda, Bangladesh, Zambia, Bhutan, India, Honduras, Morocco, Paraguay, China, Thailand, Brazil, Sri Lanka, Iran, Malaysia, Romania
14. South America (Argentina, Bolivia, Brazil, Colombia, Chile, Ecuador, Paraguay, Peru, Uruguay, and Venezuela); Mexico, and Central America and the Caribbean (Costa Rica, Cuba, Dominican Republic, El Salvador, Guatemala, Honduras, Nicaragua, Panama, and Puerto Rico)
15. Multinational including Mexico
16. Argentina, Colombia, Mexico, Ecuador, Venezuela, Paraguay, Peru, Honduras Cuba
17. Israel, Uruguay, and worldwide
18. Argentina, Bolivia, Brazil, Chile, Colombia, Ecuador, Paraguay, Peru, Uruguay, and Venezuela.
19. Bolivia, Brazil, Chile, Colombia, Cuba, Uruguay, Venezuela
20. LACs
21. Colombia, Peru
22. Latin America and the Caribbeans
23. Developing and developed countries
24. Global Scale
25. Haiti, Lesotho, Liberia, Malawi, Mexico, Sierra Leone
26. Mexico, Honduras, Colombia, Costa Rica, Argentina, Peru, Guatemala, Chile, El Salvador, Ecuador, Bolivia, Nicaragua, and Venezuela.

Table e6. Clinical conditions in the intervention studies reviewed in LAC and Non-LAC countries

| **Clinical Conditions** | **LAC (n=61)** | **Non-LAC (n=63)** |
| --- | --- | --- |
| Diabetes | 18 (29.5%) | 13 (20.6%) |
| Hypertension | 10 (16.4%) | 6 (9.5%) |
| Mental Health | 12 (19.7%) | 9 (14.3%) |
| General medical encounters | 14 (23.0%) | 23 (36.5%) |
| Maternal health | 4 (6.6%) | 7 (11.1%) |
| Cancer screening | 3 (4.9%) | 3 (4.8%) |

**Figure e10. Intervention: Clinical condition addressed, by Latin America and the Caribbean (LAC) countries**

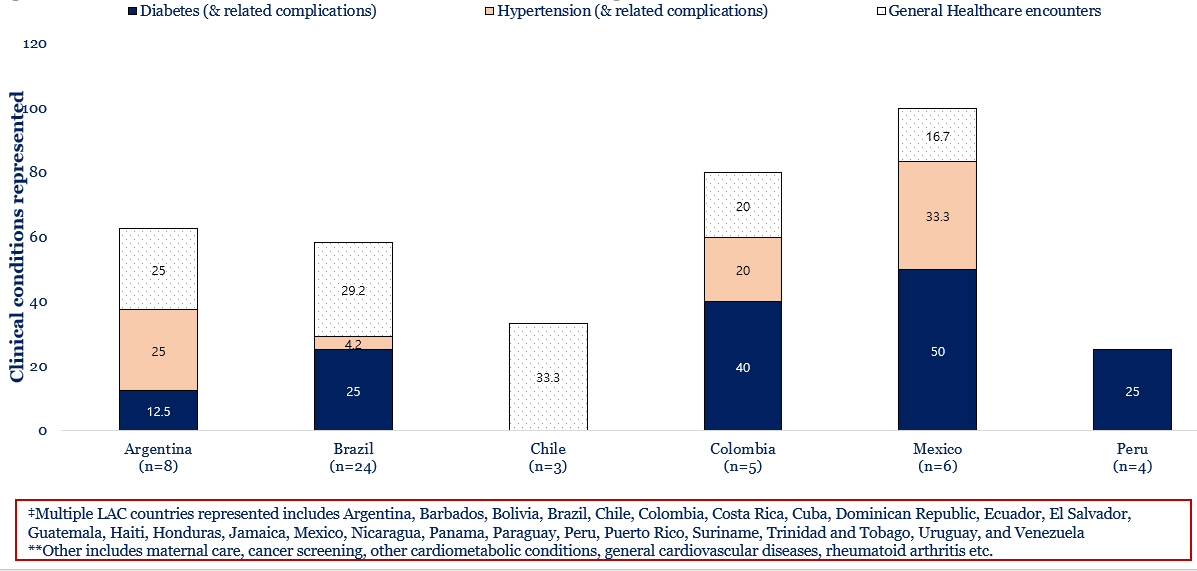

**Table e7. Characteristics of studies on interventions that may be adapted to address primary care services impacted during the COVID-19 pandemic.**

| **Author, year** | **Country** | **Participants** | **System-level domains** | | | | | | **Service delivery** | | | | | | **Practice performance** | | | | | **Consequences** | | | | | **Condition** | | | | **Int. Type** | **Outcomes** |  |  |  |  |
| --- | --- | --- | --- | --- | --- | --- | --- | --- | --- | --- | --- | --- | --- | --- | --- | --- | --- | --- | --- | --- | --- | --- | --- | --- | --- | --- | --- | --- | --- | --- | --- | --- | --- | --- |
|  |  |  | Governance/leadership | Finance/Funding | Drug/Supplies | Infrastructure | Information System | Workforce | Pop Health Mgt | Facility Org | Access/Availability | Quality | Screening/Lab | Patient perception | Process measures | Awareness | Detection/Diagnosis | Treatment | Control | Health status | Responsiveness | Equity | Efficiency | Resilience | Diabetes | Hypertension | Complications | Other |  |  | Care/ outcomes improved | No change to usual care | No mitigation/ recovery achieved | Mitigation: results limited as no comparison to usual care |
| Latin America |  |  |  |  |  |  |  |  |  |  |  |  |  |  |  |  |  |  |  |  |  |  |  |  |  |  |  |  |  |  |  |  |  |  |
| **Mesoamerica** |  |  |  |  |  |  |  |  |  |  |  |  |  |  |  |  |  |  |  |  |  |  |  |  |  |  |  |  |  |  |  |  |  |  |
| Cuevas-Budhart 2022 | Mexico | Patients |  |  |  | • |  |  |  |  | • |  |  |  |  |  |  | • |  |  |  |  | • |  |  |  |  | • | Digital health | High patient satisfaction  Geographic and financial access barriers to care were reduced (remote care vs. travel to hospital) | • |  |  | • |
| Doubova 2022 | Mexico | NA | • |  |  | • | • |  | • | • | • | • |  |  |  |  |  |  | • |  | • |  | • |  |  |  |  | • | Policy | Facility deliveries increased  Diabetes control increased  Hypertension control increased | •  •  • |  |  |  |
| Domi­nguez-Moreno 2021 | Mexico | Physicians |  |  |  |  | • | • | • |  |  | • |  | • |  |  | • | • | • |  |  |  | • | • |  |  |  | • | Digital health | Good patient & provider satisfaction  Clinical skills were challenged |  |  |  | •  • |
| Gallardo-Rincon 2022 | Mexico | Patients |  |  |  | • | • |  | • |  |  |  |  |  |  |  | • |  | • |  |  | • | • |  | • | • |  |  | Digital health | Identified obesity and hypertension and pre-hypertension  Awareness and control of NCD heath status increased |  | •  • |  |  |
| Mendoza 2022 | Mexico | Patients; HCW |  |  |  |  | • |  |  | • | • |  |  |  |  |  | • | • | • |  | • | • |  |  | • | • |  |  | Digital health | No. of consultations increased  Supports selection of patients for referral to specialist care or hospital |  |  |  | •  • |
| Monraz-Peréz 2021 | Mexico | NA |  | • |  | • | • |  |  | • | • | • |  |  |  |  |  |  |  |  | • |  | • | • |  |  |  | • | Digital health | Continuity of care, diagnosis & control  Rural access |  |  |  | •  • |
| Robles 2022 | Mexico | HCW |  |  |  |  |  | • |  | • |  |  |  |  |  |  |  |  |  |  |  |  |  | • |  |  |  | • | Digital health | Anxiety decreased  Depressive symptoms decreased  Health anxiety/ somatization decreased | •  •  • |  |  |  |
| Silva-Tinoco 2021 | Mexico | Patients |  |  |  | • | • |  |  | • | • | • |  | • |  |  |  | • | • |  | • | • | • |  | • |  |  |  | Digital health | No. of consultations increased  Perception of telemedicine care usefulness increased | •  • |  |  |  |
| **The Carribbean** |  |  |  |  |  |  |  |  |  |  |  |  |  |  |  |  |  |  |  |  |  |  |  |  |  |  |  |  |  |  |  |  |  |  |
| Quimby 2022 | Barbados | Patients |  |  | • | • |  | • | • | • | • | • |  |  |  |  |  | • | • | • |  |  | • |  | • |  |  |  | Digital health | Fidelity: visit rate  Weight loss  Systolic blood pressure  Diastolic blood pressure  Reduction in need for antihypertensive medications  Diabetes remission (FBG of <7mmol/L and A1C of <6.5%) |  | •  •  •  •  •  • |  |  |
| Morillo 2021 | Dominican Republic | Patients (pediatric) |  |  |  |  | • |  |  | • | • |  |  | • |  |  |  | • |  |  | • |  |  | • |  |  |  | • | Digital health | Good service perception by users |  |  |  | • |
| Polanco 2021 | Dominican Republic National | Patients | • |  |  | • |  | • |  | • | • |  | • |  | • |  | • | • |  | • |  |  | • | • | • | • |  | • | Digital health | Hospitalization  Hemodialysis referrals  Peritonitis rate |  | •  •  • |  |  |
| **Andean Area** |  |  |  |  |  |  |  |  |  |  |  |  |  |  |  |  |  |  |  |  |  |  |  |  |  |  |  |  |  |  |  |  |  |  |
| Betancourt-Peña 2022 | Columbia | Patients |  |  |  | • |  |  | • | • |  |  |  |  | • |  |  | • | • | • | • |  | • | • |  |  | • |  | Digital health | Cardiac physiological outcome measurements improved  Depression decreased  Clinical variables improved | •  •  • |  |  |  |
| Gomez 2021 | Colombia | Patients |  |  |  | • | • |  |  |  | • | • |  |  |  |  |  | • | • |  |  |  | • |  | • |  |  |  | Digital health | Time in range increased  Time below range decreased  Glycemic variability decreased | •  •  • |  |  |  |
| Escobar 2021 | Colombia | Patients |  |  |  | • | • |  |  | • | • | • |  | • |  |  | • | • |  | • | • | • | • |  |  |  |  | • | Digital health | Continuity of care/care provision  National coverage of care |  | •  • |  |  |
| León-Vargas 2021 | Colombia | Patients and Doctors |  |  |  | • | • |  | • | • | • |  | • |  | • |  | • | • | • | • | • |  | • | • | • |  |  |  | Mobile health app | User experience and enhancement  Online interaction and information sharing  Diabetic-specific measures of quality of life (PAID) | • | •  • |  |  |
| Hernández-Galdamez 2021 | Guatemala | Patients |  |  | • |  |  | • |  |  | • |  | • |  |  |  |  | • | • |  |  | • |  | • |  | • |  |  | Digital health | Medication access increased  Medication adherence increased  Health coaching sessions  Adherence to blood pressure monitoring increased |  |  | •  •  •  • |  |
| Alva-Arroyo 2021 | Peru | Patients, HCWs |  |  |  |  | • | • |  | • | • |  |  |  | • |  |  |  | • |  | • |  | • | • |  |  |  | • | Digital health | Care for mental health increased  Access gaps to specialist psychiatry care decreased | •  • |  |  |  |
| Ayala 2022 | Peru |  |  |  |  |  | • |  |  |  |  | • |  | • |  |  | • |  |  |  | • |  |  |  |  |  | • |  | Digital health | Successful implementation - impact on improved healthcare not clear |  |  |  | • |
| Olaza-Maguiña 2021 | Peru | Patients |  |  |  |  | • |  | • |  | • | • |  |  |  | • |  |  |  |  |  |  |  | • |  |  |  | • | Digital health | Increased knowledge of danger signs in:  Pregnancy  Childbirth  Postpartum | •  •  • |  |  |  |
| Santa-Cruz | Peru | Patients |  |  |  | • |  |  |  | • | • | • | • |  | • |  |  |  |  | • |  | • |  | • |  |  |  | • | Digital health | SRQ scores decreased  Remote physiological first aid increased  Remote Psychological First Aid plus referral to mental health services increased | •  •  • |  |  |  |
| **Southern Cone** |  |  |  |  |  |  |  |  |  |  |  |  |  |  |  |  |  |  |  |  |  |  |  |  |  |  |  |  |  |  |  |  |  |  |
| Burgos 2020 | Argentina | Patients |  |  |  |  |  |  |  |  | • | • |  | • | • |  |  |  |  |  | • |  |  | • |  | • |  |  | Digital health | Virtual visits increased  Requirement of diagnostic studies, in-person medical evaluation and hospitalization during follow-up unchanged  High patient satisfaction | • | • |  | • |
| Busso 2022 | Argentina | Patients |  |  |  | • | • |  |  |  | • |  |  |  | • |  |  | • |  |  | • | • | • | • |  |  |  | • | Digital health | Telehealth calls increased  Telehealth calls by first-time users increased  Calls initiated prescriptions  Calls resulted in resolved consultations  Calls led to specialist referral | •  •  •  • |  |  | • |
| Carro 2020 | Argentina | Patients |  |  |  | • |  |  |  |  |  |  |  |  |  |  | • | • |  |  |  |  |  | • | • |  | • |  | Digital health;  caretaker attending consul-tations | Telehealth consultations were used  Caretakers/relatives attended consultations instead of patients themselves  Diabetic foot visits were enabled  Consultation for new lesions were enabled |  |  |  | •  •  •  • |
| D’Imperio 2020 | Argentina | Health centers |  |  |  | • |  |  |  | • | • |  |  |  |  |  | • | • | • |  |  |  |  | • |  |  | • |  | Digital health | Hospitalizations due to cardiovascular diseases reduced |  |  |  | • |
| DiTommaso 2020 | Argentina | Patients; Physicians |  |  |  |  | • |  |  | • |  | • |  |  |  |  |  | • | • |  | • |  | • | • |  | • |  | • | Digital health | In-person consultations decreased  Invasive procedures decreased  Additional tests and pacemaker follow-ups were reduced | •  •  • |  |  |  |
| Falabella 2021 | Argentina | Patients |  |  |  |  |  |  |  |  | • |  |  |  |  |  |  |  | • |  | • | • |  | • |  |  |  | • | Digital health | Adherence was high  Complications- Complication rate 7%/ 1000 days  Satisfaction increased | • |  |  | •  • |
| Pedretti 2022 | Argentina | Patients, Physicians |  |  |  |  | • |  | • |  |  |  |  |  | • |  |  |  |  |  |  |  |  | • |  |  |  | • | Digital health | Digital health effectively monitored the ED readmission rates and hospitalizations rates following 14 days of Covid-19 isolation |  |  |  | • |
| Rodriguez | Argentina |  |  |  | • | • |  |  | • | • |  |  |  |  |  |  | • | • | • | • | • |  |  |  |  | • |  |  | Mobile Health | Change in hypertension control  Change in hypertension treatment levels  Change in hypertension combination therapy | •  • |  | • |  |
| Martin 2022 | Argentina | Primary care team |  |  |  |  |  | • |  | • |  |  |  |  |  |  |  |  |  |  |  |  |  | • |  |  |  | • | Mental Health | Anxiety decreased  Socio-laboral conflicts were resolved in 78%  Better management of stress, expressing mental health issues | •  •  • |  |  |  |
| Alessi 2021 | Brazil | Patients |  |  |  | • | • |  |  |  | • | • |  |  |  | • | • |  |  |  |  | • |  | • | • |  |  |  | Digital health | Maintenance of patient contact  Mental health disorders in adults with type 2 diabetes decreased  Emotional distress in adults with type 2 diabetes decreased  Eating disorders were unchanged  Sleep disorders were unchanged  Treatment adherence unchanged | •  • | •  •  •  • |  |  |
| Alves 2022 | Brazil | Patients >60 years |  |  | • |  |  |  | • |  | • |  |  | • |  |  |  | • |  |  | • |  | • |  | • | • |  |  |  | Dispensing medication to homes led to:  25% reduction in face-to-face visits  68% Satisfaction with program | •  • |  |  |  |
| Amaral 2022 | Brazil | Physicians |  |  |  |  | • | • |  |  |  |  |  |  |  |  |  |  |  |  |  |  |  | • |  |  |  | • | Digital health | Adoption of telemedicine increased  Use of electronic prescriptions increased  Patient perceptions (physician-reported) of telemedicine were acceptable |  |  |  | •  •  • |
| Aquino 2020 | Brazil | Neurologists |  |  |  |  | • | • |  | • |  | • |  |  |  |  | • | • | • |  |  |  | • | • |  |  | • |  | Digital health | More favorable perceptions on use of telemedicine |  |  |  | • |
| Aquino 2022 | Brazil | Patients | • | • |  | • | • |  |  | • | • | • | • |  | • |  | • | • | • | • | • | • | • | • |  |  |  | • | Digital health | Implementation and development |  |  |  | • |
| Bertolli 2022 | Brazil | Patients |  |  | • |  |  |  |  |  | • |  |  |  |  |  |  | • |  |  |  |  |  |  |  |  |  | • | Prescri-bing | Increased access to antiviral medication through home delivery program | • |  |  |  |
| Carneiro 2022 | Brazil | Patients |  |  |  | • | • |  |  |  | • | • |  |  | • |  |  |  |  |  | • |  |  | • |  |  |  | • | Digital health | Satisfaction with using Telehealth:  99% would use the service again,  90% had their issue resolved |  |  |  | •  •  • |
| Christinelli 2022 | Brazil | Patients with obesity |  |  |  |  | • |  | • | • |  |  | • |  |  |  |  | • | • |  |  |  | • |  |  |  |  | • | Digital health | BMI improved  Glucose improved  Insulin improved  HDL improved  Waist circumference improved  Patient perceived weight loss goals were more complicated to achieve  Reduction in physical activity  Healthy eating perceived more challenging | •  •  •  •  • |  | •  •  • |  |
| Correia 2020 | Brazil | Patients |  |  |  |  |  |  |  | • |  |  |  | • |  |  |  | • | • |  | • |  |  | • |  | • |  |  | Digital health | Teleorientation was feasible and nurses were able to support patients and provide healthcare orientations | • |  |  |  |
| Crippa 2021 | Brazil | HCW |  |  | • |  |  |  |  |  |  |  |  |  |  |  |  | • |  |  |  |  | • |  |  |  |  | • | Drug;  Mental health | Workforce emotional exhaustion, anxiety and depression was reduced | • |  |  |  |
| deMattos Matheus 2021 | Brazil | Patients |  |  | • |  |  |  |  |  | • | • |  |  |  |  | • | • | • |  |  |  | • | • | • |  |  |  | Digital health | Successful contact with T1D patients for telemonitoring during the COVID-19 pandemic.  Clinical and psychological needs were detected  Support was offered to patients | •  •  • |  |  |  |
| Fukuti 2021 | Brazil | Physicians; Nurses |  |  |  | • | • |  |  |  | • | • |  |  | • |  |  |  |  | • | • |  |  |  |  |  |  | • | Mental health | Mental health hotline triaged healthcare workers to refer them on to psychiatry care, brief supportive psychotherapy or both  A mental health app was furthermore used that encouraged 83% of users to contact the mental health hotline |  |  |  | •  • |
| Mendoz 2022 | Brazil | Patients; HCW |  |  |  | • | • |  |  | • | • | • |  |  |  |  | • | • | • |  | • | • |  |  | • | • |  | • |  | Patient rate referral to hospital was measured  Patient rate referral to specialty was measured |  |  |  | •  • |
| Ponciano 2022 | Brazil | Patients |  |  |  |  |  |  |  |  |  |  |  | • |  |  |  | • |  |  |  |  |  |  | • |  |  |  | Behavio-ral | Few difficulties maintaining some physical activity  Time flexibility, autonomy and independence were facilitators to exercise adherence  Adequate space was a barrier to adherence to the recommended exercise |  |  |  | •  •  • |
| Queiroz 2020 | Brazil | Patients |  |  |  |  |  |  | • |  |  |  | • |  |  | • | • |  |  |  |  |  | • |  | • |  |  |  | Digital health;  Team- based care | Clinical decision-making enabled through tool  Rate of referral was measured |  |  |  | •  • |
| Rossignoli 2020 | Brazil | Patients | • |  | • | • | • |  |  | • | • |  |  |  | • |  |  | • | • |  | • |  | • | • |  |  | • |  | Drug access | By adhering to established guidelines, the response effectively reduced in-person interactions at regional pharmacies while ensuring continuous user treatment. These measures successfully promoted social distancing, minimizing the risk of COVID-19 transmission for users and pharmacy teams." | • |  |  |  |
| Santana 2022 | Brazil | Patients (children with T1D) |  |  |  |  | • |  |  |  | • |  |  |  |  |  |  |  | • | • |  |  |  | • | • |  |  |  | Digital health | Hospitalization due to diabetes decompensation was unchanged  HbA1c increased |  | • | • |  |
| Scheffer 2022 | Brazil | Physicians |  |  | • |  | • |  |  | • | • |  |  |  |  |  |  |  |  |  |  | • |  |  |  |  |  | • | Digital health | For non-COVID-related care:  39% of providers used teleconsultations  30% used telemedicine for drug prescriptions and certificates  Telemedicine used for patient consultations  Younger and male providers were more likely to use telemedicine  Low telemedicine use in public primary care & outpatient services (44%) vs. private care (78%) |  |  | • | •  •  •  • |
| Sonagli 2021 | Brazil | Patients |  |  |  |  |  |  |  |  | • |  | • |  |  |  |  |  |  |  |  |  |  |  |  |  |  | • | Digital health | Telemedicine use |  |  |  | • |
| Sperling 2022 | Brazil | Patients |  |  |  |  | • |  | • |  | • |  |  |  | • |  |  |  |  |  |  | • |  |  |  |  |  | • | Digital health | Patient priority ratings for care (72%) reduced  prioritization and qualification of cases referred to specialized services improved | •  • |  |  |  |
| Tannus 2022 | Brazil | Patients |  |  | • |  |  |  |  |  |  |  |  |  |  |  |  | • | • |  |  |  |  |  | • |  |  |  | Digital health | HbA1c reduction  Overall BMI level  HbA1c improvement in patients with insulin treatment | •  • | • |  |  |
| Magario Tabuti 2022 | Brazil | Patients |  | • |  |  | • |  |  |  |  | • |  |  | • |  |  | • |  | • |  |  | • |  |  |  | • | • | Digital health | Hospital admission rates (adjusted for COVID) increased  Peritonitis unchanged  Biochemical parameters unchanged  Hospitalization for hypervolemia and infections increased |  | •  • | •  • |  |
| Delfino 2022 | Chile | Patients |  |  |  |  | • |  |  |  | • |  |  |  |  |  |  | • |  | • |  |  |  |  |  |  | • |  | Digital health | Incidence of stroke consultations/codes  Time from stroke onset to hospital admission  Number of patients receiving thrombolytic care  Decision-making to needle time | • | •  •  • |  |  |
| Garcia-Huidobro 2020 | Chile | Patients; Physicians |  |  |  | • | • |  |  |  | • |  |  | • |  |  |  | • | • |  |  |  | • | • | • |  |  |  | Digital health | Patient satisfaction similar to clinician service  Clinicians’ clinical skills challenged |  | •  • |  |  |
| Ortiz Contreras 2022 | Chile | Patients | • | • |  | • | • | • | • | • | • | • |  |  | • | • |  |  |  | • | • |  | • | • |  |  |  | • | Sexual and repro-ductive health initiatives | Mitigation strategies implemented with success  e.g. Reorganization of services, Home visits, Reorganization of health education, Use of communication technology |  |  |  | • |
| Castellano 2022 | Uruguay | HCW |  |  |  |  |  | • |  |  |  | • |  |  |  |  |  |  |  |  |  |  | • |  |  |  |  | • | Mental health | Perceived stress unchanged |  | • |  |  |
| Ferre 2021 | Uruguay | Patients; HCW |  |  |  | • | • |  |  | • | • |  |  |  |  |  | • |  | • |  | • |  |  | • | • | • |  | • | Digital health | Healthcare service use:  No. of consultations increased | • |  |  |  |
| **Multiple (LAC)** |  |  |  |  |  |  |  |  |  |  |  |  |  |  |  |  |  |  |  |  |  |  |  |  |  |  |  |  |  |  |  |  |  |  |
| Antiporta 2020 | Argentina, Bolivia, Brazil, Chile, Colombia, Ecuador, Paraguay, Peru, Uruguay, Venezuela | Mental health decision- makers in Ministry of Health | • |  | • |  |  | • | • |  | • |  |  |  | • |  |  |  |  |  |  |  |  | • |  |  |  | • | Digital health | Telemedicine was a strategy to meet mental health needs |  |  |  | • |
| Luciani 2022 | Multiple | NA | • |  |  |  |  |  |  |  |  |  |  |  | • |  |  |  |  |  |  |  |  | • |  |  |  | • | Policy | Adaptive strategies for observed NCD service disruptions helped continue care delivery  Helped maintain provider communication | •  • |  |  |  |
| Nieblas 2022 | Argentina, Brazil, Chile, Peru, Columbia, Mexico | NA | • |  |  |  | • |  |  |  | • | • |  | • | • |  |  |  |  | • |  |  |  | • |  |  |  | • | Digital health | Positive feelings towards use in patient care and management  Telemedicine during the COVID-19 in LATAM appeared effective |  |  |  | •  • |
| Camacho-Leon 2022 | Argentina, Bolivia Brazil, Chile, Columbia, Ecuador, Honduras, Mexico, Peru, UruaguayVenezuela | NA | **•** |  |  |  | **•** |  |  |  | **•** |  |  |  |  |  | **•** |  |  |  |  |  |  | **•** |  |  |  | **•** | Digital health | Access improved  Facilitated management of NCD  Cost of remote monitoring  Internet access irregular  Lack of laws for regulation | • |  | •  •  • | • |
| Other Regions |  |  |  |  |  |  |  |  |  |  |  |  |  |  |  |  |  |  |  |  |  |  |  |  |  |  |  |  |  |  |  |  |  |  |
| VieiraSilva 2022 | Worldwide | NA |  |  |  |  | • |  |  |  | • |  |  |  |  |  |  | • |  |  | • | • | • |  |  |  |  | • | Digital health | Benefits:  -continuity of care;  -accessibility (economic, social, geographical, time, and cultural)  - safety against COVID  -optimization of time;  -efficient/ practical  -reduced referrals to secondary care or hospitals  -education/ communication  Limitations:  -Access /Equity/Digital exclusion  -diagnostic imprecision/ monitoring  -confidentiality  -fragile articulation between remote and face-to-face modalities, --difficult communication with certain segments (elderly, children etc) |  |  |  | •  • |
| Garg 2021 | USA | Patients |  |  | • | • | • |  |  |  | • | • | • |  | • |  |  | • | • | • |  | • | • | • | • |  |  |  | Digital health | Effect of real-time continuous glucose monitoring on Time-in-range and glucose variability | • |  |  |  |
| Gawalko 2021 | Austria, Belgium, Germany, Denmark, Ireland, Nether-lands, Sweden, UK, Poland | Patients |  |  |  | • | • |  |  | • | • |  | • | • |  | • | • |  |  |  |  | • | • |  |  | • |  |  | Digital health | Patients reported easy use and installing  Assessment of rate and rhythm around tele-consultation was supported  Most (>80%) centers reported no problems (with cloud access, patient compliance, or quality of recordings) during implementation |  |  |  | •  •  • |
| Kruse 2022 | worldwide | Patients |  |  | • | • | • |  | • | • | • | • |  |  | • |  | • | • | • | • | • | • | • | • |  |  |  | • | Digital health | Increased satisfaction  Administrative or efficiency improved | •  • |  |  |  |
| Neves 2021 | United Kingdom, Sweden,  Italy, and Germany | Patients |  | • |  | • | • |  |  | • | • | • |  | • | • |  | • |  |  | • |  |  | • | • |  |  |  | • | Digital health | Uptake of telephone consultations  Perception that healthcare timeliness increased  Perception that efficiency increased  47% reported positive impact on effectiveness  46% reported positive impact on safety  45% reported positive impact on patient-centeredness  43% reported positive impact on equity | •  •  • |  |  |  |
| Scott 2021 | worldwide |  |  |  | • |  |  |  |  |  | • |  |  |  |  |  |  |  |  |  |  |  |  |  | • |  |  |  | Digital health | No fundamental change in medical follow-up  HbA1c, associated with positive perception of telemedicine | • | • |  |  |
| Scott 2022 | 40 countries | Patients |  |  | • |  |  |  |  |  |  | • |  | • |  |  |  |  | • |  |  |  | • |  |  |  |  | • | Digital health | Number of consultations increased  Patient satisfaction increased  Access to diabetes supplies and medication increased | •  •  • |  |  |  |
| Van Ooijen 2022 | Italy, Argentina, Malaysia and the United Arab Emirates | Patients |  |  |  | • |  |  | • |  | • |  |  |  | • |  |  |  |  | • |  | • | • |  |  |  |  | • | Digital health | Those with COVID-19- requests less likely to have had an ultrasound to determine gestation length |  |  | • |  |
| Brommeyer 2022 | Australia | Other | • |  |  |  |  |  |  | • |  |  |  |  |  |  |  |  |  |  |  |  | • |  |  |  |  | • | Digital health | Integrating digital health competencies into the training curriculum for health service managers-needed |  |  |  | • |
| DeGuzman 2022 | Australia | Other | • | • |  |  | • |  |  |  |  |  |  |  |  |  |  |  |  |  |  |  |  |  |  |  |  | • | Digital health;  Funding | GP consultations cost -introduction of additional GP telehealth funding increased average monthly cost |  |  | • |  |
| Palmer 2021 | Australia | Patients |  |  |  |  | • |  |  |  | • |  |  |  |  |  |  | • |  | • | • | • | • | • |  |  |  | • | Digital health | Safety unchanged: Pre-eclampsia  Safety unchanged: Fetal growth restriction  Safety unchanged: Gestational diabetes |  | •  •  • |  |  |
| White 2022 | Australia | General practitioners; Physicians |  |  |  | • | • |  |  |  | • |  |  |  | • |  |  |  |  |  | • |  | • | • |  |  |  | • | Digital health | Transition to telehealth advantageous but challenging  Telehealth barriers- assessing a patient’s needs, technical/legal issues related to sharing protected health information, equipment limitations, establishing when face-to-face consults are essential;  Changes in workload pressures  Causes essential modification of work practices |  |  |  | •  •  •  • |
| Johnson 2021 | Canada | HCW |  |  |  | • | • | • |  | • | • |  |  |  |  |  | • |  |  | • |  |  | • |  |  |  |  | • | Digital health | Telehealth usage  Demand for primary care  Access to primary care | •  • |  | • |  |
| Bhatti 2022 | Canada | Patients; General practitioners |  |  |  | • | • |  | • |  | • |  |  | • |  |  |  |  |  |  |  | • | • | • |  |  |  | • | Digital health | Removed certain barriers to care  Similar experience to in-person visits  Technical issues constrained care | • | • | • |  |
| Glennie 2022 | Canada | Patients |  |  |  |  |  |  |  |  |  | • |  | • |  |  | • |  | • |  |  |  |  | • | • |  |  |  | Digital health | Adult glucose monitoring improved  Child glucose monitoring improved | •  • |  |  |  |
| Ryan 2022 | Canada | Physicians |  |  |  | • | • | • |  |  | • |  |  |  | • |  |  |  |  |  | • |  | • |  |  |  |  | • | Digital health | Virtual visits and access to care increased | • |  |  |  |
| Geoffroy 2020 | France | HCW |  |  |  | • |  | • |  | • | • |  | • |  |  | • | • | • |  |  |  |  | • | • |  |  |  | • | Mental health;  Digital health | Anxiety and depressive symptoms reduced  Sleep disorders reduced  Exhaustion reduced | •  •  • |  |  |  |
| Kaushik 2021 | India | NA |  | • |  | • | • |  | • |  | **•** |  |  |  |  |  | • | • | • | • |  |  |  | • |  |  | • |  | Digital health | Telemedicine effective in cardiac care  High patient satisfaction | •  • |  |  |  |
| Meghani 2022 | India | General practitioners | • | • |  | • |  |  |  | • | • |  | • |  | • |  |  |  |  |  | • |  | • | • |  |  |  | • | Government policy | Government policies to engage and mobilize private health sector during the pandemic was variable with high engagement of private laboratories and low engagement of informal private health providers. |  |  |  | • |
| Marotta 2022 | Italy | Physicians; Nurses | • | • |  |  | • | • | • |  | • | • |  | • | • | • |  |  | • | • | • | • | • | • |  |  |  | • | Mental health | Psychological General Well-being improved  Perceived Stress reduced  Burnout: Depersonalization worsened  Emotional Exhaustion decreased  Fear of COVID-19 decreased | •  •  •  • |  | • |  |
| Dijkstra 2022 | Netherlands | NA |  |  |  | • |  |  |  | • |  |  |  |  |  |  |  |  |  |  |  |  | • |  |  |  |  | • | Digital health | Appointments delivered: unchanged  Overcrowding in hospitals’ waiting area decreased | • | • |  |  |
| Murteira 2022 | Portugal | Patients |  |  | • | • |  |  | • |  | • |  |  |  |  |  |  | • |  |  | • |  | • | • |  |  |  | • | Drug | Better access and satisfaction  Savings for citizens promoted  Reduces the burden of healthcare services | •  •  • |  |  |  |
| Alhodaib 2021 | Saudi Arabia | Policy-makers; Experts; Citizens | • | • |  | • |  |  | • | • |  |  |  | • |  |  | • | • |  |  | • |  | • |  |  |  |  | • | Policy;  Digital health | Limited ability to book medicine/medical equipment online  Personalized care decreased  Patient’s healthcare expenditures were unchanged  No change in expenditure related to travel (to healthcare service)  Access to healthcare services improved  Delivery of high-quality healthcare through digital platforms | • | •  •  • | •  • |  |
| Khan 2022 | Saudi Arabia | Patients |  |  | • |  |  | • |  |  | • |  |  | • |  | • |  | • | • |  |  |  | • | • | • |  |  |  | Team-based care | Diabetes knowledge increased  Diabetes medication adherence increased | •  • |  |  |  |
| Tourkmani 2021 | Saudi Arabia | Patients | • |  | • |  |  |  | • |  |  |  |  |  |  |  | • | • | • |  | • |  |  | • | • |  |  |  | Digital health | Glycemic control (HbA1c level) increased  In-person care visits: Successfully replaced | •  • |  |  |  |
| Dodoo 2021 | Sub- Saharan African countries | Patients; Physicians; Nurses | • | • | • | • | • | • | • | • | • | • |  | • |  |  |  |  |  |  |  | • | • |  |  |  |  | • | Digital health | Telemedicine financial barrier: cost and information and communication technology infrastructure |  |  | • |  |
| Yildirim 2022 | Turkey | Nurses |  |  |  |  |  | • |  | • |  | • |  |  | • |  |  |  |  |  |  |  | • | • |  |  |  | • | Behavio-ral;  Mental health | Mindfulness-based breathing and music therapy:  Decreased stress and work-related strain  Increased psychological well-being | •  • |  |  |  |
| Murphy 2021 | United Kingdom | Patients; General practitioners; Nurses; Practice managers |  |  |  | • | • |  |  | • | • |  |  |  | • |  |  |  |  |  |  |  | • | • |  |  |  | • | Digital health | Remote consultations increased  General perception of telehealth was positive  some risk for disparity acknowledged by providers | •  • |  | • |  |
| Papoutsi 2022 | United Kingdom | Patients; General practitioners; Nurses; Policymakers |  |  |  | • | • |  |  |  | • |  |  |  | • |  |  |  |  |  | • |  |  |  |  |  |  | • | Digital health | Addressed unmet need |  |  |  | • |
| Benis 2021 | Uruguay, Israel | Healthcare clients/ customers |  |  |  |  | • |  |  |  |  |  |  | • |  |  |  |  |  |  |  |  |  |  |  |  |  | • | Digital health | Telemedicine understood to fully replace in-person visits by patients  Patients more likely to use telemedicine if their social environment does |  |  |  | •  • |
| Adepoju 2022 | USA | Patients |  |  |  | • | • |  |  |  | • |  |  |  | • |  |  |  |  |  | • | • |  | • |  |  |  | • | Digital health | Telemedicine visits- less likely to result in a missed appointment  Chronic diseases less likely to miss telemedicine appointments | •  • |  |  |  |
| Pagan 2022 | USA | Patients |  | • |  | • | • |  |  | • | • | • |  |  | • |  | • | • | • |  | • | • | • | • |  |  |  | • | Digital health | Completion rate of primary care bookings | • |  |  |  |
| Almandoz 2021 | USA | Patients |  |  |  | • | • |  | • |  | • |  |  |  | • |  |  |  | • | • |  | • | • | • |  |  |  | • | Digital health | Older age associated with lower use of telehealth  Higher telehealth use in non-Hispanic black compared to non-Hispanic white |  |  | • | • |
| Arora 2022 | USA | Patients | * |  |  |  | * |  |  |  | • |  |  |  | * |  | * |  |  |  |  |  | • | • |  |  |  |  | Digital health | Outpatient care increased  Prescription use increased (established patients) | •  • |  |  |  |
| Baughman 2022a | USA | Patients |  |  |  | • | • |  |  |  | • | • | * |  |  |  | * | * | • |  |  |  |  | • | • |  |  | • | Digital health | Patients with telemedicine exposure had significantly better performance difference for all chronic diseases:  More testing of lipid panels (patients with CVD)  Higher testing of HbA1c (diabetes)  Higher nephropathy testing (diabetes)  Higher blood pressure control  Higher screening for cervical cancer, breast cancer and colon cancer  Higher tobacco counselling and intervention  Higher influenza vaccination  Higher pneumonia vaccination  Higher depression screening  Lower performance in 3 of 5 medication management measures | •  •  •  •  •  •  •  •  •  • |  | • |  |
| Baughman 2022b | USA | Patients |  |  |  |  | • |  |  | • |  | • | * |  |  |  | * |  | * |  | • |  | • |  |  |  |  | • | Digital health | BMI screening rate was higher using blended care (mixed office + Telemedicine consultations) vs office or telemedicine only similar  Quality performance comparable | • | • |  |  |
| Bhatia 2022 | USA | Patients | • |  |  | • |  |  |  | • |  | • |  |  |  |  | • |  |  |  | • | • |  |  | • |  |  | • | Digital health | Satisfaction with telemedicine primary care visits improved  Patients reporting a need for telemedicine | •  • |  |  |  |
| Brodar 2022 | USA | Patients |  |  |  |  | • |  |  | • | • | • | • |  | • |  | • | • |  |  | • |  |  | • | • |  |  |  | Digital health | Psychology screening rates increased  Psychology consultation rates first improved and then deteriorated | • |  | • |  |
| Burgess 2022 | USA | NA |  |  | • | • |  | • | • | • | • |  |  |  |  | • |  | • | • |  | • | • | • |  |  |  |  | • | Drug;  Prescri-bing | Medication supplies maintained  Number of staff contacts reduced  nurse time in patient rooms lowered  Antibiotic de-escalation in 57% of cases eligible | •  •  • | • |  |  |
| Chang 2021 | USA | Primary care providers (clinicians and administra-tors) |  |  |  |  | • |  |  | • | • |  |  |  |  |  |  |  |  |  |  | • |  |  |  |  |  | • | Digital health | Practice-related barriers to telehealth adoption:  Perception that quality of care via telemedicine is inferior to in-person care  Concern about reimbursement  Provider discomfort with technology  Patient-related barriers to telehealth adoption:  Patient discomfort with technology  Lack of reliable connection/device  Low use / uptake of patient portal  Language barriers  Patients not as forthcoming over video/phone |  |  |  | •  •  •  •  •  •  •  •  • |
| Contreras 2020 | USA | Patients |  | • |  |  |  |  |  |  | • |  |  |  |  |  |  |  |  |  |  |  | • |  |  |  |  | • | Digital health | Increase in telemedicine visits  Permanent regulatory solutions needed |  |  |  | •  • |
| Crowley 2022 | USA | Patients |  |  |  |  | • | • |  |  | • | • |  |  |  |  |  | • | • |  |  |  | • | • |  |  |  | • | Digital health | Diabetes distress, Diabetes self-care, and self-efficacy in patients with Persistently Poor Type 2 Diabetes improved  Body mass index unchanged  Depression unchanged  Cost slightly increased (”reasonable additional cost”) | • | •  • | • |  |
| Der-Martirosian 2022 | USA | Patients |  |  |  |  |  |  |  | • |  |  |  |  |  |  |  |  |  |  |  | • |  |  |  |  |  | • | Digital health | Telehealth use overall increased Telehealth using video increased  Telehealth differed by patient sex, patient age, provider type and rurality | •  • |  | • |  |
| Duryea 2021 | USA | Patients |  |  |  | • | • |  |  |  | • | • |  | • | • |  | • |  | • |  |  |  | • | • |  | • |  | • | Digital health | Increased number of prenatal visits attended  No change in adverse events (stillbirth, full-term NICU admission, placental abruption or arterial cord gas <ph 7.0  % with gestational hypertension unchanged  % pre-eclampsia with severe features unchanged  Pre-term birth reduced  Caesarian section reduced  Need for transfusion reduced | •  •  • | •  •  • |  |  |
| Ehmer 2022 | USA | Patients |  |  |  | • | • |  |  | • | • |  | • |  | • |  | • | • |  | • |  | • | • |  |  |  |  | • | Digital health | Number of virtual visits increased  Improvement in the attendance rates and overall increased patient contacts PROMISE clinic  No change in number of appointments completed in HEART clinic | • | • |  | • |
| Finn 2021 | USA | Patients; Administrative staff |  |  |  | • | • | • |  | • | • | • |  | • |  |  |  | • |  |  | • | • | • |  |  |  |  | • | Digital health | High acceptability of telehealth  High convenience  Patient judgement of quality significantly favored telehealth | • | •  • |  |  |
| Gao 2022 | USA | Patients |  |  |  | • | • |  | • | • | • | • |  |  | • |  | • | • |  | • | • | • | • | • |  |  |  | • | Digital health | Obstetric outcomes:  Intrapartum hospital stays unchanged  Preterm birth unchanged  Cesarean section unchanged  Newborn birthweight unchanged |  | •  •  •  • |  |  |
| Huang 2022 | USA | Patients |  |  |  |  | • |  |  |  | • |  |  |  |  |  |  |  |  |  |  | • |  |  |  |  |  | • | Digital health | Choice of video vs phone visits differed by age, ethnicity, diagnosis, SES, availability of high-speed internet, prior video/telehealth experience and depending on seeing own vs new provider |  |  |  | • |
| Iglesias 2022 | USA | Community health workers; Patients |  |  |  |  | • |  | • |  | • | • |  | • | • | • |  |  | • | • |  |  |  |  | • | • |  | • | Behavio-ral | Days exercising/week increased  Daily fruit consumption increased  Daily vegetable consumption increased  Self-reported general health increased  Depression scores were unchanged | •  •  •  • | • |  |  |
| Kerkhoff 2022 | USA | Patients |  |  |  |  |  |  | • |  | • |  | • | • | • |  | • |  |  |  |  | • | • |  | • |  |  |  | Scree-ning | Acceptability of the rapid testing program was high (friendly staff, efficiency, convenient location)  Satisfaction with screening test | •  • |  |  |  |
| Khosla 2022 | USA | Patients |  |  |  |  |  |  |  |  | • | • | • |  |  |  | • |  | • | • | • | • |  | • |  | • |  |  | Digital health | Pre-existing racial disparities in postpartum hypertension follow-up were entirely removed with teleconsultation implementation  General 6-week postpartum visit attendance  Readmission within 6 weeks after delivery | • | •  • |  |  |
| Lee 2022 | USA | Patients |  |  | • |  |  | • |  | • | • | • |  | • |  |  |  | • | • | • | • |  | • | • |  |  |  | • | Team- based care for Drug use | Healthcare facility & laboratory visits decreased  % time in therapeutic range (all patients remaining on warfarin) was unchanged  % time in therapeutic range (patients with extended ‘international normalized ratio’ monitoring interval) was unchanged | • | •  • |  |  |
| Moon 2021 | USA | Patients |  |  |  | • |  |  | • | • | • |  |  |  |  |  |  |  |  |  |  | • |  | • |  |  |  | • | Behavio-ral;  Mental health;  Digital health | Service volume increased  Participant numbers increased |  |  |  | •  • |
| Noss 2022 | USA | Patients |  |  |  | • | • |  |  |  | • |  |  |  |  |  |  |  |  |  |  | • |  |  |  |  |  | • | Digital health | Overall no-show/ cancellation rate decreased  Unchanged access in age 60+ group  Lower access in age <18y group  Access in patients identifying as Black improved | •  • | • | • |  |
| Nguyen 2022 | USA | Patient |  |  |  |  | • |  |  |  | • |  |  | • |  |  |  |  |  |  |  | • |  | • |  |  |  | • | Digital health | Patient satisfaction increased (comfort, convenience, value) |  |  |  | • |
| Ornelas 2022 | USA | General practitioners |  |  |  |  |  | • |  |  |  |  |  | • | • |  |  |  |  | • |  | • |  |  |  |  |  | • | Mental health | Telehealth and in-person program were comparable in:  High participant satisfaction  High perceived efficacy  Attendance | •  •  • |  |  |  |
| Pagán 2022 | USA | Patients |  |  |  | • |  |  |  | • | • |  |  |  |  |  |  |  |  |  | • | • |  | • |  |  |  | • | Digital health | Primary care visits increased  Access improved  Video vs. telephone modality: Disparities among some groups possible | •  • |  | • |  |
| Peahl 2021 | USA | Patients; Physicians |  |  |  | • | • |  |  | • | • |  |  | • |  |  |  | • |  | • | • |  | • | • |  |  |  | • | Digital health | 69% of patients and 96% of providers reported that virtual visits improved access to care  53% of patients and 62% of providers believed that virtual visits were safe  Nearly all believed that home blood pressure cuffs were important for virtual visits | • |  |  | •  • |
| Price 2022 | USA | Physicians |  |  |  | • | • | • |  |  |  |  | • |  |  |  |  |  |  |  |  |  |  |  |  |  |  | • | Digital health | Half of the surveyed physicians (51%) believe that there will be an increased incidence of late-stage cancer, despite the high use of telehealth.  A third of participants reported postponement of cancer screenings (breast – 35%, colon- 33%, cervical – 31%) |  |  | • | • |
| Quinton 2022 | USA | Patients |  |  |  |  |  |  |  |  |  | • |  |  | • |  |  |  |  |  |  |  | • |  | • |  |  |  | Digital health | Five quality indicators of diabetes care were comparable to pre-pandemic levels among patients receiving telemedicine, but not among those receiving in-person care only.  Accordingly, telemedicine achieved full mitigation to achieve:  systolic blood pressure <140 mmHg;  HbA1c <8.0%;  using a statin;  using aspirin;  tobacco non-use | •  •  •  •  •  • |  |  |  |
| Ramsey 2022 | USA | Patients |  |  |  | • | • |  |  | • | • |  |  |  | • |  |  | • |  | • |  | • |  | • | • |  |  |  | Digital health | Higher likelihood of returning for follow-up in-person examination if telehealth was used, independent of age, gender, race, language, residence, severity of diabetic retinopathy, and vision  Racial inequities in accessing eye examinations entirely removed among those people who had used telehealth | •  • |  |  |  |
| Reisinger-Kindle 2021 | USA | Patients |  |  |  | • | • |  |  | • | • | • |  |  | • |  | • |  |  | • | • |  | • | • |  |  |  | • | Digital health | Slightly later diagnosis of hypertensive disease  Earlier diagnosis of gestational diabetes | • | • |  |  |
| Schlegel 2022 | USA | Patients |  |  |  | • | • |  |  | • | • |  |  |  | • |  |  |  |  |  | • |  | • |  |  |  |  |  | Digital health | Number of diagnoses reported per patient consultation did not significantly differ between in-person, telephone and video consultations |  | • |  |  |
| Sepucha 2022 | USA | Patients |  |  |  | • |  | • | • | • | • |  | • |  | • | • | • |  |  | • |  |  | • |  |  |  |  | • | Behavio-ral;  Digital health | Higher shared decision-making scores  Less decisional conflict |  |  |  | •  • |
| Stephens 2022 | USA | HCW |  |  |  |  |  |  |  |  |  |  |  |  |  |  |  |  |  |  |  |  |  |  |  |  |  |  | Digital health | High satisfaction  High sense of social support | •  • |  |  |  |
| Singh 2022 | USA | HCW |  |  |  | • | • |  |  |  | • |  |  |  | • |  |  |  |  |  | • |  | • |  |  |  |  | • | Digital health | Continuity of behavioral care achieved in >85% of providers independent from the mode of telehealth delivery  Lower level of care continuity in rural regions | • |  | • |  |
| Vaughan 2021 | USA | Patients |  |  |  |  | • |  |  |  |  | • |  |  |  |  |  | • |  |  |  |  | • | • |  |  |  |  | Behavio-ral;  Diet;  Drug;  Health & safety;  Mental health;  Digital health | Reduction in HbA1c  Reduction in hypoglycemia  Reduction in weight  Reduction in medication non-adherence  Achievement in six American Diabetes Association prevention measures  Reduction in diastolic blood pressure  No change in systolic blood pressure | •  •  •  •  •  • | • |  |  |
| Yuan 2021 | USA | Patients |  |  |  | • | • |  |  |  | • | • |  |  |  |  | • | • |  |  | • | • | • | • |  | • | • | • | Digital health | Unclear whether use of remote care led to a reduction in unnecessary overuse, or whether it created unmet need:  Fewer medication orders  Lower odds of ordering electrocardiograms  Lower odds of ordering echocardiograms |  |  |  | NA |
| Tanenbaum 2022 | USA | Patients |  |  | • |  |  |  |  |  | • |  |  | • |  |  |  |  | • |  |  |  | • |  | • |  |  |  | Digital health | High-quality care not diminished  Convenience of care improved  Perceived lack of support | • | • | • |  |

Table Notes: CVD, Cardiovascular disease ; ED, Emergency department ; FBG, fasting blood plasma glucose ; GP, General Practitioner ; HCW, Health Care Workers,; NA, Not Applicable ; NCD, Non-communicable disease ; PAID, Problem Areas In Diabetes ; SRQ, Self-Reporting Questionnaire ; T1D, Type 1 diabetes

* initial increase in consultation rates to pre-COVID levels seen, but these declined during the succeeding COVID surge

**Figure e11. Disruptions and Interventions mapped to PHCPI domains and sub-domains by disease condition**

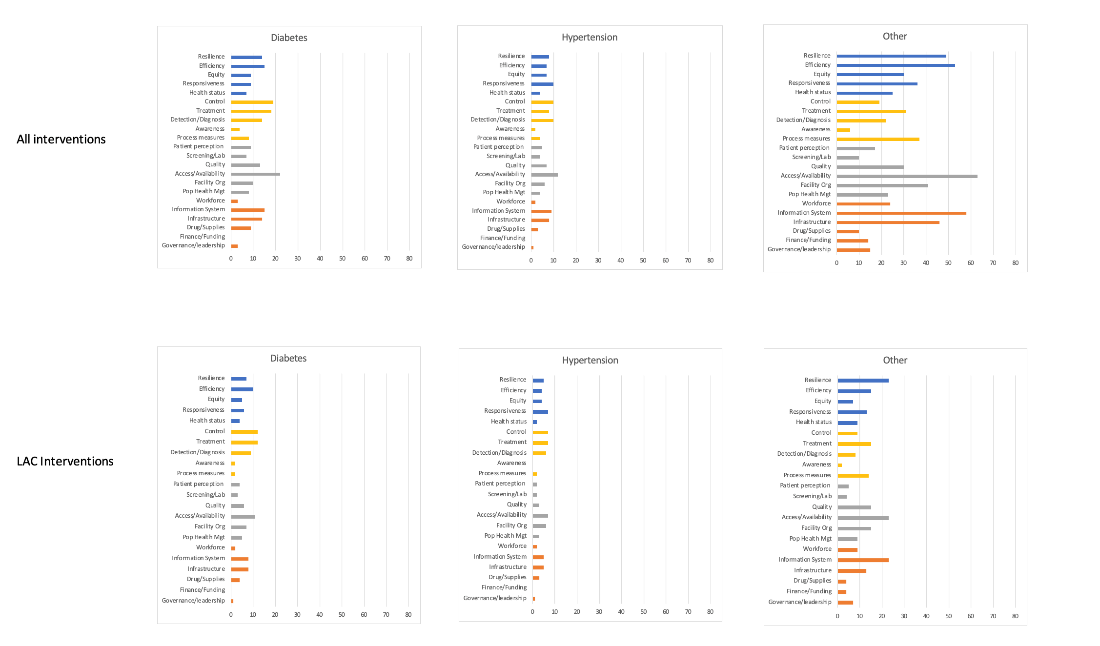

**Table e8: Description of remote care strategies studies or implemented within Latin America and the Caribbean**

| **Study** | **Country** | **Target population/ Disease state** | **Sample size** | **Intervention type** | **Setting** | **Aim of intervention** | **Clinical Outcome** | **Implementation Outcome** |
| --- | --- | --- | --- | --- | --- | --- | --- | --- |
| Falabella et al 2021 | Argentina | Patients with Enteral Nutrition | 18 patients | Telephone or videocalls | Hospital Outpatient | Monitoring | Side effects=15    Satisfaction 72.2% Good  27.7% Excellent    Professional care  66.6% V Good  22.2% Excellent  11.1% Good | - **Feasibility**  **- Acceptance**    (reduce geographic inequity and improve  accessibility to health care) |
| Betancourt-Peña 2022 | Columbia | Patients with Cardio-vascular disease (CVD) (included **hypertension**) | 31 patients | Telerehabilitation via Google meet 3 times a week + telephone calls | Specialist clinic | Monitoring  -Supervised exercise | Improvement in clinical parameters, functional capacity and depression | **- Effectiveness**    (programme judged effective) |
| Silva-Tinoco 2021 | Mexico | **Diabetes** | 192 patients  1118 consultations | Telephone calls | Primary care | Consultations | -98.4% access to telephone  73.4% computer or smartphone  69.4% internet    -263 prescriptions | **- Utility**  **- Acceptance**    (95% judged teleconsultation useful) |
| Ortiz-Contreras 2022 | Chile | Sexual and reproductive health | N/A | Tele-medicine | Primary care | Telehealth used for consultations, education, prevention and support | 41 initiatives implemented with high use of technology | **-Feasibility** |
| Morillo 2022 | Dominican Republic | Mental health | 14 patients | Telephone call or Whatsapp call | Tertiary care | Monitoring | -Patient perception of service 50% Good  -Altered sleep pattern | **- Acceptance**    (50% positive perception of healthcare service) |
| Christinelli 2022 | Brazil | Patients with obesity | 22 remote intervention  25 in person | Whatsapp call | Community healthcare clinic | Multidisciplinary remote intervention | Remote intervention improved clinical variables | **-Effectiveness** |
| Alva-Arroyo 2021 | Peru | Mental health | 75,398 teleconsul-  tations and monitoring, 4411 mental health orientations, 295 teleinterconsultations, 42 teletraining and 29 educational sessions in mental health for general population | Telephone call, videocalls and digital platform | Tertiary care | Consultation  Monitoring  Training  Education | Permitted continuity of mental health care | - **Utility**      (reduces gaps in access to specialized care in psychiatry due to COVID-19) |
| Monraz-Pérez 2021 | Mexico | N/A | N/A | Videocalls and web-based | Tertiary care | Consultation/ Monitoring  CME | Telemedicine used for training of healthcare professionals and video-consultations with patients | **-Feasibility** |
| Huancahuari-Ayala 2022 | Peru | Ophthal-mology | 308 patients | Web-based (Zoom) | Tertiary care | -Tele-orientation (triaging for face-to face visit)  -Consultation | 4.55% required face to face visit | -**Effectiveness**  **-Feasibility**    (lowered risk of exposure COVID 19) |
| Di Tommaso 2020 | Argentina | Electrophysiology | 205 patients  263 consultations | Whatsapp (calls, messages, videocall) | Public hospital | -Tele-orientation (triaging)    -Consultation | Average number consultations 7.8 messages.    1) Solved via WhatsApp: 154 patients;  2) Referred to a local hospital: 25 patients  3) Referred to specialty hospital: 26 patients | **-Utility**  **-Effectiveness**  **-Low Cost**      (reduced the number of patients exposed  to in-person consultations -  no increased cost, optimization of resources) |
| Carneiro 2022 | Brazil | General healthcare encounters    Mental health (40.4%)    Respiratory health (35.8%) | 22664 consultations | Whatsapp videocall | N/A | Tele-orientation (triaging)    Consultation | -96.0% were discharged and 4.0% were referred for face to face care  - +95.77/100 score for recommending service to family/friends | **-Utility**  **-Effectiveness**  **-Acceptance**  (ease of whatsapp videocall contributed to success and reduced risk of exposure) |
| Tabuti 2022 | Brazil | Peritoneal dialysis patients (includes patients with **diabetes)** | 747 patients | Phone call or video conference | Peritoneal dialysis centre (specialist clinics) | Monitoring | -Biochemical parameters no change signifi- cantly after transition to telemedicine.  -No assoc. between telemedicine and peritonitis rates.  -Hospitalization rates increased | -**Not effective**    (telemedicine without training can result in more adverse events) |
| Sperling 2022 | Brazil | General healthcare encounters | 622 patients | Telephone call | Primary healthcare | Triaging (managing referrals to specialty care)  Consultation | Approved referrals represented 51.9% of cases | **-Feasibility**  **-Effectiveness**    (improved equitable access to specialized services) |
| Sonagli 2021 | Brazil | Screening / Follow up Breast Cancer | 77 patients | Web based digital platforms | Tertiary care | Diagnosis and Monitoring | 36 (46.8%) follow‐up,  20 (26%)  screening,  10 (13%) benign breast disease evaluation, 7 (9%) second opinion,  and 4 (5.2%) for general info | **-Feasibility**  **-Effectiveness**      (reduce in person appointments during pandemic  45 (58.4%) breast image exams and  30 (39%) in person appointment) |
| Scheffer 2022 | Brazil | Physician | 1183 physicians | Not specified | N/A | -Used for meetings and CME    -Consult-ations with patients | Telemedicine used to  discuss clinical cases -54.9%,  for in-service meetings 48.1% and in CME 39.7%,    30.6% tele-consultation with patients | **-Feasibility** |
| Santana 2022 | Brazil | **Pediatric Type 1 diabetes** | 143 patients | Telephone calls, messages | Pediatric hospital | Monitoring | -Sending glycemic controls,  checking insulin doses,  presence of acute complications    - HbA1c values  became worse in 46% of patients | -**Feasibility**  **- Not Effective**    (worsening control) |
| Robles 2022 | Mexico | Mental health | 26 healthcareworkers | No specified | Tertiary care (psych-iatric hospital) | Consultation (Telepsycho-therapy) | -4.6 sessions average    -Intervention had a significant effect,    -Majority of HCW were  ‘‘totally satisfied’’ | **-Utility**  **-Effectiveness**  **-Acceptance** |
| Quieroz 2020 | Brazil | **Type 2 diabetes** | 627 | Smartphone based retinal camera | Primary Care | Monitoring | -333 with no DR, 40 with non-  referable DR and 66 with referable DR | -**Feasibility**  **-Low Cost**  -**Effectiveness**    (remote teleophthalmology increases DR screening in underserved área) |
| Polanco 2020 | Dominican Republic | Peritoneal Dialysis patients (includes patients with **diabetes**) | 946 patients | Whatsapp calls or telephone calls. Photographs submitted through whatsapp | Tertiary Care | -Monitoring | -8720 (91%) virtual consultations  - hemodialysis transfer rate 0.29%, peritonitis rate 0.11episode per patient/yr  -18 patients COVID + (2%) | -**Utility**  **-Effectiveness**    (monitoring, controlled complications, and PD lives protected). |
| Nieblas 2022 | Argentina, Brazil, Chile, Peru, Columbia, Mexico | General healthcare encounters | Systematic review, 24 papers | Multiple modalities | N/A | -Monitoring  -Consultation  -Education | -Generally Effective    -Patient and provider acceptance | -**Effectiveness**  **-Acceptance**    (~70% that expressed a posi-  tive feeling) |
| Mendoza 2022 | Mexico | General healthcare encounters (includes **diabetes and hypertension**) | 7,472 calls | Mobile text messages and  Video call | Hospital based | -Monitoring  -Triaging  -Consultation  -Counselling  -Prescriptions | 7,163 calls  respiratory symptoms,    1,514 for therapeutic  monitoring (refill of prescriptions),    and 8,183 for other dis-  orders (mental health) | **-Utility**    (improve access to healthcare) |
| Martin 2022 | Argentina | Mental Health | 68 Healthcare workers | Psychological intervention (in person (4), zoom/ virtual platform (3), Whatsapp videocall (10) Whatsapp call, (5) Whatsapp message (4), Telphone call (26)) | Primary Care | -Counselling | - Improvement in commun-ication and selfcare, more proactive attitude, reduced anxiety and stress    - Satisfaction  Excelent- 14 Very good- 16 Good- 11 Adequate -1 Inadequate - 3 | -**Effectiveness**  **- Acceptance**      (Access to mental healthcare) |
| León-Vargas 2021 | Columbia | **Diabetes** | 6 patients + 2 endocrinologists | Tidepool Web based App | Specialist care | -Monitoring | -A score of 81.3 in the  system usability scale for patients  - PAID score (17.5 (13-37)) was lower vs. baseline (21.5 (13-28)), less distress, although no significant changes were detected | -**Feasibility**  **-Acceptance** |
| Hernández-Galdame 2021 | Guatemala | **Hypertension** | 677 Ministry of Health Intervention group vs. 605 Control group | Telephone calls | Healthcare centre | -Monitoring  -Education  -Medication access | -Delivery of anti-hypertensive medications,73% intervención group vs. 51% Controls  -Adherence greater in intervention group (80% vs 65%)    -Healthcare coaching sessions (62% in intervention gp)    -Use of a home blood pressure monitoring (81% in intervention group) | -**Feasibility**    (access to antihypertensive medications  and health services impaired less than expected) |
| Gomez 2021 | Columbia | **Diabetes** | 91 patients with type 1 diabetes switched to hybrid closed loop | Web based programme (Zoom video conference) | Specialist centre | -Virtual training/education    -Monitoring | Mean TIR improved  81.6% ± 7.6  -TBR <70 mg/dL reduced  (2,7% ± 2,28 vs 1,83% ± 1,67,)  -glycemic variability,32.4 %vs 29.7% | **-Effectiveness**  **-Feasibility**    (improvement in control and safety confirmed) |
| Garcia-Huidobro 2020 | Chile | General healthcare encounters | 3962 Patients and 263 Physicians | Web based programme (Zoom video conference) | Primary and hospital care | Consultations | Patient and physician perception and satisfaction  -patients reported similar satisfaction to in person visits  -physicians 61.8% clinical skills challenged | -**Lower Acceptance among physicians**      (patient and provider) |
| Fukuti 2021 | Brazil | Mental health | 395 Healthcare workers | Telephone and  Web based  Consultation + mobile phone app for screening | Tertiary care/ Hospital | Consultations | Symptoms of anxiety,  depression and sleep disturbances.    -Diagnosis of Adjustment, Anxiety, and Mood  disorders    - 88.6%  would recommend program | **-Feasibility**  -**Acceptance** |
| Escobar 2021 | Columbia | General healthcare encounters | 56,560 teleconsultations | Web based  Consultation (Microsoft Teams video-call) | Tertiary care/ Hospital | Consultations | Patient satisfaction - excellent/good 98.32% and  excellent 84.39% | **-Acceptance**    **(**Barriers:  -staff- required training  -patients- age, level of education, lack of computer skills,  inadequate internet coverage) |
| Domínguez-Moreno 2021 | Mexico | Neurology patients | 304 patients | Whatsapp videocall or telephone call | Tertiary care | Monitoring | -86.4% preferred teleneurology (concerns about COVID-19)    -96% satisfied | **-Acceptance**    (Improved access to neurology service) |
| Delfino 2022 | Chile | Stroke patients (complication of **hypertension**) | 1078 patients | Videocall via mobile phone app | Tertiary care | Consultation | Telestroke team consulted from emergency room    -No difference in number of patients receiving reperfusión or time to admision vs pre-pandemic.  -Decision-making-to-needle  time was significantly shorter | **-Effectiveness**    (Telestroke team as effective during pandemic as pre pandemic) |
| deMattos Matheus 2021 | Brazil | **Type 1 diabetes** | 321 patients | Telephone calls or text messages | Tertiary care | Monitoring | -44 (18.5%) patients reported COVID-19-like symptoms    - Psychological symptoms 137 (60.4%) patients and 30 (12.7%) required remote psychological assistance | **-Feasibility** |
| Camacho-Leon 2022 | Argentina, Bolivia, Brazil, Chile, Columbia, Ecuador, Honduras, Mexico, Peru, Uruguay, Venezuela | Narrative review | N/A | Multiple modalities | N/A | Not reported | N/A | **-Feasibility**    -Address inequalities in healthcare access    -Barriers include lack of legislation,    - Logistical barriers such as power outages, lack of Internet access, and low smartphone access. |
| Busso 2022 | Argentina | General healthcare encounters | N/A  Administrative records | Calls-possibly all types | Not reported | Consultation  Monitoring  Prescriptions | -Number of calls increased by 230%, and the number of first-time callers by 198%  -General medicine and family medicine had greatest increase in calls  - Prescriptions calls increased by 332%.  -Calls that led to referrals grew by 190%. | **-Utility**    (Change in demand for telemedicine with onset of pandemic) |
| Burgos 2020 | Argentina | Heart failure, pulmonary **hypertension** and heart transplant | -During COVID, intervention group: 313 patients    -Pre-Covid, Control group: 21 | Virtual visits | Tertiary care/ hospital | Monitoring | Acceptability of vistual visit:  - easy to carry out 90%  - average rating 9.76±0.5 out of 10. | **-Acceptance** |
| Aquino 2022 | Brazil | Neurology patients | 243 teleconsultations, 77% first appointments | Telephone call or  Videocalls | Hospital | Consultation Monitoring | 20.16% first appointments, referred to  face-to-face consultation and 21.81% to primary care. | **-Utility**    **(**Effective access to neurology services) |
| Aquino 2021 | Brazil | Perceptions of Neurologists | 162 neurologists, | Mobile phone, messaging | Not reported | Consultation  Monitoring | 63.6% applying telemedicine  - tele-consultation: 62% -telediagnosis: 18%  -tele  interconsultation: 31%  -tele-monitoring: 16%  -tele-orientation: 62%  -teletriage: 11% | **-Utility**  **-Acceptance**    **(**increased among neurologists) |
| Amaral 2022 | Brazil | Physicians practicing telemedicine | 2,052 physicians | Digital platforms/ web based | Private clinics | Consultation  Monitoring  Prescriptions | 47.8% not practicing telemedicine  Of those that did, 88.33% had no training | **-Feasibility**  **-Acceptance** |
| Alessi 2021 | Brazil | **Mental health in type 2 diabetes** | 91 (46 intervention group, 45 control group) | Telehealth Intervention: telephone calls and educational materials on mental health, healthy habits, and  diabetes care | Tertiary care | Monitoring  Education | After 16 weeks, screening for mental health disorders, 37.0% in tele-intervention group vs. 57.8% control group (P=0.04) | **-Effectiveness**    (remote contact with health professionals reduces mental health disorders in Type 2 diabetes) |
| Santa-Cruz 2022 | Peru | Mental Health general population | 2027 adults | Digital tool (ChatBot-Juntos), with Self-Reporting Questionnaire (SRQ) to screen for psychological distress. Psicological first aid (PFA) provided + referral for mental healthcare. | N/A | Screening  Counselling | At 3 months.  SRQ scores↓ (median of 9 to 5 ( p < 0.001)), and for PFA + referral, SRQ score ↓  (11 to 6 ( p < 0.001)) | **-Feasibility**  **-Effectiveness:**  Digital technology use for screening and mental health support. Unsure if use of technology improves mental healthcare outcomes |
| Olaza-Maguiña 2021 | Peru | Antenatal care and education | 128 pregnant women (64 telehealth group and 64 face to face group) | Telehealth asynchronous education sessions + antenatal visits | Tertiary Care | Education  Monitoring | Knowledge of danger signs in pregnancy, childbirth and post-partum significantly greater in telehealth group | **-Feasibility**  **-Effectiveness**  telehealth intervention increased the knowledge of danger signs in pregnant women |
| Gomes de Castro 2020 | Brazil | General healthcare encounters | 329  Consultations in 3 weeks | Whatsapp messages, audio, and video calls | Primary care | Consultation  Prescriptions | -Implement-ation of telemedicine was efficient  -Positive patient satisfaction  -Permitted healthcare delivery  26% required face-to face | **-Utility**  **-Effectiveness**  **-Acceptance**    Access to rural areas |
| Da Silva Correia 2020 | Brazil | Patients with **hypertension** | 53 patients | Telephone contact with nurses | Hospital or tertiary care | Education | Basic guidance on hypertension control and COVID-19 -to avoid trips to emergency services, reducing the risk of infection | **-Feasibility** |
| Quimby 2022 | Barbados | Patients with **diabetes** | 5 patients remote and 31 face to face | Whats app and Zoom | Home care or visits | Monitoring | 12-week intervention, daily 840-kcal allowance, and weekly monitoring of weight and blood glucose.  4 participants lost 11.3-14.3kg; 1 gained 0.4kg vs. average weight loss of 6.8kg in cohort 1 All achieved diabetes remission | **-Feasibility**  **-Effectiveness** |
| Garelli 2022 | Argentina | Patients with type 1 **diabetes** | 5 patients | Insumate platform connects a smartphone with the insulin pump and glucose monitor | Phase 3 study | Monitoring | Ambulatory use of the Automatic Regulation of Glucose algorithm is feasible, safe and effective. | **-Feasibility** |
| Vieira Silva 2022 | Both LAC (Brazil) and Non-LAC | Digital strategies | N/A | Telephone calls (74%), video calls (64%), patient portal (28%), smartphone apps (13%), text messages (8%), email (8%), EMR (5%), social network (3%) | Primary Care | Consultation, prescriptions, exams,  monitoring, health guidelines, issuance of certificates, treatments,  screening, diagnosis,  referrals,  self-monitoring, and risk  classification. | Impact on quality of care mixed: positive 43%, negative 14%, both 43% | **-Acceptance**  **-Utility** |
| PAHO 2020 | Americas | Telemedicine implemented in 62% countries (14/23) | N/A | Telemedicine | N/A | Strategies to maintain non- communicable disease services | Triaging to  prioritize care, telemedicine to replace in person  consultations, and dispensing medicines | **-Feasibility** |
| Scott 2020 | 89 countries (North America 37%, South America 4%) | Telemedicine | 7477 patients with type 1 diabetes (survey responses) | Telemedicine | N/A | Use and perception  of telemedicine in people with type 1 diabetes using web-based questionnaire | -28% remote care through telephone (72%) or video-calls (28%). 86% found it useful 75% plan to continue remote care  - HbA1c, positively associated with positive perception of telemedicine. | **-Acceptance**    **-Effectiveness**  **-Utility** |
| van Ooijen 2021 | 181 countries, sub-analysis of Italy, Argentina, Malaysia and the United  Arab Emirates (UAE | Abortion requests through a telemedicine abortion service | 4962 women and pregnant people | Online consultation to request a medical abortion | N/A | Impact of the pandemic on access to abortion  care. | Pandemic limiting access to reproductive healthcare worldwide (40%  requesting an abortion  stated COVID-19 as reason) | **-Feasibility** |

**Table e9: Mental Health Interventions within Latin America and the Caribbean**

| **Study** | **Country** | **Target population/ Disease** | **Sample size** | **Intervention type** | **Setting** | **Aim of intervention** | **Clinical Outcome** | **Implementation Outcome** |
| --- | --- | --- | --- | --- | --- | --- | --- | --- |
| Santa-Cruz 2022 | Peru | Mental health of general population | 2027 | Chat-Bot juntos used to screen for psychological distress | Primary care | -Screening for psychological distress using the ChatBot-Juntos.    Difference in SRQ scores between screening and 3 months follow-up in participants who received remote Psychological first aid (PFA)  Participants with a mental health condition or safety concern referred to mental health services. | 1581 (77.9%) screened positive for psychological distress.  63% of these received PFA  32.1% referred to mental health services  only PFA: median SRQ score changed from 9 to 5 ( p < 0.001)]  PFA+ referral: median SRQ score changed from 11 to 6 ( p < 0.001) | **Feasibility**  **Effectiveness** (more research needed to determine if this contributes to improved mental outcomes) |
| Santos 2022 | Brazil | Healthcare workers | 12 community health workers | Administration of auriculotherapy, reiki, and aromatherapy | Community healthcare | Analyze the repercussions of comple-mentary practices on health professionals during the pandemic | The workers felt valued, improved mental and emotional well-being, reducing anxiety, improved quality of life, motivating the staff and enhancing the quality of care provided. | **Feasibility**  No control group |
| Morillo 2022 | Dominican Republic | Users/ Patients attending childrens hospital | 44 mental healthcare patients | Virtual follow up of patients | Tertiary care | Evaluation of virtual follow up of mental health services provided by the paediatric hospital | Perception of service: 55% good, 45% bad, 5% regular.  Insomnio 32% Increased number of hours of sleep 20% Combination 20%  Waking at night 8%  Contact with department of mental health 52% | **Feasibility**  **Acceptance patients** |
| Miliauskas 2022 | Brazil | Mental health patients | 2428 patients monitored with telephone calls and 50 telehealth intervention | Monitoring with telephone calls + telehealth intervention | Community healthcare | Re-organization of healthcare delivery- Online appointments and creation of a list of mental health patients to care | Mean number of appointments per week in a 12 week period: 5.08 pre and 4.16 post pandemic | **Utility** |
| Alva-Arroyo 2021 | Peru | Mental health patients | -75,398 teleconsul-  tations and monitoring, -4411 mental health consultations,  -295 teleinterconsultations, -42 teletraining -and 30 videos in internet for general population | Telephone call, videocalls and digital platform | Tertiary care | Consultation  Monitoring  Training  Education | Permitted continuity of mental health care | **Utility** |
| Castellano 2022 | Uruguay | Healthcare workers | 15 healthcare professionals | On-line mindfulness-based stress reduction program | Tertiary care | Feasibility of On-line mindfulness-based stress reduction program for health workers | Statistically significant improvement in the Cohen Scale scores post-intervention, with a decrease of 8 points (p-value <0.007) at 3 months | **Feasibility** |
| Robles 2022 | Mexico | Healthcare workers | 26 (total: 31; only 26 with post-treatment tests completed) | Tele psychotherapy  -standard CBT for anxiety and depression,  -standard CBT for somatic symptoms,  -techniques for coping  COVID-19-stress | Tertiary care | Remote psychological intervention, in particular implementation of CBT (cognitive behavioural therapy) | The number of sessions was 4.6 (2.43).  253 HCW were referred for treatment; only 22.2% (n = 56) began treatment. Of these, 25 (44.6%) dropped out, and 31 (55.4%) completed the intervention—although 5 (16.1%) failed to complete the post-intervention assessment.  Pre-post test results showed significant improvement in  -Anxiety, depressive symptoms, somatic symptoms  The intervention had a significant effect (delta Cohen’s  coefficients ‡1), and the majority of HCW were  ‘‘totally satisfied’’ with its contents and considered it ‘‘not  complex’’ (95.2% and 76.1%, respectively) | **Utility**  **Acceptance among HCW** |
| Martin 2022 | Argentina | Healthcare workers | 68 psychological iterventions | Mental health intervention (virtual) | Primary care | Health care personnel offered mental health intervention, evaluated with a satisfaction questionnaire | Improvements included better management of stress, expressing mental health issues, ask for help at work when necessary, and better relations with colleagues. | **Feasibility** |
| Fukuti 2021 | Brazil | Healthcare workers | 395 intervention group and 131 control group | Hotline + Web-based Clinical consultations and brief supportive psychotherapy sessions | Hospital-based | Program to protect healthcare workers’ mental health | Prevalence rates of anxiety (37.4%), depression (23.6%), sleep problems (30.4%), and burnout (75.8%).  Feedback from 70 respondents. Of those  respondents, 88.6% assigned a grade of 8 or higher and  affirmed that they would recommend the program to others | **Utility** |
