## Appendix 2 for "Disruption to diabetes and hypertension care during the COVID-19 pandemic in Latin America and the Caribbean and mitigation approaches: A Scoping Review"

### **Equitable Partnership Declaration**

*Researcher considerations*

1. Please detail the involvement that researchers who are based in the region(s) of study had during a) study design; b) clinical study processes, such as processing blood samples, prescribing medication, or patient recruitment; c) data interpretation; and d) manuscript preparation, commenting on all aspects. If they were not involved in any of these aspects, please explain why.

   *This question is intended for international partnerships; if all your authors are based in the area of study, this question is not applicable.*

   *This should include a thorough description of their leadership role(s) in the study. Are local researchers named in the author list or the acknowledgements, or are they not mentioned at all (and, if not, why)? Please also describe the involvement of early career researchers based in the location of the study. Some of this information might be repeated from the Contributors section in the manuscript. Note: we adhere to ICMJE authorship criteria when deciding who should be named on a paper.*

| |
|---|
| **a) Study design:** The study design was co-developed by researchers from both Latin American and Caribbean (LAC) countries and high-income countries (HICs). LAC-based researchers, including students and early career researchers, were involved in conceptualizing the research questions, adapting the search strategy, and providing input on the protocol. |
| **b) Clinical study processes:** Not applicable as this was a scoping review without clinical processes. |
| **c) Data interpretation:** LAC-based researchers, including those originally from, or lived and worked for substantial length of time in the LAC region (RM, KL, AJ, AE, IN, TA, MFFM, CS, JF, RBM, SYA) were involved in any of the following: screening studies, extracting data, assessing study quality, and synthesizing and interpreting the findings. They provided crucial contextual knowledge to inform data synthesis and reporting. The expert committee members from Barbados, Brazil, Mexico and Peru reviewed the manuscript and provided input and are listed in the acknowledgements. |
| **d) Manuscript preparation:** LAC-based researchers, who are co-authors, contributed substantially to drafting the manuscript, revising it critically for important intellectual content, and approving the final version to be published. The expert committee members from Barbados, Brazil, Mexico and Peru reviewed the manuscript and provided input and are listed in the acknowledgements. |

2. Were the data used in your study collected by authors named on the paper, or have they been extracted from a source such as a national survey? ie, is this a secondary analysis of data that were not collected by the authors of this paper. If the authors of this paper were not involved in data collection, how were data interpreted with sufficient contextual knowledge?

   The Lancet Global Health *believe contextual understanding is crucial for informed data analysis and interpretation.*

| |
|---|
| The data for this scoping review were extracted from published literature identified through systematic searches - included LAC specific databases such as LILACS (Latin American and Caribbean Health Sciences Literature), not collected directly by the authors.<br>To ensure contextual interpretation, the author team included LAC-based researchers who provided essential regional knowledge to guide data interpretation and formulation of the recommended actions. |

3. How was funding used to remunerate and enhance the skills of researchers and institutions based in the area(s) of study? And how was funding used to improve research infrastructure in the area of study?

   *Potentially effective investments into long-term skills and opportunities within institutions could include training or mentorship in analytical techniques and manuscript writing, opportunities to lead all or specific aspects of the study, financial remuneration rather than requiring volunteers, and other professional development and educational opportunities.*

   *Improvements to research infrastructure could be funding of extended trial designs (such as platform trials) and use of master protocols to enable these designs, establishment of long-term contracts for research staff, building research facilities, and local control of funding allocation.*

| |
|---|
| **Skills:** Funding supported the involvement of LAC-based researchers in the review process, enabling them to contribute to and lead various aspects of the study.<br>This provided opportunities to enhance research - systematic review of the literature and manuscript writing skills. |
| **Research infrastructure:**<br>No direct funding was used to improve research infrastructure in LAC for this review. |

4. How did you safeguard the researchers who implemented the study?

   *Please describe how you guaranteed safe working conditions for study staff, including provision of appropriate personal protective equipment, protection from violence, and prevention of overworking.*

| |
|---|
| As a scoping review, this study did not involve primary data collection that may have placed local researchers at risk. All researchers participated voluntarily as part of the author team. |

*Benefits to the communities and regions of study*

5. How does the study address the research and policy priorities of its location?

   *How were the local priorities determined and then used to inform the research question? Who decided which priorities to take forward? Which elements of the study address those priorities?*

| |
|---|
| The research topic, disruptions to NCD care during the pandemic, was identified as a priority issue for LAC countries by PAHO and LAC-based researchers and policymakers engaged in the study.<br>The findings aim to guide strategies to build more resilient health systems in LAC. |

6. How will research products be shared in the community of study?

*For instance, will you be providing written or oral layperson summaries for non-academic information sharing? Will study data be made available to institutions in the region(s) of study?* The Lancet Global Health *encourages authors to translate the summary (abstract) into relevant languages after paper editing; do you intend to translate your summary?*

> The results will be disseminated through an open-access publication to enable global access, including for LAC researchers and institutions.
> A policy brief will be developed, published, and distributed through PAHO and our expert committee members, to facilitate dissemination to governments of the region and decision makers.
> Lay summaries will be developed and the abstract will be translated into Spanish and Portuguese.

7. How were individuals, communities, and environments protected from harm?

a) *How did you ensure that sensitive patient data was handled safely and respectfully? Was there any potential for stigma or discrimination against participants arising from any of the procedures or outcomes of the study?*

> As a scoping review of aggregate, published data, this study did not involve human subjects or the collection of sensitive personal data that could lead to stigma or discrimination.
> No environmental impacts occurred.

b) *Might any of the tests be experienced as invasive or culturally insensitive?*

> Not Applicable

c) *How did you determine that work was sensitive to traditions, restrictions, and considerations of all cultural and religious groups in the study population?*

> Not Applicable

d) *Were biowaste and radioactive waste disposed of in accordance with local laws?*

> Not Applicable

e) *Were any structures built that would have impacted members of the community or the environment (such as handwashing facilities in a public space)? If so, how did you ensure that you had appropriate community buy-in?*

> Not Applicable

f) *How might the study have impacted existing health-care resources (such as staff workloads, use of equipment that is typically employed elsewhere, or reallocation of public funds)?*

> Not Applicable

8. Finally, please provide the title (eg, Dr/Prof, Mr/Mrs/Ms/Mx), name, and email address of an author who can be contacted about this statement. This can be the corresponding author.


**Name:** Dr. Kunihiro Matsushita
